# A systems immunology study comparing innate and adaptive immune responses in adults to COVID-19 mRNA (BNT162b2/mRNA-1273) and adenovirus vectored vaccines (ChAdOx1-S) after the first, second and third doses

**DOI:** 10.1101/2022.09.22.22280180

**Authors:** Feargal J. Ryan, Todd S. Norton, Conor McCafferty, Stephen J. Blake, Natalie E. Stevens, Jane James, Georgina L. Eden, Yee C. Tee, Saoirse C. Benson, Makutiro G. Masavuli, Arthur EL Yeow, Arunasingam Abayasingam, David Agapiou, Hannah Stevens, Jana Zecha, Nicole L. Messina, Nigel Curtis, Vera Ignjatovic, Paul Monagle, Huyen Tran, James D. McFadyen, Rowena A. Bull, Branka Grubor-Bauk, Miriam A. Lynn, Rochelle Botten, Simone E. Barry, David J. Lynn

## Abstract

We longitudinally profiled immune responses in 102 adults who received BNT162b2 (Pfizer-BioNTech) or ChAdOx1-S (Oxford-AstraZeneca) as their primary vaccinations. Bloods were collected pre-vaccination, 1-7 days after the 1^st^, 2^nd^ and 3^rd^ doses (BNT162b2 or mRNA-1273) to assess innate and early adaptive responses, and ∼28 days after the 2^nd^ and 3^rd^ doses to assess immunogenicity. Using a multi-omics approach including RNAseq, cytokine multiplex assay, proteomics, lipidomics, and flow cytometry we identified key differences in the immune responses induced by the ChAdOx1-S and BNT162b2 vaccines that were correlated with subsequent antigen-specific antibody and T cell responses or vaccine reactogenicity. We observed that vaccination with ChAdOx1-S but not BNT162b2 induced a memory-like response after the first dose, which was correlated with the expression of several proteins involved in complement and coagulation. The COVID-19 Vaccine Immune Responses Study (COVIRS) thus represents a major resource to understand the immunogenicity and reactogenicity of these COVID-19 vaccines.

## Background

The BNT162b2 (Pfizer-BioNTech), ChAdOx1-S (Oxford-AstraZeneca), and mRNA-1273 (Moderna) vaccines are highly effective at preventing severe COVID-19 (Falsey et al., 2021, Polack et al., 2020, Baden et al., 2021), though protection against infection wanes within 4-6 months after the second dose and two doses provide reduced protection against infection by more recent SARS-CoV-2 variants (Lustig et al., 2022, Andrews et al., 2022). A third dose of mRNA vaccine is recommended and has been shown to induce superior immunogenicity and effectiveness (Tseng et al., 2022, Yoon et al., 2022, Lustig et al., 2022). The adaptive immune responses induced by these vaccines including anti-Spike and anti-Receptor-Binding Domain (RBD) binding antibody titers, neutralizing antibody titers, memory B cell responses, and T cell responses are increasingly well understood (Feng et al., 2021, Levin et al., 2021, GeurtsvanKessel et al., 2022, Zhang et al., 2022). Recent studies are also shedding light on how immune responses induced in the hours and days post-vaccination relate to subsequent immunogenicity (Arunachalam et al., 2021, Provine et al., 2021, Takano et al., 2022). For example, frequency of non-classical monocytes, dendritic cells (DC), natural killer (NK) cells, and NKT-like cells following the second dose of BNT162b2 were negatively correlated with neutralizing antibody titers (Takano et al., 2022). In contrast, activation of mucosal-associated invariant T (MAIT) cells has been shown to be positively correlated with T cell responses induced by ChAdOx1-S (Provine et al., 2021). Despite these efforts there are still significant gaps in our knowledge of the immune processes underpinning the differences in immunogenicity between vaccines. BNT162b2 and ChAdOx1-S are also associated with relatively high levels of reactogenicity ranging from self-resolving symptoms (e.g. pain at the injection site, headache, fever, chills, and myalgia) to rare, serious adverse events (AEs) such myocarditis (Patone et al., 2022, Mevorach et al., 2021, Barda et al., 2021), or thrombosis with thrombocytopenia syndrome (TTS; also known as vaccine-induced immune thrombotic thrombocytopenia (VITT) (Greinacher et al., 2021, Schultz et al., 2021, Scully et al., 2021, Simpson et al., 2021), respectively. To date, no studies have systematically compared innate and early adaptive immune responses to vaccination with BNT162b2 and ChAdOx1-S at a systems level in the same cohort, nor has it been investigated how immune responses induced immediately after the third dose compare to those following the first and second.

Here, we report the results of the COVID-19 Vaccine Immune Responses Study (COVIRS), a systems vaccinology study in which we have longitudinally profiled early and late immune responses after the first, second, and third doses of vaccine in a South Australian cohort of 102 healthy adults who received the BNT162b2 (mRNA) or ChAdOx1-S (adenovirus vectored) vaccines as their first vaccinations in the absence of community transmission of SARS-CoV-2. A subset of participants were also assessed following a third dose of either the BNT162b2 or mRNA-1273 vaccines

## Results

To investigate the relationship between early innate and adaptive immune responses induced post-vaccination (p.v.) and vaccine immunogenicity and reactogenicity, 146 adults living in Adelaide, South Australia were recruited in 2021 to the COVID-19 Vaccine Immune Responses Study (COVIRS). The 1^st^ and 2^nd^ doses of vaccine were administered in the absence of SARS-CoV-2 community transmission due to strict quarantine and border control measures in place in South Australia. Forty-four participants withdrew prior to sample collection. The remaining participants received two doses of either the BNT162b2 (n=86) or ChAdOx1-S (n=16) vaccines for their 1^st^ and 2^nd^ doses. Samples were also collected from a subset of participants who received a 3^rd^ dose of either BNT162b2 (n=38) or mRNA-1273 (n=14) **(Fig. 1A)**. Participants were 70% female with a mean age of 39 years at enrolment (standard deviation (SD) ± 11 years). There was no significant difference between those who received 2 doses of the BNT162b2 compared to those who received 2 doses of ChAdOx1-S in terms of their age, sex, body mass index, comorbidities (e.g. hypertension, asthma), self-reported alcohol consumption or cigarette smoking **(Fig. 1B, Table S1)**. Participants completed a detailed survey (**Supplementary File 1**) to report any adverse events (AEs) one week after each vaccine dose (>95% completion rate).

**Figure 1.**
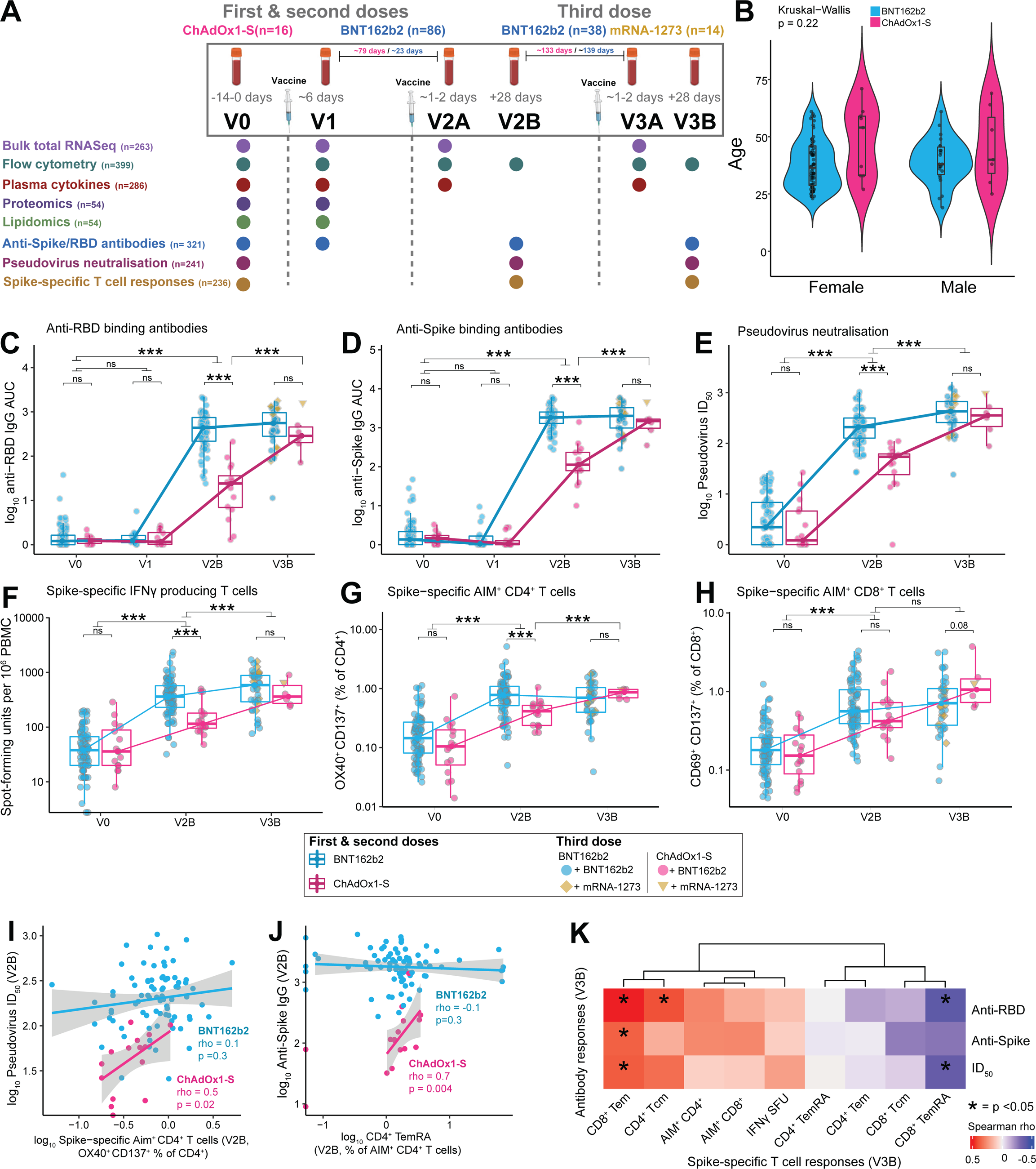
Antigen-specific antibody and T cell responses in adults vaccinated with the BNT162b2, mRNA-1273 or ChAdOx1-S vaccines. (A) Sample collection timepoints and assays performed. Assay sample numbers represent the total number of samples analyzed. (**B)** Age and sex of participants. **(C)** Anti-RBD and **(D)** anti-Spike IgG titers, **(E)** pseudovirus neutralizing antibody titers (ID_50_), **(F)** Spike-specific IFNγ spot-forming units (SFU), **(G)** Spike-specific AIM^+^ CD4^+^ T cells, (**H**) Spike-specific AIM^+^ CD8^+^ T cells, pre- and post-vaccination with BNT162b2 or ChAdOx1-S (1^st^& 2^nd^ doses) and 3^rd^dose of either BNT162b2 or mRNA-1273. **(I)** Correlation (Spearman) between ID_50_ and the proportion of Spike-specific AIM^+^CD4^+^ T cells at V2B, in participants vaccinated with BNT162b2 or ChAdOx1-S. **(J)** Correlation (Spearman) between anti-Spike IgG titers and the proportion of Spike-specific CD4^+^ TemRA cells (CD45RA^+^CCR7^-^) at V2B. **(K)** Heatmap of Spearman correlations between Spike-specific T cell responses and antibody responses at V3B (combined data for all vaccines). Data in B-H are represented as violin (B) or boxplots (C-H), with the box denoting the 25^th^ and 75^th^ percentiles, the whiskers the 5^th^ and 95^th^ percentiles, the middle bar is the median. Statistical significance was assessed in B using a Kruskal-Wallis test, in C-H using Wilcoxon Rank Sum tests. * P < 0.05, *** P < 0.001, ns = not significant.

### Three doses of COVID-19 vaccines induce high levels of protective antibodies and Spike-specific T cells in healthy, SARS-CoV-2 naïve adults irrespective of the first vaccine

Anti-Spike and anti-RBD total IgG titers and pseudovirus neutralizing antibody (NAb) titers were measured pre-vaccination (V0), and ∼28 days after the 2^nd^ (V2B) and 3^rd^ (V3B) doses. Anti-Spike/RBD IgG titers were also measured in a subset of participants (n=32) ∼6 days after the 1^st^ dose (V1). Anti-Spike/RBD IgG titers at V2B and V3B were positively correlated with each other and with NAb titers **(Fig. S1A-F)**. All participants had robust anti-RBD **(Fig. 1C)** and anti-Spike **(Fig. 1D)** IgG responses, with at least a 35-fold increase in anti-Spike IgG titers at V2B. Similar increases were observed in NAb titers **(Fig. 1E)**. We observed a significant but relatively weak negative correlation between age and anti-RBD/Spike and NAb titers at V2B **(Fig. S1G-I)** consistent with previous reports of age-dependent vaccine responses (Collier et al., 2021). No correlation with age was detected following the 3^rd^ dose **(Fig. S1J-L)**. Participants who received the ChAdOx1-S vaccine for their 1^st^ and 2^nd^ vaccinations had lower anti-Spike/RBD and NAb titers at V2B compared to those who received BNT162b2 **(Fig. 1C-E)**. After a 3^rd^ dose of either BTN162b2 or mRNA-1273, however, antibody titers were comparable in participants who received either ChAdOx1-S or BTN162b2 initially **(Fig. 1C-E)**.

Spike-specific T cell responses were assessed at V0, V2B and V3B by activation-induced marker (AIM) and ELISpot assays following stimulation of PBMCs with a peptide pool covering the full length of the Spike protein. AIM^+^ CD4^+^ and CD8^+^ T cell responses were correlated with each other, and with the ELISpot data **(Fig. S1M-O)**. A small population of Spike-reactive T cells was also present at baseline, which may be cross-reactive T cells derived from prior exposure to other endemic coronaviruses and/or environmental antigens (Bartolo et al., 2022, Mateus et al., 2021, Loyal et al., 2021). There was a positive correlation (*p* = 0.07) between the proportion of AIM^+^CD4^+^ T cells pre-vaccination and anti-RBD/Spike and NAb titers after the 2^nd^ dose (V2B). Vaccination with either BNT162b2 or ChAdOx1-S induced significantly increased antigen-specific T cell responses, relative to baseline, as measured by IFNγ secretion in ELISpot assays **(Fig. 1F)**. T cell responses in ChAdOx1-S vaccinated participants were significantly lower at V2B relative to BNT162b2 vaccinated participants, however, responses were not different after the 3^rd^ dose of BTN162b2 or mRNA-1273 **(Fig. 1F)**. AIM assays demonstrated a similar reduced frequency of AIM^+^CD4^+^ T cells after ChAdOx1-S at V2B, which recovered following a 3^rd^ vaccination **(Fig. 1G)**. There was no difference in AIM^+^CD8^+^ T cells in participants vaccinated with ChAdOx1-S or BNT162b2 at either V2B or V3B **(Fig. 1H)**. The majority of AIM^+^CD4^+^ T cells had an effector memory (Tem) surface phenotype **(Fig. S1S)**, whereas AIM^+^CD8^+^ T cells predominantly exhibited a Tem/TemRA surface phenotype **(Fig. S1T)**. The proportions of AIM^+^CD4^+^ or CD8^+^ T cells were not correlated with anti-RBD/Spike/NAb titers at V2B in participants vaccinated with BNT162b2. In contrast, following ChAdOx1-S, AIM^+^CD4^+^ T cells were positively correlated with NAb titers **(Fig. 1I)**, and the proportion of TemRA

AIM^+^CD4^+^ T cells was positively correlated with anti-Spike IgG titers **(Fig. 1J)**. Following the 3^rd^ dose of either BNT162b2 or mRNA-1273, CD8^+^ Tem cells were positively correlated with anti-Spike/RBD and NAb titers **(Fig. 1K)**, suggesting that individuals who mount robust CD8^+^ T cell responses also have a strong humoral response. The proportion of CD8^+^ TemRA cells, on the other hand, was negatively correlated with anti-RBD and NAb titers at V3B.

### Vaccination with the ChAdOx1-S but not BNT162b2 vaccine induces a memory-like cTfh and plasmablast response after the first dose

We performed total RNA sequencing to assess transcriptome-wide changes in peripheral blood at V0, V1, and ∼1-2 days after the 2^nd^ (V2A) and 3^rd^ (V3A) doses, generating more than 12 billion reads across 263 samples (mean 46M 2x150bp reads per sample, **Fig. S2A**, **Table S2)**. Consistent with a previous study reporting only a transient increase in the expression of type I interferon (IFN-I) inducible genes 1-2 days after the 1^st^ dose of BNT162b2 (Arunachalam et al., 2021), no differentially expressed genes (DEGs) were identified at V1 (mean 6 days post 1^st^ dose) in participants vaccinated with BNT162b2 **(Fig. 2A)**. Gene Set Enrichment Analysis (GSEA) did, however, detect an enrichment for interferon alpha inducible genes **(Table S2)**, and we observed a correlation between days p.v. and the expression of the gene, *IFI27* **(Fig. S2B**).

**Figure 2.**
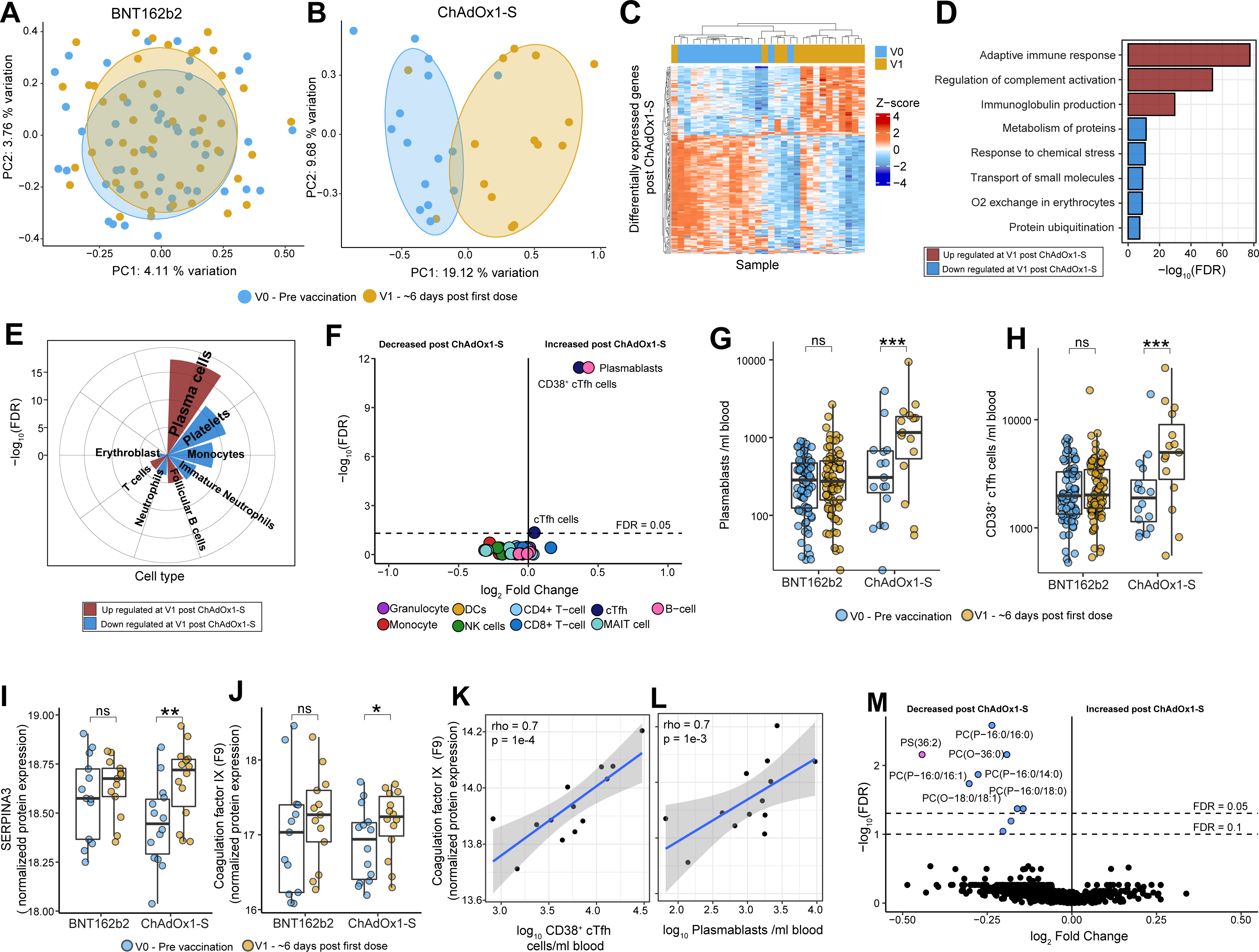
Multi-omics assessment of responses to a single dose of BNT162b2 or ChAdOx1-S at ∼6 days post-vaccination (V1). Multidimensional scaling analysis (MDS) of whole-blood gene expression profiles (RNAseq) at V0 and V1 after **(A)** BNT162b2 (n=66) or **(B)** ChAdOx1-S (n=15). **(C)** Heatmap of differentially expressed genes (DEGs) at V1 compared to V0 in participants vaccinated with ChAdOx1-S. **(D)** Selected Reactome pathways and Gene Ontology (GO) terms and **(E)** cell types, enriched among DEGs at V1 in participants vaccinated with ChAdOx1-S. **(F)** Volcano plot of immune cell populations after one dose of ChAdOx1-S. The number of **(G)** plasmablasts and **(H)** CD38^+^ cTfh cells at V1 in participants vaccinated with ChAdOx1-S (n=16) or BNT162b2 (n=77). **(I-J)** Normalized expression of selected proteins identified as differentially abundant (DA) in plasma at V1 compared to V0 in after ChAdOx1-S (n=14) or BNT162b2 (n=13). Correlation (Spearman) between coagulation factor IX protein expression at V1 and **(K)** CD38^+^ cTfh cells and **(L)** plasmablasts at V1 in participants vaccinated with ChAdOx1-S. **(M)** Volcano plot of DA lipid species at V1 compared to V0 in participants vaccinated with ChAdOx1-S. Data in G-J are represented as boxplots (See Fig. 1). Statistical significance was assessed in D and E using a hypergeometric test, in F-H, and M using a generalized linear model, and I-J with *limma*. *P < 0.05, **P < 0.01, ***P < 0.001, ns = not significant, FDR = false discovery rate.

In contrast, 424 DEGs were found at ∼6 days p.v. with ChAdOx1-S **(Fig. 2B-C**, **Table S2)**. Pathway enrichment analyses revealed that genes up-regulated at V1 after ChAdOx1-S were enriched for roles in the adaptive immune response, complement activation and immunoglobulin production **(Fig. 2D)**. Cell type enrichment analysis further suggested that up-regulated genes were strongly enriched for genes expressed in plasma cells **(Fig. 2E)**, particularly immunoglobulin genes **(Fig. S2C)**. In contrast, down-regulated genes were enriched for genes expressed in platelets and monocytes. To further investigate these signatures, comprehensive multi-parameter flow cytometry analysis was used to characterize changes in 54 different immune cell sub-populations **(Table S3**, **Supplementary File 2-3)**. Consistent with our RNASeq data, we did not detect any changes in immune cell populations at V1 following BNT162b2 **(Table S3)**. In contrast, vaccination with ChAdOx1-S had a striking effect on plasmablasts (CD19^+/dim^CD20^-/dim^CD38^++^CD27^++^) (Sanz et al., 2019) and circulating T-follicular helper cells (cTfh; CD3^+^CD4^+^CXCR5^+^PD-1^+^), in particular CD38^+^ cTfh cells (recently activated), which were increased following the 1^st^ dose of ChAdOx1-S **(Fig. 2F-H, Fig. S2D)**. Four ChAdOx1-S participants did not have increased plasmablasts or CD38^+^ cTfh cells **(Fig. 2G-H)**, however, these individuals had samples collected at 5 days p.v. **(Fig. S2E)**, suggesting that this plasmablast/cTfh response is only evident on or after day 6 p.v. PD-1 expression has been correlated with the activation of cTfh cells (Herati et al., 2017) and consistent with this, we observed increased PD-1 expression (MFI) on cTfh cells following the 1^st^ dose of ChAdOx1-S **(Fig. S2F)**.

The induction of plasmablasts and cTfh cells following the 1^st^ dose of the ChAdOx1-S is analogous to what is observed following influenza vaccination, where pre-existing immunity to influenza from previous infection or vaccination results in an increase in CD38^+^ cTfh cells and plasmablasts in the blood at ∼7 days p.v. (Herati et al., 2017, Bentebibel et al., 2013). The magnitude of plasmablast and cTfh response following influenza vaccination correlates with subsequent serum antibody titers (Bentebibel et al., 2013), however, we did not detect a correlation between the magnitude of the plasmablast or CD38^+^ cTfh cell response induced following the 1^st^ dose of ChAdOx1-S vaccine and antibody titers at V2B **(Fig. S2G-H)**, nor did we detect anti-Spike/RBD binding antibodies at V1 **(Fig. 1C-D)**. Taken together, these data suggest a memory response to a component of the ChAdOx1-S vaccine or a complex involving the vaccine (Baker et al., 2021). Vaccination with ChAdOx1-S has, in rare instances, been associated with TTS after the 1^st^ dose, a condition in which antibodies are generated that recognize platelet factor 4 (PF4) leading to activation of the coagulation cascade (Scully et al., 2021). We measured anti-PF4 IgG titers in serum collected from participants at V0 and at V1 following vaccination with ChAdOx1-S (n=14) and from an age- and sex-matched subset of BNT162b2 vaccinated participants (n=13), however, there was no statistically significant difference detected **(Fig. S2I)**.

### Plasma proteomics reveals a correlation between the expression of complement and coagulation related proteins and the memory-like cTfh and plasmablast response induced after the first dose of ChAdOx1-S

To further characterize the molecular changes following the 1^st^ dose of ChAdOx1-S, we performed untargeted proteomics on plasma samples (n=54) collected from the same participants for whom we measured anti-PF4 antibodies. Vaccination with ChAdOx1-S led to changes in the expression of 9 proteins in plasma at V1 **(Table S4)**, whereas no changes were detected following BNT162b2. Following ChAdOx1-S, for example, there was increased expression of SERPINA3, an acute phase protein **(Fig. 2I)**, complement component 9 (C9) **(Fig. 2J)**, and APOC2 **(Fig. S2J)**, a cofactor activating lipoprotein lipase. The expression of other proteins including the glycoprotein HRG **(Fig. S2K, Table S4),** were decreased following vaccination with ChAdOx1-S. Pathway enrichment analysis revealed complement activation related pathways were enriched among up-regulated proteins **(Table S4)**. The expression of multiple coagulation and complement related proteins including coagulation factors IX, XII, and Protein C, as well as complement component C3, C8A, C8B, and CFH, were strongly correlated with the number of CD38^+^ cTfh cells and plasmablasts induced following the 1^st^ dose of ChAdOx1-S **(Fig. 2K-L, Table S4)**. Taken together these data suggest that the magnitude of the memory response induced following the 1^st^ dose of the ChAdOx1-S vaccine is correlated with the expression of proteins involved in thrombosis.

We also performed untargeted lipidomics on the same set of 54 plasma samples. Consistent with our proteomics and RNASeq data, we found no changes in plasma lipid levels at V1 following BNT162b2, however, 9 lipid species were reduced following the 1^st^ dose of ChAdOx1-S **(Fig. 2M**, **Fig. S2L-O, Table S4)**. Lipids decreased following ChAdOx1-S were predominantly phosphatidylcholines, an abundant cellular phospholipid which are required for the generation of antibody-secreting cells (Brewer et al., 2016). 68 lipid species were correlated with the number of CD38^+^ cTfh cells and/or plasmablasts induced at V1 following ChAdOx1-S, including multiple negative correlations between the levels of phosphatidylcholine species and the number of plasmablasts **(Fig. S2P, Table S4)**, consistent with increased energy requirements for antibody production in ChAdOx1-S recipients at this timepoint.

### Similar changes to the blood transcriptome are induced ∼1-2 days after the second and third doses, irrespective of vaccine

Next, we assessed transcriptome-wide changes in the peripheral blood at V2A and V3A. Surprisingly, multidimensional scaling (MDS) analysis revealed that a 2^nd^ dose of either BNT162b2 or ChAdOx1-S induced remarkably similar changes to the transcriptome at V2A **(Fig. 3A)**. Furthermore, gene expression changes induced following a 3^rd^ dose of BNT162b2 or mRNA-1273 were indistinguishable from each other or from those changes induced after the 2^nd^ dose of BNT162b2 or ChAdOx1-S. Compared to pre-vaccination baseline, following BNT162b2, there were 611 and 1,015 DEGs (with a fold-change of at least 1.5 fold) at V2A and V3A, respectively **(Fig. 3B**, **Table S2)** including genes encoding transcription factors, such as *STAT1* and chemokines, such as *CXCL10* **(Fig. 3C-D)**. Similar numbers of DEGs were observed following vaccination with ChAdOx1-S at V2A (533 DEGs) or mRNA-1273 (1,549 DEGs) at V3A **(Table S2)**. Direct statistical comparison of gene expression responses after the 2^nd^ (BNT162b2 vs. ChAdOx1-S at V2A) or 3^rd^ dose (BNT162b2 vs. mRNA-1273 at V3A) did not identify any DEGs (FDR < 0.05), nor did comparing V2A to V3A, indicating that changes in gene expression induced at both timepoints were very similar, irrespective of the vaccine. Pathway analysis revealed that genes up-regulated at either V2A or V3A were strongly enriched for roles in interferon and cytokine signaling, antigen presentation, and the complement cascade **(Fig. 3E**, **Table S2)**.

**Figure 3.**
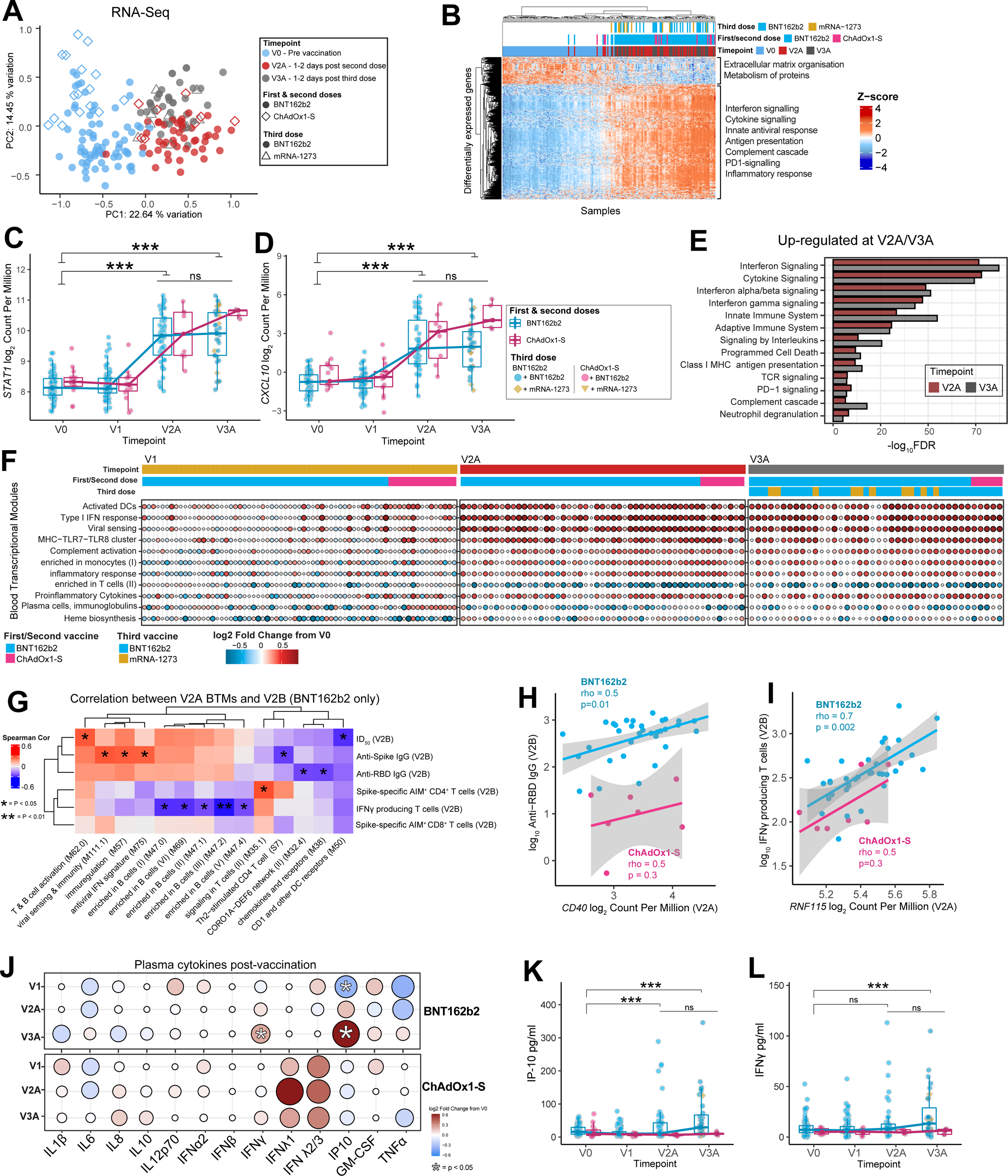
Gene expression and cytokine responses ∼1-2 days after the second and third doses of vaccine. (A) MDS of whole blood gene expression data at V0, V2A (BNT162b2 n=46, ChAdOx1-S n=8), and V3A (BNT162b2 n=32, mRNA-1273 n=10). **(B)** Heatmap of DEGs at V2A. Normalized gene expression of **(C)** *STAT1* and **(D)** *CXCL10* post-vaccination. **(E)** Selected Reactome pathways and GO terms enriched among DEGs at V2A/3A. **(F)** Selected blood transcriptional modules (BTMs) that were differentially expressed at V1, V2A or V3A post-vaccination. **(G)** Heatmap of Spearman correlations between BTM activity at V2A and antigen-specific antibody and T cell responses at V2B. Only BTMs with at least one statistically significant correlation (*p* < 0.05) are shown. Correlation (Spearman) between gene expression of **(H)** *CD40* at V2A and anti-RBD IgG titers at V2B and **(I)** *RNF115* at V2A and Spike-specific IFNγ SFU at V2B. **(J)** Fold-change in plasma cytokine levels at V1, V2A and V3A post-vaccination. The concentration of **(K)** IP-10 and **(L)** IFNγ in plasma pre-and post-vaccination. Data in C-D and K-L are represented as boxplots (See Fig. 1). Statistical significance was assessed in C-D using *edgeR*, in E using a hypergeometric test, and in J-L with a generalized linear model. *P < 0.05, ***P < 0.001, ns = not significant.

We next assessed transcriptional changes using Gene Set Variation Analysis (GSVA) (Hänzelmann et al., 2013). We used this approach to calculate an “activity score” for a set of >200 pre-defined blood transcriptional modules (BTMs) (Li et al., 2014), reflecting changes in expression of these BTMs in each participant at each timepoint p.v. **(Fig. 3F, Table S2)**. At V1, this approach detected the increase in plasma cell gene expression after ChAdOx1-S reported above, but also decreased activity of BTMs related to heme biosynthesis and platelet activation. Consistent with our pathway analysis, there was an increase in the activity of BTMs for antigen presentation, type I interferon response, and inflammation at V2A and V3A, irrespective of vaccine **(Fig. 3F, Table S2)**. No BTMs were identified as differentially active when comparing responses at V2A to V3A, further emphasizing the similarity in transcriptional responses after the 2^nd^ and 3^rd^ dose. Next, we assessed whether the activity of any BTMs at V2A or V3A were correlated with antigen-specific antibody or T cell responses at V2B or V3B (including anti-Spike/RBD/NAb titers, AIM^+^ CD4^+^/CD8^+^ cells, or the number of IFNγ-producing T cells). Several interesting and biologically plausible correlations were detected at *p* < 0.05 (**Fig. 3G & Fig. S3A)**, however, given the extremely large number of features measured across our multi-omics analyses, the majority of these correlations were not significant after correction for multiple testing. Correlations detected included a positive correlation between a T and B cell activation related BTM (M62) and NAb titers at V2B after BNT162b2 **(Fig. S3B)** and a positive correlation between an IFN related BTM (M75) and anti-Spike IgG titers (**Fig. S3C)**. Additionally, the “Signaling in T cells (II)” BTM (M35.1) was positively correlated with the proportion of AIM^+^CD4^+^ T cells at V2B **(Fig. S3D**). Interestingly, those BTMs that correlated with ChAdOx1-S immunogenicity were different to those identified for BNT162b2 and included several negative correlations between antigen presentation related BTMs and the proportion of AIM^+^CD4^+^ T cells at V2B **(Fig. S3A, Table S5)**. We also assessed correlations at the level of individuals genes, identifying hundreds of correlations before correction for multiple testing **(Table S5)**. For example, *CD40* expression in blood at V2A was positively correlated with anti-RBD titers at V2B **(Fig. 3H)**. Similarly, the expression of *RNF115* **(Fig. 3I)**, part of the RIG-I signaling pathway induced upon viral infection, was positively correlated with the number of IFNγ producing T cells at V2B. A further 15 BTMs at V3A were correlated with antibody titers or T-cell responses at V3B **(Fig. S3E, Table S5)**. Notably, there were several positive correlations between the activity of antigen presentation, monocyte, and DC related BTMs at V3A and the proportion of AIM^+^CD8^+^ T cells or IFNγ producing T cells at V3B, whereas plasma cell and memory B cell related modules were negatively correlated. Only 1 BTM (M58, B cell development/activation) was positively correlated with anti-RBD, anti-Spike and NAb titers.

Our transcriptional data indicated the increased gene expression of several cytokine and chemokine genes p.v. We next used a multiplex immunoassay (LEGENDplex™ Human Anti-Virus Response 13-plex Panel, BioLegend) to quantify 13 cytokines/chemokines in plasma at V0, V1, V2A and V3A (**Fig. 3J-L**). We did not detect a difference in cytokine levels at V1 in participants vaccinated with either BNT162b2 or ChAdOx1-S. Consistent with previous reports (Bergamaschi et al., 2021, Takano et al., 2022) and our gene expression data, plasma IP-10 levels were elevated in BNT162b2 recipients ∼1-2 days after the 2^nd^ and 3^rd^ doses (**Fig. 3K**). IFNγ levels were also significantly increased after the 3^rd^ dose (**Fig. 3L**). Although no cytokines/chemokines were significantly elevated following ChAdOx1-S (**Fig. 3J-L**), this could be due to the smaller sample size compared to the BNT162b2 group.

### Lymphopenia and activation of immune cells ∼1-2 days after the second and third doses of vaccine

As reported above, we performed flow cytometry analysis to characterize changes in 54 immune cell sub-populations at V0 and all timepoints p.v. with BNT162b2 induced a mild lymphopenia at V2A and V3A **(Fig. 4A-B, Fig. S5A, Table S3)**, which included a reduction in CD3^+^ T cells (**Fig. 4C**), multiple NK cell subsets (**Fig. 4A-B, D**), both conventional (cDC) and plasmacytoid DCs (pDC), multiple subsets of CD4^+^ and CD8^+^ T cells, cTfh cells and memory B cell subsets (**Fig. 4A-B, Table S3**). Intermediate monocytes were increased at V2A and V3A after BNT162b2 (**Fig. 4E**). No populations were detected as altered following ChAdOx1-S compared to V0, however, as can be observed **(Fig. 4B),** the trend was similar to BNT162b2 and the lack of statistical significance is likely due to the smaller sample size of ChAdOx1-S participants. When directly statistically comparing immune cell populations following ChAdOx1-S and BNT162b2, aside from the differences at V1 reported above, only classical monocytes were significantly different, and were increased at V2A after ChAdOx1-S (**Fig. 4F)**. No differences in immune cell populations were detected at V3A or V3B when comparing between the different vaccine groups.

**Figure 4.**
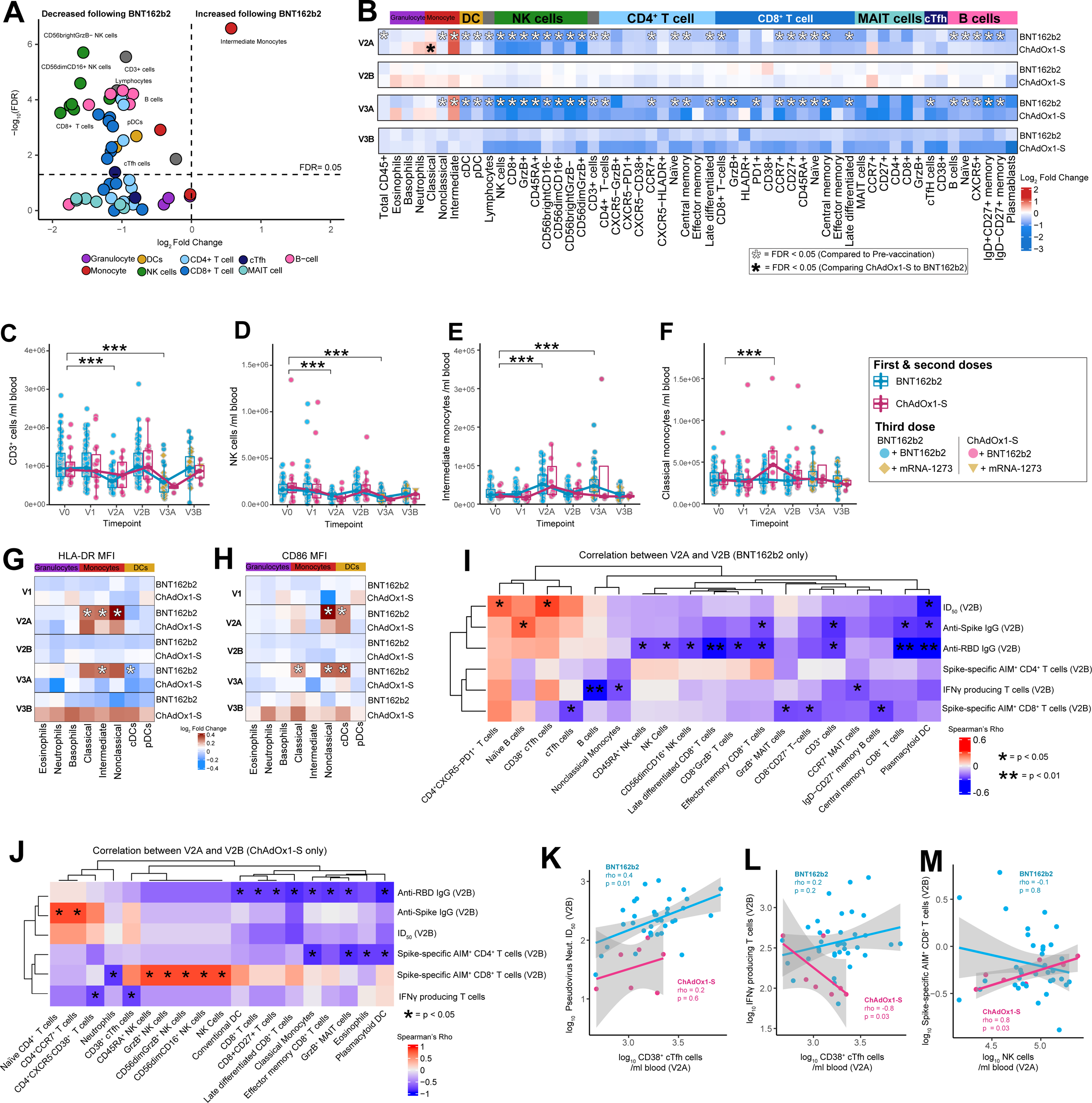
Flow cytometry analysis of major immune cell populations in peripheral blood ∼1-2 days after the second and third doses of vaccine. (A) Volcano plot showing changes in immune cell populations in BNT162b2 vaccinated participants (n=44) at V2A relative to V0. **(B)** Heatmap showing the fold-change (log_2_ transformed) in immune cell populations in BNT162b2 or ChAdOx1-S vaccinated participants at V2A, V2B, V3A and V3B relative to V0. **(C)** CD3^+^ cells, **(D)** NK cells, **(E)** intermediate monocytes, and **(F)** classical monocytes (counts/ml) in peripheral blood pre- and post-vaccination. Heatmap showing fold-change in the expression of myeloid cell activation markers **(G)** HLA-DR and (**H**) CD86 at V1-V3B relative to V0. Heatmaps of correlations between immune cell populations at V2A and antigen-specific antibody and T cell responses at V2B following **(I)** BNT162b2 or **(J)** ChAdOx1-S. Only populations with at least one significant correlation (*p* < 0.05) shown. Correlation between the number of CD38^+^ cTfh cells at V2A with **(K)** ID_50_ and **(L)** Spike-specific IFNγ SFU, at V2B. **(M)** Correlation between NK cells at V2A with Spike-specific AIM^+^CD8^+^ T cells at V2B. Statistical significance was assessed in A-H with a generalized linear model. Spearman correlations shown in I-M. *P < 0.05, ***P < 0.001.

We also examined myeloid activation markers and found that after 2 doses of BNT162b2, the expression of HLA-DR was increased on all monocyte subsets assessed (**Fig. 4G**). ChAdOx1-S recipients showed a similar trend, but again these differences did not reach statistical significance. HLA-DR expression was similarly increased on monocyte subsets at V3A after BNT162b2, although these differences were only statistically significant for intermediate monocytes. Expression of CD86 was increased on nonclassical monocytes and cDCs at V2A and V3A, and on classical monocytes at V3A, in participants vaccinated with BNT162b2 (**Fig. 4H**).

The frequencies of multiple immune cell populations at V2A were correlated with antigen-specific antibody or T cell responses at V2B **(Fig. 4I-J**). For example, the number of pDCs, which decreased after vaccination, was negatively correlated with antibody and T cell responses at V2B, while CD38^+^ cTfh cells and PD-1 expressing CD4^+^ T cells were positively correlated with NAb titers (**Fig. 4 I**). The correlations observed following ChAdOx1-S were different to those observed following BTN162b2 **(Fig. 4J)**. For example, multiple CD8^+^ T cell and innate immune cell subsets were negatively correlated with anti-RBD titers at V2B, while multiple NK cell subsets were positively correlated with the proportion of AIM^+^CD8^+^ T cells at V2B (**Fig. 4J**). Following BNT162b2, the number of CD38^+^ cTfh cells at V2A was positively correlated with NAbs at V2B (**Fig. 4K**). In some cases, opposing correlations were observed. For example, CD38^+^ cTfh cells at V2A were negatively correlated with Spike-specific IFNγ-secreting cells at V2B following ChAdOx1-S; but were positively correlated following BNT162b2 (**Fig. 4L**). Similarly, the frequency of NK cells at V2A were positively correlated with AIM^+^CD8^+^ T cells at V2B after ChAdOx1-S, but negatively correlated after BNT162b2 (**Fig. 4M**). No immune cell populations at V3A were correlated with antibody responses at V3B, however 10 immune cell populations were negatively correlated with T-cell responses (**Table S5**), including negative correlations between naïve CD4^+^ T cells at V3A and the number of Spike-specific IFNγ-secreting cells at V3B, and between CD4^+^ MAIT cells at V3A and the proportion of AIM^+^CD8^+^ T cells at V3B (**Table S5**).

### Pre-vaccination immune status is correlated with vaccine immunogenicity

As previous studies suggest the pre-vaccination immune state is predictive of antigen-specific responses to other vaccines (Tsang et al., 2014), we sought to assess whether this is also true for COVID-19 mRNA or adenovirus vectored vaccines. We found multiple immune cell populations, BTMs and cytokine and chemokine levels pre-vaccination were correlated with antigen-specific antibody or T-cell responses at V2B following BNT162b2 (**Fig. 5A**) or ChAdOx1-S (**Fig. S5B**). For example, the number of CD38^+^ cTfh cells pre-vaccination was positively correlated with NAb titers at V2B following vaccination with BNT162b2 (**Fig. 5B**). Unexpectedly, we found that both the transcriptional activity of B cell-related BTMs pre-vaccination and number of B cells in blood, as assessed by flow cytometry, were negatively correlated with antigen-specific T-cell responses at V2A p.v. with either BNT162b2 or ChAdOx1-S **(Fig. 5A, C-D, Fig. S5B, Table S5**). We also identified that the pre-vaccination concentration of IP-10 was positively correlated with anti-Spike IgG titers following BNT162b2 **(Fig. 5E)**. We also identified correlations between BTMs, immune cell populations, and cytokine concentrations with antibody and T cell responses induced following the 3^rd^ dose (either BNT162b2 or mRNA-1273) (**Fig. 5F, Table S5**). For example, the number of naïve B cells pre-vaccination was correlated with anti-Spike titers at V3B, whereas BTM M146 (MHC−TLR7−TLR8 cluster) was positively correlated with the proportion of AIM^+^ CD8^+^ T cells at V3B (**Fig. 5F**).

**Figure 5.**
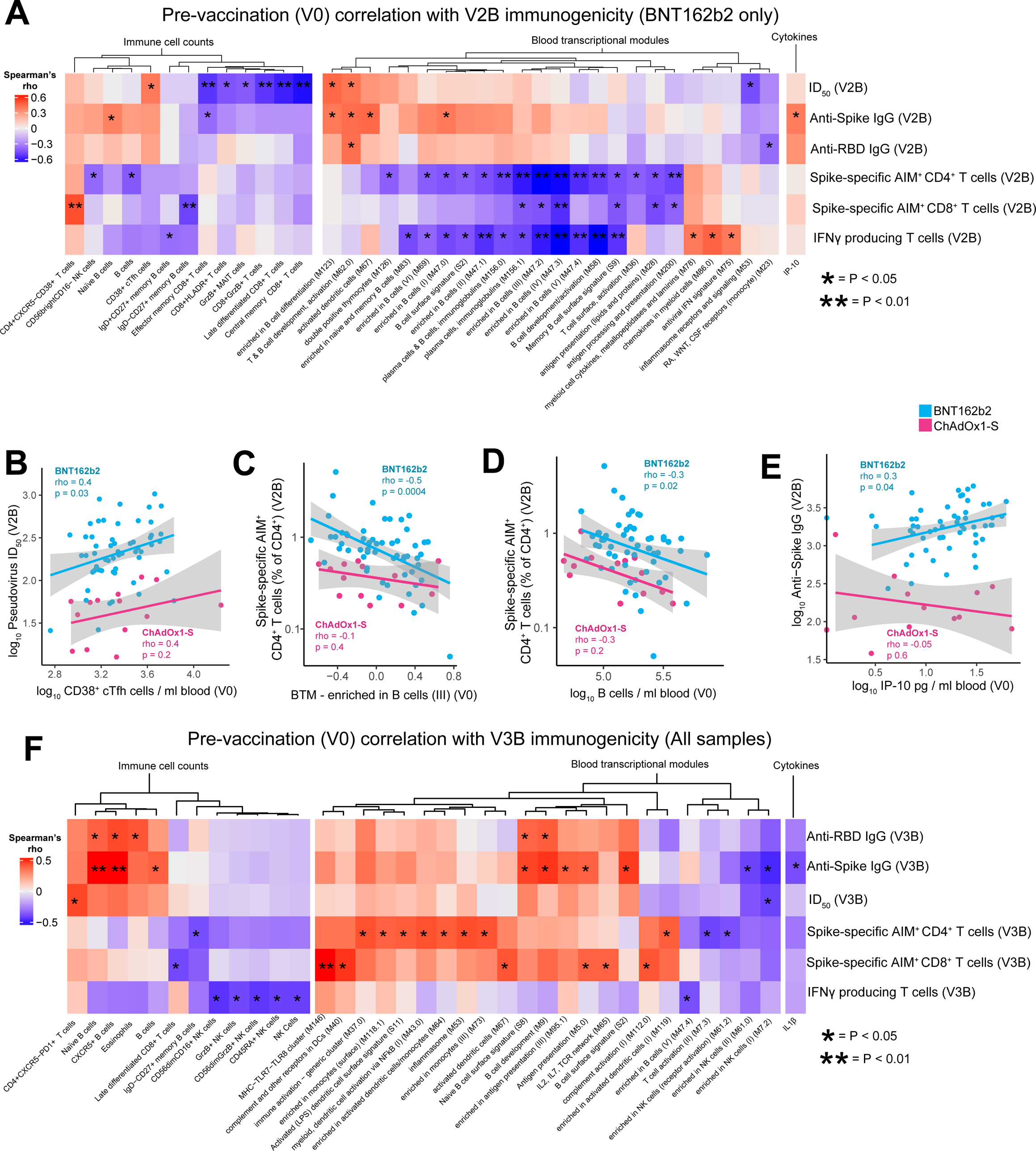
Pre-vaccination immune status is correlated with antigen-specific antibody and T cell responses post the second or third doses of vaccine. (A) Heatmap showing correlations between BTM activity score, immune cell counts or cytokine concentration at V0, and antigen-specific antibody and T cell responses at V2B. **(B)** Correlation between CD38^+^ cTfh cells at V0 and ID_50_ at V2B. **(C)** Correlation between activity of BTM 47.2 (enriched in B cells III) at V0 and Spike-specific AIM^+^CD4^+^ T cells at V2B. **(D)** Correlation between B cells (counts/ml) at V0 and Spike-specific AIM^+^CD4^+^ T cells at V2B. **(E)** Correlation between IP-10 in plasma at V0 and anti-Spike IgG titers at V2B. **(F)** Heatmap of correlations between BTM activity, immune cell counts or cytokine concentration at V0 and antigen-specific responses at V3B. (**A** and **F**) Only BTMs, immune cell populations, or cytokines with at least one statistically significant correlation (*p* < 0.05) are shown. Spearman correlations shown in A-F. *P < 0.05, **P < 0.01.

### Reactogenicity post-vaccination is associated with higher T cell responses but not antibody responses

A survey recording symptoms of vaccine reactogenicity was sent to participants one week after each vaccination (>95% completion rate, **Table S6**). No participants reported serious AEs (e.g. TTS). Following the 1^st^ dose, participants who received ChAdOx1-S reported higher rates of fatigue, fever, chills, muscle pain and headache compared to those vaccinated with BNT162b2 (**Fig. 6A**). In contrast, following the 2^nd^ dose of vaccine, higher rates of chills were detected in those vaccinated with BNT162b2 (**Fig. 6B**). There was no significant difference in reported AEs between those vaccinated with BNT162b2 or mRNA-1273 for their 3^rd^ dose (**Fig. 6C**). These patterns of AEs were consistent with prior reports (Therapeutic Goods Administration, 2022). In contrast to prior reports (Takano et al., 2022), we did not detect associations between reported AEs and antigen specific anti-Spike/RBD of NAb titers induced by either vaccine at any timepoint (data not shown). However, higher frequencies of HLA-DR^+^CD8^+^ T cells, PD-1^+^CD8^+^ T cells, and CCR7^+^CD8^+^ T cells were observed in those who experienced fatigue, headache, or fever, respectively, after the 1^st^ dose of ChAdOx1-S (**Fig. 6D-F**). HLA-DR^+^ CD8^+^ T cells were also increased in those who experienced a headache after the 1^st^ dose of BNT162b2 (**Fig. 6G**). The frequency of plasmablasts at V1 was also associated with pain at the injection site and headache following the 1^st^ dose of BNT162b2 (**Fig. 6H-I**) but was not correlated with subsequent antibody or T cell responses. Participants reporting chills at V2A following vaccination with BNT162b2 had lower levels of IL8, IP-10 and TNFα (**Fig. 6J-L)**. Interestingly, those who reported fever following the 2^nd^ dose of BNT162b2 had increased numbers of Spike-specific IFNγ-secreting cells at V2B (**Fig. 6M**) and an increased proportion of Spike-specific AIM^+^CD4^+^ T cells **(Fig. 6N)**. Following the 3^rd^ dose (BNT162b2 or mRNA-1273), we observed that fatigue was associated with an increased proportion of Spike-specific IFNγ-secreting cells and AIM^+^ CD4^+^ and AIM^+^ CD8^+^ T cells (**Fig. 6O-Q**). These data suggest a relationship between reactogenicity and increased T cell responses following vaccination with COVID-19 mRNA vaccines.

**Figure 6.**
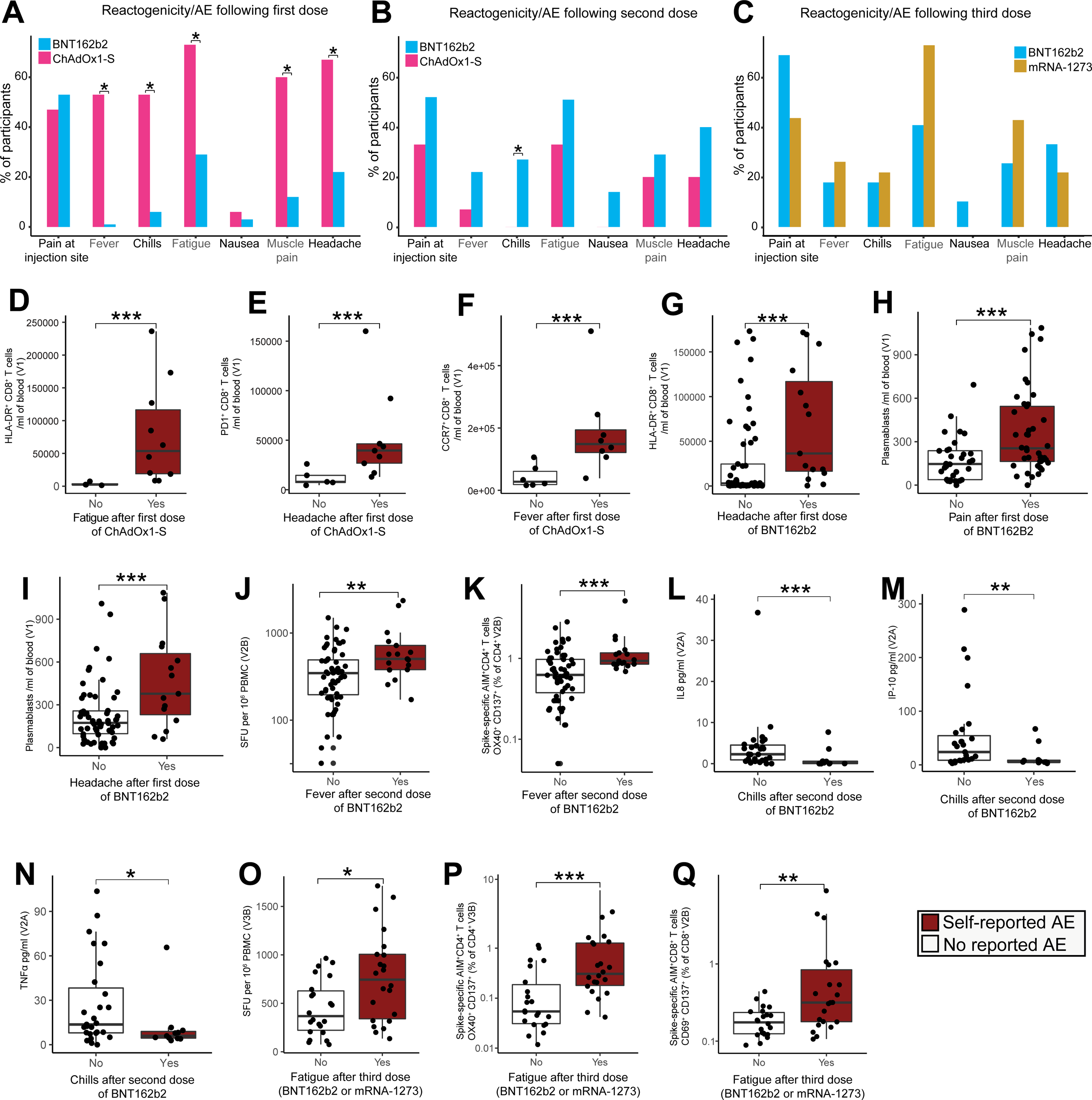
Reactogenicity following the BNT162b2, mRNA-1273 or ChAdOx1-S vaccines. The proportion of participants who reported adverse events (AE) following the **(A)** 1^st^, **(B)** 2^nd^, or **(C)** 3^rd^ dose of vaccine. Activated CD8^+^ T cells subsets following the 1^st^ dose of ChAdOx1-S in participants who reported **(D)** fatigue, **(E)** headache or **(F)** fever. **(G)** HLA-DR^+^CD8^+^ T cells in participants who reported headache after the 1^st^ dose of BNT162b2. **(H)** Plasmablasts in participants who reported pain at the injection site after the 1^st^ dose of BNT162b2. **(I)** Plasmablasts in participants who reported headache after the 1^st^dose of BNT162b2. **(J)** Spike-specific IFNγ SFU at V2B in participants who reported fever after a 2^nd^dose of BNT162b2. **(K)** AIM^+^ CD4^+^ T cells at V2B in participants who reported fever after a 2^nd^ dose of BTN162b2. Concentration of **(L)** IL8, **(M)** IP-10, and **(N)** TNFα at V2A in participants who reported chills after the 2^nd^ dose of BNT162b2. **(O)** Spike-specific IFNγ SFU at V3B in participants who reported fatigue after a 3^rd^ dose (BNT162b2/mRNA-1273). **(P)** AIM^+^CD4^+^ T cells at V3B in participants who reported fatigue after a 3^rd^dose of BNT162b2 or mRNA-1273. **(Q)** The proportion of Spike-specific AIM^+^CD8^+^ T cells at V3B in participants who reported fatigue after a 3^rd^ dose of BNT162b2 or mRNA-1273. Data in D-Q are represented as boxplots (see Fig. 1). Statistical significance was assessed in A-C using Fisher’s Exact tests, and in D-Q using Wilcoxon Rank Sum tests. *P < 0.05, **P < 0.01, ***P < 0.001.

## Discussion

Vaccination is a cornerstone of the global strategy to control the spread of SARS-CoV-2 with tens of millions of lives saved to date (Watson et al., 2022). While randomized controlled trials have demonstrated the efficacy of both mRNA and adenoviral vectored vaccines against severe COVID-19 (Folegatti et al., 2020, Baden et al., 2021, Polack et al., 2020, Voysey et al., 2021), such trials typically do not include in-depth, systems-level, assessments of the immune response to vaccination, limiting our knowledge of the precise mechanisms which determine optimal antigen-specific immune responses to different COVID-19 vaccine types. Here, we present a longitudinal, systems-level, analysis of the early (days) and late (weeks-months) immune responses induced in a cohort of 102 SARS-CoV-2 naïve, healthy adults, vaccinated with either mRNA (BNT162b2, mRNA-1273) or adenoviral vectored (ChAdOx1-S) vaccines. We comprehensively profiled immune responses in blood before vaccination, at 1-7 days following the first, second and third doses, using a multi-omics approach that included transcriptomics, proteomics, lipidomics, cytokine profiling and multi-parameter flow cytometry, and compared these responses to antigen-specific antibody and T cell responses induced ∼28 days after the second and third doses. Our multi-omics approach has identified previously unreported differences in the immune response to different COVID-19 vaccines and identified elements of the pre-vaccination immune status, as well as innate responses induced 1-2 days post-vaccination, which correlate with antigen-specific antibody and T cell responses.

Following the first dose of ChAdOx1-S, we detected significant differences in the blood transcriptome that were not observed in BNT162b2 vaccinated participants. Analysis of these transcriptional changes suggested the induction of a plasma cell response within 6 days of the first dose of ChAdOx1-S. Multiparametric flow cytometry confirmed this, revealing a significant increase in both plasmablasts and recently activated cTfh cells in the blood of ChAdOx1-S vaccinated individuals 6-7 days after the first dose. This response is remarkably similar to the anamnestic responses observed following seasonal influenza vaccination (Bentebibel et al., 2013, Koutsakos et al., 2018) and suggests pre-existing immunity to a component of the ChAdOx1-S vaccine or vector. Pre-existing immunity in the form of ChAdOx1 neutralizing antibodies is thought to be uncommon (Dicks et al., 2012, Ramasamy et al., 2020), but there has been little investigation into ChAdOx1 cross-reactive T cell memory and the consequences this could have. ChAdOx1-S vaccination has recently been shown to expand a pre-existing CD4^+^ memory T cell pool that also responds to the human adenovirus 5 (Ad5) hexon protein, which shares ∼80% sequence homology with the ChAdOx1 hexon protein (Saggau et al., 2022), strongly suggesting that cross-reactive T cell epitopes are shared between human adenoviruses and the ChAdOx1 viral vector. It is possible then that a proportion of the expanding cTfh cells we observe 6-7 days post-vaccination with ChAdOx1-S are human adenovirus-specific memory cells that cross-react with a component of ChAdOx1. The specificity of the plasmablasts we observe following ChAdOx1-S vaccination, however, remains unclear, as they were not associated with the production of Spike-specific antibodies after the first dose or at any other timepoint, nor with anti-PF4 antibody levels.

Interestingly, our proteomics analyses revealed that there was a strong positive correlation between the abundance of multiple coagulation and complement proteins in plasma and the magnitude of the cTfh and plasmablast response induced 6-7 days after the first dose of ChAdOx1-S, suggesting a link between the induction of this memory-like response and the expression of proteins involved in thrombosis. Adenoviral vectored vaccines have been associated with rare cases of TTS, driven in part by the production of auto-antibodies against PF4, with clinical symptoms manifesting from 7 days post-vaccination (Greinacher et al., 2021, Schultz et al., 2021, Scully et al., 2021). How these PF4 auto-antibodies manifest in those rare cases of TTS is unclear, but it has been demonstrated that several adenovirus vaccine vector candidates, including ChAdOx1, bind to PF4 *in vitro* (Baker et al., 2021, Michalik et al., 2022). If the same phenomenon occurs *in vivo,* PF4-specific B cells could internalize and present PF4-ChAdOx1 complexes to ChAdOx1 cross-reactive memory T cells, leading to intermolecular epitope spreading and the production of PF4 auto-antibodies (Vanderlugt and Miller, 2002, Cornaby et al., 2015), consistent with a proposed mechanism for TTS (Baker et al., 2021). Although we were unable to detect a statistically significant increase in PF4 auto-antibodies in ChAdOx1-S vaccinated participants, none of whom developed TTS, further investigation is warranted to assess if the cTfh cells and plasmablasts that appear following the first dose of adenovirus-vectored vaccines are specific for the vector, or the antigen expressed, and the consequences this may have for vaccine-induced AEs, such as TTS.

Interestingly, given the differences observed after the first dose, similar transcriptional, cytokine/chemokine and immune cell population changes were induced ∼1-2 days following the second or third dose, irrespective of vaccine. Differential gene expression analyses identified several hundred genes changed in response to vaccination, and through correlation analyses we were able to link the expression of specific genes (e.g., *CD40*) and BTMs, induced ∼1-2 days post-vaccination, to the magnitude of subsequent antigen-specific antibody and T cell responses assessed ∼28 days later. A mild lymphopenia was induced ∼1-2 days after the second and third vaccine dose, which was observed irrespective of vaccine administered. This lymphopenia was primarily driven by reduced numbers of T, B and NK cells, likely due to migration from the peripheral blood to lymphoid organs. The magnitude of the mild lymphopenia was also correlated with antigen-specific responses ∼28 days later. Interestingly, the only cell populations detected as increased in this period following vaccination were classical (CD14^+^CD16^-^) and intermediate (CD14^+^CD16^+^) monocytes, that were increased by ChAdOx1-S or BNT162b2 vaccination, respectively. The increase in classical monocytes is particularly noteworthy as they were negatively correlated with anti-RBD antibody responses induced by ChAdOx1-S. In other contexts, monocyte recruitment has been shown to be capable of disrupting formation of germinal centers (Biram et al., 2022, Sammicheli et al., 2016). We also identified a positive correlation between the number of CD38^+^ cTfh cells at V2A and pseudovirus neutralizing antibody titers at V2B in BNT162b2 vaccinated, but not ChAdOx1-S participants. Additionally, the frequency of specific immune cell populations and the transcriptional activation of BTMs pre-vaccination were correlated with subsequent vaccine-specific B and T cell responses. Irrespective of vaccine, an overt B cell signature pre-vaccination was associated with lower vaccine-specific T cell responses at V2B, while abundant cytotoxic CD8^+^ T cell subsets pre-BNT162b2 vaccination was associated with lower NAb titers at V2B. These data suggest that pre-vaccine immune signatures can be used to predict the magnitude of subsequent vaccine-specific immune responses.

Consistent with prior reports (Zhang et al., 2022, Khoo et al., 2022, Tarke et al., 2022), the proportion of AIM^+^CD4^+^ T cells was lower in participants that received ChAdOx1-S relative to BNT162b2 vaccinated participants. However, following a third dose of a mRNA vaccine, AIM^+^CD4^+^ T cells in ChAdOx1-S vaccinated participants recovered to levels comparable to triple mRNA vaccinated participants (Khoo et al., 2022, Bánki et al., 2022). The proportion of AIM^+^CD4^+^ T cells at V2B in ChAdOx1-S vaccinated participants was also positively correlated with NAb titers, which were reduced in ChAdOx1-S vaccinated participants relative to BNT162b2 participants at V2B, but recovered after a heterologous mRNA vaccination. These data suggest that a possible underlying cause for the lower antibody titers following ChAdOx1-S vaccination may be suboptimal CD4^+^ T cell responses, which provide less help to promote humoral responses. In support of this, the number of IFNγ-secreting cells at V2B in ChAdOx1-S vaccinated participants was negatively correlated with the number of cTfh cells as determined by flow cytometry. As Th1 and Tfh cells represent mutually exclusive T cell fates (Lönnberg et al., 2017, Nakayamada et al., 2011), ChAdOx1-S vaccination may drive a Th1 biased response at the expense of Tfh differentiation, resulting in poorer antibody responses as compared to the BNT162b2 vaccine.

Whether vaccine reactogenicity translates into enhanced immunogenicity is currently not well understood, with conflicting reports (Röltgen et al., 2022, Lee et al., 2021). Here, reactogenicity following the first dose of ChAdOx1-S was associated with increased activation of CD8^+^ T cells, and increased numbers of circulating plasmablasts in the blood of BNT162b2 vaccinated individuals. Interestingly, we did not detect any significant associations between reactogenicity and anti-Spike/RBD or NAb titers. However, fever and fatigue were associated with more robust AIM^+^ T cell responses following the second and third dose of vaccine, respectively. We also identified a significant association between reactogenicity and CD8^+^ T cell activation which, in turn, was negatively correlated with anti-Spike IgG titers. Together these data suggest that reactogenicity is associated with enhanced vaccine-specific T cell responses, but not vaccine-specific antibody titers.

In conclusion, COVIRS represents a significant resource for understanding the early innate and adaptive immune responses to mRNA and adenovirus vectored COVID-19 vaccines, and how these responses relate to immunogenicity and reactogenicity. The generation of high titers of Spike-specific binding and neutralizing antibodies is currently the best correlate of protection following vaccination against SARS-CoV-2, although the contribution of vaccine-induced T cells in protection from SARS-CoV-2 infection is becoming increasingly apparent (McMahan et al., 2021, Liu et al., 2022). Identifying the factors that promote the optimal generation of vaccine-specific T and B cell responses is a key priority and our study provides a wealth of data to inform future rational vaccine design.

### Limitations

Our study has several limitations, including an imbalance of in the number of participants receiving BNT162b2 compared to the ChAdOx1-S and mRNA-1273 vaccines. This was unavoidable as BNT162b2 was the recommended vaccine for the majority of participants in our cohort and vaccine administration was not under our control. The cost and complexity of the multi-omics analyses performed also limits the number of participants that can be included in such systems vaccinology studies, though we note that our sample size is at the upper end of comparable studies in the field. Finally, analyses were performed on peripheral blood; analysis of immune responses in draining lymph nodes would be an exciting addition in future studies.

## Supporting information

STROBE Checklist

## Data Availability

Multi-omics datasets and R code are available via the Lynn Laboratory BitBucket (https://bitbucket.org/lynnlab/covirs). RNASeq data have been deposited in the Gene Expression Omnibus (GEO) under accession GSE199750. The mass spectrometry proteomics data have been deposited to the ProteomeXchange Consortium via the PRIDE partner repository with the dataset identifier PXD036608

## Acknowledgements

We would like to thank all of the study participants. This work was funded by BioPlatforms Australia, Flinders Foundation and EMBL Australia Group Leader funding awarded to DJL. The proteomics component of this study was funded by AstraZeneca. This work was also supported in part by a MRFF Coronavirus research response grant (APP2015305) and The Hospital Research Foundation Group COVID-19-SA grant. We thank SA Pathology for help with sample collection and the South Australian Genomics Centre (SAGC) for help with RNA sequencing. The SAGC is supported by the National Collaborative Research Infrastructure Strategy (NCRIS) via BioPlatforms Australia and by the SAGC partner institutes. We thank the Garvan Institute for providing the sequin controls for RNA sequencing. The MR1 tetramer technology was developed jointly by Drs. McCluskey, Rossjohn, and Fairlie, and the material was produced by the NIH Tetramer Core Facility as permitted to be distributed by the University of Melbourne. Flow cytometry analysis was performed at the ACRF Cellular Imaging and Cytometry Core Facility in SAHMRI. The ACRF Facility is generously supported by the Detmold Hoopman Group, Australian Cancer Research Foundation, and Australian Government through the Zero Childhood Cancer Program.

## Author contributions

The study was conceived and designed by D.J.L. F.J.R. performed the multi-omics analyzes under the supervision of D.J.L with input from S.J.B., T.S.N., C.M., and N.E.S.; S.J.B. and N.E.S. generated the flow cytometry data with help from S.C.B.; N.E.S. generated the cytokine data. T.S.N. conducted the T cell ELISpot and AIM assays. J.J, G.L.E., Y.C.T., and M.A.L managed and processed the clinical samples. C.M., V.I. J.Z. and P.M. generated the proteomics data. H.S., H.T. and J.D.M. performed the lipidomics analysis. M.G.M., A.E.L.Y. and B.G-B. generated the binding antibody data. A.A., A.D and R.B. generated the neutralizing antibody data. Sample collection protocols were adapted from protocols originally developed by N.L.M. and N.C. Participant surveys were developed by N.L.M and N.C.; R.B. and S.E.B. were responsible for participant coordination. The manuscript was written by F.J.R., T.S.N., N.E.S, S.J.B and D.J.L with contributions and approval from all the authors.

## Supplementary Data

**Supplementary File 1:** Reactogenicity survey sent to participants.

**Supplementary File 2:** Flow cytometry gating strategy.

**Supplementary File 3:** Plots of all immune cell populations measured in COVIRS as both counts and frequencies over all timepoints.

**Figure S1: (A-F)** Correlations between anti-RBD, anti-Spike and pseudovirus neutralizing antibody titers at V2B and V3B. Correlations between anti-RBD, anti-Spike and pseudovirus neutralizing antibody titers at **(G-I)** V2B, and V3B **(J-L)**, with participant age. Correlations between Spike-specific T-cell responses **(M-R)** (the number of Spike-specific IFNγ SFU, the proportion of Spike-specific AIM^+^CD4^+^ T cells, the proportion of Spike-specific AIM^+^CD8^+^ T cells) at V2B and V3B. Bar plots showing the composition of the AIM^+^ **(S)** CD4^+^ and **(T)** CD8^+^ T cell compartments. Spearman correlations shown in A-R.

**Figure S2: (A)** MDS analysis of whole-blood gene expression profiles (RNAseq) in participants pre-vaccination (V0), ∼6 days after a 1^st^ dose of BNT162b2 (n=66) or ChAdOx1-S (n=15), ∼1-2 days after a 2^nd^ dose (V2A) of BNT162b2 (n=46) or ChAdOx1-S (n=8), and ∼1-2 days after a 3^rd^ dose (V3A) of BNT162b2 (n=32) or mRNA-1273 (n=10). (**B)** Normalized gene expression of *IFI27* in blood of BNT162b2 vaccinated participants plotted by day of sample collection post the 1^st^ dose. (**C)** Heatmap showing the expression of immunoglobulin genes identified as differentially expressed (DE) between V0 and V1. Data were adjusted for sex and batch effects prior to MDS analysis and visualization of the heatmap. (**D**) Representative dot plot of CCR7 and CD38 expression on cTfh (CD3^+^CD4^+^CXCR5^+^PD1^+^) and plasmablasts (CD19^+/dim^CD20^-/dim^CD38^++^CD27^++^) at V0 and V1 after 1^st^ dose of BNT162b2 or ChAdOx1-S. **(E)** CD38^+^ cTfh cells plotted by day of sample collection for ChAdOx-1 participants. (**F)** Fold-change in the mean fluorescence intensity (MFI) of PD-1 expression on cTfh cells at V1 compared to V0. Correlation (Spearman) between the number of **(G)** CD38^+^ cTfh cells or **(H)** plasmablasts at V1 and ID_50_ at V2B. (**I)** Fold-change in anti-PF4 optical density (OD) at V1 compared to V0. Normalized protein abundance in plasma of **(J)** APOC2 and **(K)** HRG. vsn = Variance stabilizing normalization. See Table S4 for complete list of differentially abundant proteins. **(L-O)** Selected lipid species identified as differentially abundant at V1 in participants vaccinated with ChAdOx1-S. See Table S4 for complete list. **(P)** Correlation between plasmablasts at V1 and the level of phosphocholine (O-18:0/18:1) in plasma. See Table S4 for all significant correlations. Statistical significant assessed in F and I with a Wilcox rank sum test, in J-O with a generalized linear model. *P < 0.05, ***P < 0.001

**Figure S3: (A)** Heatmap of Spearman correlations between BTM activity at V2A and antigen-specific antibody and T cell responses at V2B for participants who received ChAdOx1-S. Only BTMs with at least one statistically significant correlation (*p* < 0.05) are shown. Correlation between the activity of **(B)** T and B cell activation BTM (M62.0) with ID_50_ at V2B, **(C)** antiviral IFN signature BTM (M75) and anti-Spike IgG at V2B and **(D)** signaling in T cells BTM (M35.1) with Spike-specific AIM^+^ CD4^+^ T cells at V2B. **(E)** Heatmap of correlations between BTM activity at V3A and antigen-specific antibody and T cell responses at V3B (All participants). Only BTMs with at least one statistically significant correlation (*p* < 0.05) are shown. *P < 0.05, **P < 0.01

**Figure S4:** Cytokine concentrations in plasma pre-vaccination (V0) and at V1, V2A and V3A post-vaccination with BNT162b2 or ChAdOx1-S. Data are represented as Tukey style boxplots. Statistical significance in A-K was assessed using a generalized linear model. No statistically significant (*p* < 0.05) differences were detected for these cytokines.

**Figure S5: A)** Volcano plot of immune cell counts at V3A relative to V0. **B) (A)** Heatmap showing Spearman correlations between BTM activity score, counts of immune cells or cytokine concentration, at V0 and antigen-specific antibody or T cell responses at V2B following 2 doses of ChAdOx1-S.

**Table S1.** COVIRS sampling and demographic summary.

**Table S2.** RNAseq data, differentially expressed genes at each timepoint, pathway and blood transcriptional module (BTM) analysis.

**Table S3.** Flow cytometry results.

**Table S4.** Proteomics and lipidomics results.

**Table S5.** Correlation between multi-omics data pre- and post-vaccination with antigen-specific antibody and T cell responses at V2B and V3B.

**Table S6.** Summary of self-reported reactogenicity following each vaccine dose.

## Methods

### Participant recruitment

This study was performed in accordance with the ethical principles consistent with the latest version of the Declaration of Helsinki (version Fortaleza 2013), Good Clinical Practice (GCP) and according to the Australian National Health and Medical Research Council Guidelines for Research published in the National Statement on the Ethical Conduct in Human Research (2007; updated 2018). 146 healthy adult participants were recruited in Adelaide, South Australia between April 8^th^ and November 1^st^, 2021 under protocols approved by the CALHN Human Research Ethics Committee, Adelaide, Australia (Approval No. 14778) or by the Royal Children’s Hospital, Melbourne, Australia, Human Research Ethics Committee (Approval No. 62586). Strict international and interstate border control measures to prevent community transmission of SARS-CoV2 in South Australia were in place until late November 2021. Participant inclusion criteria were being healthy, aged 18+ years, voluntary informed consent, and the ability to attend study follow-up visits. Exclusion criteria included any previous positive SARS-CoV-2 test prior to vaccination, a COVID-19 vaccine administered prior to baseline blood sample collection, or any other vaccines received 14 days prior to COVID-19 vaccine.

Participants received two doses of either the BNT162b2 (n=86) or ChAdOx1-S (n=16) vaccines for their 1^st^/2^nd^ vaccinations. Participants were asked to provide a blood sample before their 1^st^COVID-19 vaccine and then ∼6 days after the 1^st^ dose (V1), and ∼1-2 days (V2A) and ∼28 days after the 2^nd^ dose (V2B). In a subset of participants, blood samples were collected ∼1-2 days (V3A) and ∼28 days (V3B) after a 3^rd^ dose of BNT162b2 (n=38) or mRNA-1273 (Moderna, n=14) vaccine. Venous blood (∼29 ml/individual) was collected in multiple blood tubes including serum separator (CAT serum separator clot activator) tubes processed for serum; sodium citrate tubes processed for plasma; sodium heparin tubes for peripheral blood mononuclear cell (PBMC) isolation; and lithium heparin tubes for whole blood preservation in RNAlater solution (Thermo Fisher Scientific, Waltham, USA) or FACSlyse buffer (BD Biosciences, Franklin Lakes, USA) used for RNA extraction and flow cytometry analysis, respectively. Blood tubes were processed within 3 hours of collection. A survey recording any symptoms of vaccine reactogenicity was sent to participants for completion one week after each vaccination (**Supplementary File 1**). The 3^rd^ survey also captured any known COVID-19 positive events among participants when community transmission began to occur in South Australia in December 2021, following the lifting of border restrictions.

### SARS-CoV-2 protein purification and ELISA

Recombinant SARS-CoV-2 spike protein and receptor-binding domain (RBD) were produced for ELISA. Briefly, Prefusion SARS-CoV-2 ectodomain (isolate WHU1, residues1-1208) with HexaPro mutations (Hsieh et al., 2020) (kindly provided by Adam Wheatley) and SARS-Cov-2 receptor-binding domain (RBD) with C-terminal His-tag (Amanat et al., 2020) (residues 319-541; kindly provided by Florian Krammer) were overexpressed in Expi293 cells and purified by Ni-NTA affinity and size-exclusion chromatography. Recombinant proteins were analyzed via a standard SDS-PAGE gel to check protein integrity. Gels were stained with Comassie Blue (Invitrogen) for 2 h and de-stained in distilled water overnight. For ELISA, MaxiSorp 96-well plates were coated overnight at 4°C with 5 µg/ml of recombinant RBD or Spike proteins. After blocking with 5% w/v skim milk in 0.05% Tween-20/Phosphate-Buffered Saline (PBST) at room temperature (RT), serially diluted (heat inactivated) sera were added and incubated for 2 h at RT. Plates were washed 4 times with 0.05% PBST and secondary antibodies added. Secondary antibodies were diluted in 5% skim milk in PBST as follows: Goat anti-Human IgG (H+L) Secondary Antibody HRP (1:30,000; Invitrogen) and incubated for 1 h at RT. Plates were developed with 1-Step™ Ultra TMB Substrate (Thermo Fisher) and stopped with 2M sulphuric acid. OD readings were read at 450 nm on a Synergy HTX Multi-Mode Microplate Reader. AUC calculation was performed using Prism (GraphPad, CA, USA), where the X-axis is half log_10_ of sera dilution against OD_450_ on the Y-axis.

### SARS-CoV-2 pseudovirus neutralization assay

SARS-CoV-2 neutralization assays were performed as previously described (Abayasingam et al., 2021). In brief, SARS-CoV-2 pseudovirus were generated by co-transfecting expression plasmids containing SARS-CoV-2 Spike (kindly provided by Dr Markus Hoffmann (Hoffmann et al., 2020)), and the murine leukemia virus (MLV) gag/pol and luciferase vectors (kindly provided by Prof. Francois-Loic Cosset (Keck et al., 2009, Bartosch et al., 2003)), in CD81KO 293T cells (kindly provided by Dr Joe Grove (Kalemera et al., 2020)), using the mammalian Calphos transfection kit (Takara Bio, Shiga, Japan). Pseudovirus culture supernatants were harvested 48 h post transfection and concentrated 10-fold using 100,000 MWCO Vivaspin centrifugal concentrators (Sartorius, Göttingen, Germany) by centrifugation at 2000 x *g* and stored at –80°C. For neutralization assays, pseudovirus was diluted in media to be 1000 – 5000-fold more infectious than negative background (based on pseudovirus lacking SARS-CoV-2 Spike). Diluted pseudovirus were incubated for 1 h with heat inactivated (56°C for 30 min) participant serum, followed by the addition of polybrene at a final concentration of 4 µg/ml (Merck, St. Louis, USA), prior to addition to 293T-ACE2 over-expressed cells (kindly provided by A/Prof Jesse Bloom (Crawford et al., 2020)). 293T-ACE2 cells were seeded 24 h earlier at 1.5 × 10^4^ cells per well in 96-well, white flat-bottom plates (Merck). Cells were spinoculated at 800 x *g* for 2 h and incubated for 2 h at 37°C, prior to media change. After 72 h, the cells were lysed with a lysis buffer (Promega, Madison, USA) and Bright Glo reagent (Promega) was added at a 1:1 ratio. Luminescence (RLU) was measured using CLARIOstar microplate reader (BMG Labtech, Ortenberg, Germany). Neutralization assays were performed in triplicates. Percentage neutralization of pseudovirus was calculated as (1 – RLU_treatment_/RLU_no treatment_) × 100. The 50% inhibitory concentration (ID_50_) titer was calculated using a non-linear regression model in Prism (GraphPad, CA, USA).

### PBMC isolation

Sodium-heparin anticoagulated whole blood (8-12 ml) was diluted in 12 ml of endotoxin-free sterile PBS, layered onto 12 ml Lymphoprep (Stem Cell Technologies, Vancouver, Canada) and spun at 800 x *g* for 30 min, RT, with no brake. PBMC were isolated from the resultant interphase layer via pipetting, washed twice in PBS, pelleted at 600 x *g* for 5 min at RT, and resuspended in 1.5 ml freezing medium (10% DMSO, 90% FCS). PBMC (500 µl) were aliquoted into cryovials, placed in a Mr. Frosty™ Freezing Container (Nalgene, Rochester, USA) and stored at -80°C overnight. Frozen PBMC cryovials were transferred to liquid nitrogen for long-term storage within 1-2 days.

### Flow cytometry analysis of immune cell populations in whole blood

Sodium-heparin anticoagulated whole blood (200 μl) was incubated with 1700 µl FACS™ Lysing Solution (BD Biosciences) for 10 min at RT, mixing 3 times. Samples were centrifuged at 600 x *g* for 5 min, the supernatant aspirated and the cells resuspended in 400 μl FACSLyse. 200 μl aliquots were snap frozen at -80°C and transferred to liquid nitrogen within 1 week for long term storage. For analysis, samples were rapidly thawed at 37°C in batches, plated into a 96 well U-bottom plate on ice and spun at 600 x *g* for 5 min. Samples were then washed twice in 240 μl FACS buffer (PBS, 0.1% BSA, 2mM EDTA), and split into two different panels. Cells were incubated with 3 μl TruStain FcX (Biolegend, San Diego, USA) in 15 μl Brilliant Violet Stain buffer (BD Biosciences) on ice for 15 min and then stained with of one of two master-mixes of antibodies (a 13-color pan-leukocyte panel or a 17-color lymphocyte panel) in 30 μl final volume for 30 min on ice. The stained cells were washed with 200 µl FACS buffer, centrifuged 600 x *g* for 5 min and the pan-leukocyte panel was resuspended in FACS buffer for analysis on a flow cytometer, while the lymphocyte panel was resuspended in 100 μl 1x Permeabilization Buffer (Foxp3/Transcription Factor Staining Buffer Set, eBioscience, San Diego, USA) and incubated on ice for 10 min. The lymphocyte panel was spun at 600 x *g* for 5 min and stained with anti-Granzyme B (BD Biosciences) in 30 μl 1 x Perm buffer (BD Biosciences) on ice for 30 min. Samples were then washed once with 200 µl Perm buffer and once with 200 µl FACS buffer, centrifuging for 600 x *g* for 5 min between washes. Cells were then resuspended in FACS buffer and run immediately on a BD FACS Symphony flow cytometer (BD Biosciences). Liquid counting beads (BD Biosciences) were added to each sample to enumerate cells.

### Flow cytometry data acquisition & analysis

To control for batch effects, the BD FACS symphony lasers were calibrated with dye conjugated standards (Cytometer Set &Track beads) run every day. All voltages of photomultiplier tubes (PMT)s were adjusted to negative unstained control baseline typically log scale 10^2^. Antibodies were titrated for optimal signal over background prior to use. Compensation was set with beads matched to each panel antibody combination using spectral compensation using FlowJo Software v18 (FlowJo, Ashland, USA). Additionally, all samples across timepoints from each individual participant were analyzed in a single batch. A SSC-A/FSC-A plot was used to determine sample size and complexity. Lymphocytes, monocyte, and granulocyte gates were based on physical parameters. Events were gated for FCS-A as well as SSC-A linearity, and restricted FSC-H values for doublet discrimination. Populations of cells were expressed as a proportion of parent gated cells, proportion of the highest order lineage gate namely: Lymphocytes, B cells, T cells or CD45^+^ cells, or as absolute cells per ml of blood. Gating strategy for both panels are shown in **(Supplementary File 2)**. Flow cytometry data were imported into *R* v4.2 for further statistical analyses. Changes in cell counts or activation was assessed using a mixed effect linear model implemented in *R* v4.2 (glm function) controlling for individual and processing batch. The Benjamini and Hochberg method was employed to correct for multiple comparisons. Statistical significance was determined as FDR < 0.05. Data visualizations were generated using the *Spectre* v1.0.0 (Ashhurst et al., 2022) and *ggplot2* v3.3.6 (Wickham, 2016) packages.

### Activation Induced Marker (AIM) assay

Cryopreserved PBMCs were thawed and 10^6^ live cells were plated out in 96 well U-bottom plates. Spike protein peptide pool (GenScript, New Jersey, USA) was added to each well at 1 µg/ml and cells incubated for 20-24 h at 37°C. For surface staining of AIM markers, cells were incubated in 1 µg/ml human Fc block (BD Biosciences) and fixable viability solution 780 (BD Biosciences) in PBS for 15 min and washed in PBS. An antibody cocktail containing surface antibodies against CD3 (clone UCHT1, BD Biosciences), CD4 (clone M-T477, BD Biosciences), CD8 (G42-8, BD Biosciences), CD69 (clone FN50, BD Biosciences), CD137 (clone 4B4-1, Biolegend), OX40 (clone Ber-ACT35, Biolegend), CCR7 (clone 2-L1-A, BD Biosciences), and CD45RA (clone HI100, BD Biosciences) was added directly to cells and incubated for a further 30 min at 4°C. Following surface staining, cells were washed twice in PBS. All samples were acquired on a BD FACS Symphony and analyzed using FlowJO software v18 (FlowJo, Ashland, USA). Data were imported into *R* v4.2 and visualized with the *ggplot2* v3.3.6 package.

### ELISpot assay

ELISpot plates (MERCK-Millipore, Burlington, Vermont, USA) were coated with human IFNγ capture antibody (1-D1K, Mabtech, Stockholm, Sweden; 5 μg/ml) overnight at 4°C.

Plates were blocked with 1x ELISA/ELISpot blocking buffer (Thermo Fisher Scientific) for at least 1 h at RT then 2x10^5^ thawed PBMCs were seeded per well and stimulated for 20-24h with pools of SARS-CoV-2 peptides (GenScript, 1 μg/ml). After stimulation, plates were washed 3x in PBS-T and incubated for 2 h with human biotinylated IFNγ detection antibody (7-B6-1, Mabtech; 1:500). Plates were then washed 3x with PBS-T followed by a 1 h incubation with streptavidin-HRP (BD biosciences, 1:1000) and developed with 3-Amino-9-ethylcarbazole (AEC) substrate (BD Biosciences). Spot forming units (SFU) were quantified with ImmunoSpot software (Cellular Technology Limited, Ohio, USA). Results were expressed as IFNγ-SFU/10^6^ PBMCs. Data were imported into *R* v4.2 and visualized with *ggplot2* v3.3.6.

### Human anti-PF4 ELISA

For anti-PF4 ELISAs, a commercial kit (Abbexa, Cambridge, UK) was used as per manufacturer’s instructions. Human serum samples were diluted 1:50 in PBS. Plates were read on a Synergy HTX Multi-mode plate reader (Biotek) at 450nm. Data were imported into *R* v4.2 and visualized with *ggplot2* v3.3.6.

### RNA extraction and library preparation

RNA extraction and genomic DNA elimination was carried out using the RiboPure™ RNA Purification kit for blood (Invitrogen) as per the manufacturer’s instructions. Final elution into 50 μl RNase-free water. A further RNA precipitation reaction was carried out. Briefly, RNA was resuspended 2.5x in 100% ethanol and 10% sodium acetate (Merck) and spun at 12,000 x *g* for 15 min at 4°C. Samples were washed in 75% ethanol. Pellets were air dried and resuspended in 29 μl RNase free water and total RNA yield was determined by analysis of samples using a TapeStation (Agilent, Santa Clara, USA) and Qubit (ThermoFisher Scientific, Waltham, USA). Total RNA was converted to strand-specific Illumina compatible sequencing libraries using the Nugen Universal Plus Total RNASeq library kit from Tecan (Mannedorf, Switzerland) as per the manufacturer’s instructions (MO1523 v2) using 12 cycles of PCR amplification for the final libraries. An Anydeplete probe mix, targeting both human ribosomal and adult globin transcripts (HBA1, HBA2, HBB, HBD), was used to deplete these transcripts. The final library pool was converted to an MGI compatible circularized template using the MGIEasy Universal Library Conversion Kit (MGI, Shenzhen, China) before sequencing of the library pool (2x150 bp paired-end reads) was performed on an MGI DNBSEQ-G400.

### RNASeq analysis

Sequence read quality was assessed using *FastQC* version 0.11.9 (Andrews, 2010) and summarized with *MultiQC* version 1.10.1 (Ewels et al., 2016) prior to quality control with *Trimmomatic* version 0.38 (Bolger et al., 2014) with a window size of 4 nucleotides and an average quality score of 30. Following this, reads which were <50 nucleotides after trimming were discarded. Reads that passed all quality control steps were then aligned to the human genome (GRCh38 assembly) using *HISAT2* version 2.1.0 (Kim et al., 2015). The gene count matrix was generated with *FeatureCounts* version 1.5.0-p2 (Liao et al., 2014) with Ensembl version 101 annotation (Howe et al., 2021). The count matrix was then imported into *R* 4.2 for further analysis and visualization in *ggplot2* v3.3.6. Counts were normalized using the trimmed mean of M values (TMM) method in *EdgeR* version 3.38 (Robinson et al., 2010) and represented as counts per million (cpm). Prior to multidimensional scaling analysis *svaseq* v3.44 was applied to remove batch effects and other unwanted sources of variation in the data (Leek, 2014). Differential gene expression analysis was performed using the glmLRT function in *EdgeR* adjusting for sex and batch (run) in the model. Genes with <3 cpm in at least 15 samples were excluded from the differential expression analysis. The Benjamini and Hochberg method was employed to correct for multiple comparisons.

Statistical significance was determined as FDR < 0.05. Pathway and Gene Ontology (GO) overrepresentation analysis was carried out in *R* using a hypergeometric test. Cell type enrichment analysis was carried out in *R* using the using the camera function in the *EdgeR* library with cell type gene sets from the human cell atlas bone marrow dataset in the Molecular Signatures database collection C8 (*R* package *msigdbr* v7.5.1) (Dolgalev, 2020). Blood Transcriptional Module (BTM) analysis was carried out using a pre-defined set of modules defined by Li *et al*. as an alternative to pathway-based analyses (Li et al., 2014). Gene Set Variation Analysis (GSVA) (Hänzelmann et al., 2013) was used to calculate a per sample activity score for each of the modules (excluding unannotated modules labelled as ‘TBA’). *limma* v3.51.0 (Ritchie et al., 2015) was used to identify modules that were differentially active at at least one timepoint.

### Proteomics analysis

Plasma samples were processed using the USP^3^ protocol as outlined previously (Dagley et al., 2019). Briefly, 2 μl plasma samples were reduced/alkylated in 1% SDS lysis buffer using 10 mM Tris (2-Carboxyethyl) phosphine (TCEP, Merck) and 40 mM 2-chloracetamide (2-CAA, Merck) for 10 min at 95°C. Sample volumes containing 25 μg protein were transferred to a 0.5 ml low-bind deep-well plate (Eppendorf, Hamburg, Germany) and were incubated with 20 μl of magnetic PureCube Carboxy agarose beads (Cube Biotech, Monheim, Germany) (pre-washed three times in MilliQ water) and acetonitrile (Merck, 70% v/v) for 20 min at RT using the ThermoMixer C (Eppendorf) shaking at 400 rpm. Samples were placed on a magnetic rack, supernatant removed, and three 200 μl washes performed: twice with 70% ethanol, and one with neat acetonitrile (ACN). After washing, all traces of solvent were evaporated using a CentriVap (Labconco, Kansas City, USA), and digestion buffer (100 mM Tris-HCl, pH 8) was added to the beads containing Lys-C (Wako, Chuo-Ku, Japan) and SOLu-Trypsin (Merck) each at a 1:50 enzyme to protein ratio, and digestion performed for 1 h at 37°C with shaking at 400 rpm. Digests were desalted using in-house C18 stage tips containing 2 plugs as previously described (Rappsilber et al., 2007), vacuum centrifuged to dryness, and peptides reconstituted in 75 μl 0.1% formic acid (Merck) and 2% ACN (Merck) in preparation for LC-MS/MS. Mass spectrometry analyses were performed on a M-class (Waters, Milford, USA) coupled to a timsTOF Pro (Bruker, Billerica, USA) equipped with a CaptiveSpray source. Peptides (1 µl) were separated by reverse-phase chromatography on a C_18_ fused silica column (inner diameter 75 µm, OD 360 µm × 15 cm length, 1.6 µm C18 beads) packed into an emitter tip (IonOpticks, Middle Camberwell, Australia), using a nano-flow HPLC (M-class, Waters). The HPLC was coupled to a timsTOF Pro (Bruker) equipped with a CaptiveSpray source. Peptides were loaded directly onto the column at a constant flow rate of 400 nl/min with buffer A (99.9% Milli-Q water, 0.1% FA) and eluted with a 90-min linear gradient from 2 to 34% buffer B (99.9% ACN, 0.1% FA). The timsTOF Pro (Bruker) was operated in diaPASEF mode using Compass Hystar 5.1. The settings on the TIMS analyzer were as follows: Lock Duty Cycle to 100% with equal accumulation and ramp times of 100 ms, and 1/K0 Start 0.6 V·s/cm^2^ End 1.6 V·s/cm^2^, Capillary Voltage 1400V, Dry Gas 3 l/min, Dry Temp 180°C. Methods were set up using the instrument firmware (timsTOF control 2.0.18.0) for data-independent isolation of multiple precursor windows within a single TIMS scan. The method included two windows in each diaPASEF scan, with window placement overlapping the diagonal scan line for doubly and triply charged peptides in the m/z – ion mobility plane across 16 × 25 m/z precursor isolation windows (resulting in 32 windows) defined from *m/z* 400 to 1,200, with 1 Da overlap, with CID collision energy ramped stepwise from 20 eV at 0.8 V·s/cm^2^ to 59eV at 1.3 V·s/cm^2^. DIA-NN 1.8 (Demichev et al., 2021) was used for searching of timsTOF diaPASEF .d files. The option for library-free search was enabled, and data were searched against reviewed sequences from the human

Uniprot Reference Proteome (downloaded May 2021). The search was set to trypsin specificity, peptide length of 7-30 residues, cysteine carbidomethylation as a fixed modification, and the maximum number of missed cleavages at 2. Additionally, n-terminal acetylation and oxidation of methionine were set as variable modifications with the maximum number of variable modifications set to 1. In addition, the maximum mass accuracy was set to 10 ppm for both MS1 and MS2 spectra and the quantification strategy to robust LC (high precision). Match between runs (MBR) was enabled, and all other settings left as default. For reporting precursor and protein numbers, outputs were filtered at precursor q-value < 1% and PG protein q-value < 1%, and the PG.MaxLFQ was used to obtain the normalized quantity for protein groups based on proteotypic peptides (i.e. unique proteins). Data processing and analysis were performed using *R* v4.2. Only proteins that were quantified in at least 50% of replicates in at least one condition were kept. The data were normalized using variance stabilizing method implemented in *limma* v3.51.0. Differential analysis was performed using *limma* v3.51.0. The Benjamini and Hochberg method was employed to correct for multiple comparisons. Statistical significance was determined as FDR < 0.1.

### Lipidomics

Lipid extraction was performed as previously described (Huynh et al., 2019). In brief, 10 µl of plasma was mixed with 100 µl of butanol:methanol (1:1) with 10 mM ammonium formate which contained a mixture of internal standards. Samples were then vortexed and set in a sonicator bath for 1 h at RT. Samples were then centrifuged (14,000 x *g*, 10 min, 20°C) before transferring into the sample vials for analysis. Analysis of plasma extracts was performed on an Agilent 6495C QQQ mass spectrometer with an Agilent 1290 series HPLC system and a ZORBAX eclipse plus C18 column (2.1x100 mm 1.8 mm, Agilent) with the thermostat set at 45°C. Mass spectrometry analysis was performed with dynamic scheduled multiple reaction monitoring (dMRM). Chromatogram integration was performed using Agilent MassHunter v10.0 and quantification of lipid species was determined by comparison to the relevant internal standard. Differential analysis was performed using a mixed effect linear model implemented in R (glm function). The Benjamini and Hochberg method was employed to correct for multiple comparisons. Statistical significance was determined as FDR < 0.1. Data were visualized with *ggplot2* v3.3.6.

### Multiplex cytokine analysis

A flow cytometric bead multiplex assay (LEGENDplex™ Human Anti-Virus Response 13-plex Panel, Biolegend) was used to quantify 13 cytokines (IL-1β, IL-6, IL-8, IL-10, IL-12p70, IFN-α, IFN-β, IFN-λ1, IFN-λ2/3, IFN-γ, TNF-α, IP-10/CXCL10 & GM-CSF) in sodium citrate plasma samples collected at V0, V1, V2A and V3A, according to the manufacturer’s instructions. Briefly, samples were thawed at RT and centrifuged (2000 x *g*, 5 min, 4°C) prior to 1:1 dilution with Assay Buffer (Biolegend). 9.5 μl of diluted sample was mixed with 9.5 μl Assay Buffer and 9.5 μl premixed capture beads in a V-bottom plate and incubated protected from light (200 x *g*, 2 h, RT). Samples were washed (200 μl LEGENDplex™ wash buffer) and incubated for a further 1 h (200 x *g* RT) with 9.5 μl premixed detection antibody before the addition of 9.5 μl Streptavidin-PE and another 30 min incubation. Samples were washed (150 μl wash buffer) before resuspension in 150 μl washing buffer for acquisition. Samples were immediately acquired on a BD LSR Fortessa X-20 (BD Biosciences) with High Throughput Sampler. Bead populations were identified based on forward/side scatter and the degree of APC-H7 fluorescence. PE fluorescence (geometric mean fluorescence intensity) was determined for each sample and cytokine concentrations were interpolated from a standard curve (sigmoidal 4-parameter logistic curve) using GraphPad Prism 9 software. Differential analysis was performed using a mixed effect linear model implemented in *R* (glm function). Data were visualized with the *ggplot2* v3.3.6 package.

### Multi-omics correlation analysis

Spearman correlation analysis was performed using the *Hmisc v4.7-0* package in *R* v4.2 (Harrell Jr and Harrell Jr, 2019). Statistical significance was defined as p < 0.05. In order to avoid spurious correlations driven by differences in antibody titers or T-cell responses induced following the ChAdOx1-S and BNT162b2 vaccines, the correlation analyses were performed for each vaccine group separately for antibody tires at V2B. At V3B we did not detect significant differences in antibody titers between the different vaccine groups, so all samples were used for the correlation analysis. A subset of significant Spearman correlations were also evaluated with a generalized linear model adjusting for participant meta data to ensure the relationships were robust to sex and age.

## References

Abayasingam, A., Balachandran, H., Agapiou, D., Hammoud, M., Rodrigo, C., Keoshkerian, E., Li, H., Brasher, N. A., Christ, D., Rouet, R., Burnet, D., Grubor-Bauk, B., Rawlinson, W., Turville, S., Aggarwal, A., Stella, A. O., Fichter, C., Brilot, F., Mina, M., Post, J. J., Hudson, B., Gilroy, N., Dwyer, D., Sasson, S. C., Tea, F., Pilli, D., Kelleher, A., Tedla, N., Lloyd, A. R., Martinello, M., Bull, R. A. & Group, C. S. 2021. Long-term persistence of RBD(+) memory B cells encoding neutralizing antibodies in SARS-CoV-2 infection. Cell Rep Med, 2, 100228.

Amanat, F., Stadlbauer, D., Strohmeier, S., Nguyen, T. H., Chromikova, V., Mcmahon, M., Jiang, K., Arunkumar, G. A., Jurczyszak, D. & Polanco, J. 2020. A serological assay to detect SARS-CoV-2 seroconversion in humans. Nature medicine, 26, 1033–1036.

Andrews, N., Stowe, J., Kirsebom, F., Toffa, S., Rickeard, T., Gallagher, E., Gower, C., Kall, M., Groves, N., O’connell, A. M., Simons, D., Blomquist, P. B., Zaidi, A., Nash, S., Iwani Binti Abdul Aziz, N., Thelwall, S., Dabrera, G., Myers, R., Amirthalingam, G., Gharbia, S., Barrett, J. C., Elson, R., Ladhani, S. N., Ferguson, N., Zambon, M., Campbell, C. N. J., Brown, K., Hopkins, S., Chand, M., Ramsay, M. & Lopez Bernal, J. 2022. Covid-19 Vaccine Effectiveness against the Omicron (B.1.1.529) Variant. N Engl J Med, 386, 1532–1546.

Andrews, S. 2010. FastQC: A Quality Control Tool for High Throughput Sequence Data.

Arunachalam, P. S., Scott, M. K. D., Hagan, T., Li, C., Feng, Y., Wimmers, F., Grigoryan, L., Trisal, M., Edara, V. V., Lai, L., Chang, S. E., Feng, A., Dhingra, S., Shah, M., Lee, A. S., Chinthrajah, S., Sindher, S. B., Mallajosyula, V., Gao, F., Sigal, N., Kowli, S., Gupta, S., Pellegrini, K., Tharp, G., Maysel-Auslender, S., Hamilton, S., Aoued, H., Hrusovsky, K., Roskey, M., Bosinger, S. E., Maecker, H. T., Boyd, S. D., Davis, M. M., Utz, P. J., Suthar, M. S., Khatri, P., Nadeau, K. C. & Pulendran, B. 2021. Systems vaccinology of the BNT162b2 mRNA vaccine in humans. Nature, 596, 410–416.

Ashhurst, T. M., Marsh-Wakefield, F., Putri, G. H., Spiteri, A. G., Shinko, D., Read, M. N., Smith, A. L. & King, N. J. C. 2022. Integration, exploration, and analysis of high-dimensional single-cell cytometry data using Spectre. Cytometry A, 101, 237–253.

Baden, L. R., El Sahly, H. M., Essink, B., Kotloff, K., Frey, S., Novak, R., Diemert, D., Spector, S. A., Rouphael, N., Creech, C. B., Mcgettigan, J., Khetan, S., Segall, N., Solis, J., Brosz, A., Fierro, C., Schwartz, H., Neuzil, K., Corey, L., Gilbert, P., Janes, H., Follmann, D., Marovich, M., Mascola, J., Polakowski, L., Ledgerwood, J., Graham, B. S., Bennett, H., Pajon, R., Knightly, C., Leav, B., Deng, W., Zhou, H., Han, S., Ivarsson, M., Miller, J., Zaks, T. & Group, C. S. 2021. Efficacy and Safety of the mRNA-1273 SARS-CoV-2 Vaccine. N Engl J Med, 384, 403–416.

Baker, A. T., Boyd, R. J., Sarkar, D., Teijeira-Crespo, A., Chan, C. K., Bates, E., Waraich, K., Vant, J., Wilson, E., Truong, C. D., Lipka-Lloyd, M., Fromme, P., Vermaas, J., Williams, D., Machiesky, L., Heurich, M., Nagalo, B. M., Coughlan, L., Umlauf, S., Chiu, P. L., Rizkallah, P. J., Cohen, T. S., Parker, A. L., Singharoy, A. & Borad, M. J. 2021. ChAdOx1 interacts with CAR and PF4 with implications for thrombosis with thrombocytopenia syndrome. Sci Adv, 7, eabl8213.

Bánki, Z., Mateus, J., Rössler, A., Schäfer, H., Bante, D., Riepler, L., Grifoni, A., Sette, A., Simon, V. & Falkensammer, B. 2022. Heterologous ChAdOx1/BNT162b2 vaccination induces stronger immune response than homologous ChAdOx1 vaccination: The pragmatic, multi-center, three-arm, partially randomized HEVACC trial. EBioMedicine, 80, 104073.

Barda, N., Dagan, N., Ben-Shlomo, Y., Kepten, E., Waxman, J., Ohana, R., Hernan, M. A., Lipsitch, M., Kohane, I., Netzer, D., Reis, B. Y. & Balicer, R. D. 2021. Safety of the BNT162b2 mRNA Covid-19 Vaccine in a Nationwide Setting. N Engl J Med, 385, 1078–1090.

Bartolo, L., Afroz, S., Pan, Y. G., Xu, R., Williams, L., Lin, C. F., Tanes, C., Bittinger, K., Friedman, E. S., Gimotty, P. A., Wu, G. D. & Su, L. F. 2022. SARS-CoV-2-specific T cells in unexposed adults display broad trafficking potential and cross-react with commensal antigens. *Sci Immunol*, eabn3127.

Bartosch, B., Bukh, J., Meunier, J. C., Granier, C., Engle, R. E., Blackwelder, W. C., Emerson, S. U., Cosset, F. L. & Purcell, R. H. 2003. In vitro assay for neutralizing antibody to hepatitis C virus: evidence for broadly conserved neutralization epitopes. Proc Natl Acad Sci U S A, 100, 14199–204.

Bentebibel, S. E., Lopez, S., Obermoser, G., Schmitt, N., Mueller, C., Harrod, C., Flano, E., Mejias, A., Albrecht, R. A., Blankenship, D., Xu, H., Pascual, V., Banchereau, J., Garcia-Sastre, A., Palucka, A. K., Ramilo, O. & Ueno, H. 2013. Induction of ICOS+CXCR3+CXCR5+ TH cells correlates with antibody responses to influenza vaccination. Sci Transl Med, 5, 176ra32.

Bergamaschi, C., Terpos, E., Rosati, M., Angel, M., Bear, J., Stellas, D., Karaliota, S., Apostolakou, F., Bagratuni, T., Patseas, D., Gumeni, S., Trougakos, I. P., Dimopoulos, M. A., Felber, B. K. & Pavlakis, G. N. 2021. Systemic IL-15, IFN-gamma, and IP-10/CXCL10 signature associated with effective immune response to SARS-CoV-2 in BNT162b2 mRNA vaccine recipients. Cell Rep, 36, 109504.

Biram, A., Liu, J., Hezroni, H., Davidzohn, N., Schmiedel, D., Khatib-Massalha, E., Haddad, M., Grenov, A., Lebon, S., Salame, T. M., Dezorella, N., Hoffman, D., Abou Karam, P., Biton, M., Lapidot, T., Bemark, M., Avraham, R., Jung, S. & Shulman, Z. 2022. Bacterial infection disrupts established germinal center reactions through monocyte recruitment and impaired metabolic adaptation. Immunity, 55, 442–458.e8.

Bolger, A. M., Lohse, M. & Usadel, B. 2014. Trimmomatic: a flexible trimmer for Illumina sequence data. Bioinformatics, 30, 2114–20.

Brewer, J. W., Solodushko, V., Aragon, I. & Barrington, R. A. 2016. Phosphatidylcholine as a metabolic cue for determining B cell fate and function. Cell Immunol, 310, 78–88.

Collier, D. A., Ferreira, I., Kotagiri, P., Datir, R. P., Lim, E. Y., Touizer, E., Meng, B., Abdullahi, A., Collaboration, C.-N. B. C.-., Elmer, A., Kingston, N., Graves, B., Le Gresley, E., Caputo, D., Bergamaschi, L., Smith, K. G. C., Bradley, J. R., Ceron-Gutierrez, L., Cortes-Acevedo, P., Barcenas-Morales, G., Linterman, M. A., Mccoy, L. E., Davis, C., Thomson, E., Lyons, P. A., Mckinney, E., Doffinger, R., Wills, M. & Gupta, R. K. 2021. Age-related immune response heterogeneity to SARS-CoV-2 vaccine BNT162b2. Nature, 596, 417–422.

Cornaby, C., Gibbons, L., Mayhew, V., Sloan, C. S., Welling, A. & Poole, B. D. 2015. B cell epitope spreading: mechanisms and contribution to autoimmune diseases. Immunol Lett, 163, 56–68.

Crawford, K. H. D., Eguia, R., Dingens, A. S., Loes, A. N., Malone, K. D., Wolf, C. R., Chu, H. Y., Tortorici, M. A., Veesler, D., Murphy, M., Pettie, D., King, N. P., Balazs, A. B. & Bloom, J. D. 2020. Protocol and Reagents for Pseudotyping Lentiviral Particles with SARS-CoV-2 Spike Protein for Neutralization Assays. Viruses, 12, 513.

Dagley, L. F., Infusini, G., Larsen, R. H., Sandow, J. J. & Webb, A. I. 2019. Universal Solid-Phase Protein Preparation (USP3) for Bottom-up and Top-down Proteomics. Journal of Proteome Research, 18, 2915–2924.

Demichev, V., Yu, F., Teo, G. C., Szyrwiel, L., Rosenberger, G. A., Decker, J., Kaspar-Schoenefeld, S., Lilley, K. S., Mülleder, M., Nesvizhskii, A. I. & Ralser, M. 2021. High sensitivity dia-PASEF proteomics with DIA-NN and FragPipe. bioRxiv, 2021.03.08.434385.

Dicks, M. D., Spencer, A. J., Edwards, N. J., Wadell, G., Bojang, K., Gilbert, S. C., Hill, A. V. & Cottingham, M. G. 2012. A novel chimpanzee adenovirus vector with low human seroprevalence: improved systems for vector derivation and comparative immunogenicity. PLoS One, 7, e40385.

Dolgalev, I. 2020. msigdbr: MSigDB gene sets for multiple organisms in a tidy data format. R package version, 7.

Ewels, P., magnusson, M., Lundin, S. & Kaller, M. 2016. MultiQC: summarize analysis results for multiple tools and samples in a single report. Bioinformatics, 32, 3047–8.

Falsey, A. R., Sobieszczyk, M. E., Hirsch, I., Sproule, S., Robb, M. L., Corey, L., Neuzil, K. M., Hahn, W., Hunt, J., Mulligan, M. J., Mcevoy, C., Dejesus, E., Hassman, M., Little, S. J., Pahud, B. A., Durbin, A., Pickrell, P., Daar, E. S., Bush, L., Solis, J., Carr, Q. O., Oyedele, T., Buchbinder, S., Cowden, J., Vargas, S. L., Guerreros Benavides, A., Call, R., Keefer, M. C., Kirkpatrick, B. D., Pullman, J., Tong, T., Brewinski Isaacs, M., Benkeser, D., Janes, H. E., Nason, M. C., Green, J. A., Kelly, E. J., Maaske, J., Mueller, N., Shoemaker, K., Takas, T., Marshall, R. P., Pangalos, M. N., Villafana, T., Gonzalez-Lopez, A. & Astrazeneca, A. Z. D. C. S. G. 2021. Phase 3 Safety and Efficacy of AZD1222 (ChAdOx1 nCoV-19) Covid-19 Vaccine. N Engl J Med, 385, 2348–2360.

Feng, S., Phillips, D. J., White, T., Sayal, H., Aley, P. K., Bibi, S., Dold, C., Fuskova, M., Gilbert, S. C., Hirsch, I., Humphries, H. E., Jepson, B., Kelly, E. J., Plested, E., Shoemaker, K., Thomas, K. M., Vekemans, J., Villafana, T. L., Lambe, T., Pollard, A. J., Voysey, M. & Oxford, C. V. T. G. 2021. Correlates of protection against symptomatic and asymptomatic SARS-CoV-2 infection. Nat Med, 27, 2032–2040.

Folegatti, P. M., Ewer, K. J., Aley, P. K., Angus, B., Becker, S., Belij-Rammerstorfer, S., Bellamy, D., Bibi, S., Bittaye, M., Clutterbuck, E. A., Dold, C., Faust, S. N., Finn, A., Flaxman, A. L., Hallis, B., Heath, P., Jenkin, D., Lazarus, R., Makinson, R., Minassian, A. M., Pollock, K. M., Ramasamy, M., Robinson, H., Snape, M., Tarrant, R., Voysey, M., Green, C., Douglas, A. D., Hill, A. V. S., Lambe, T., Gilbert, S. C., Pollard, A. J. & Oxford, C. V. T. G. 2020. Safety and immunogenicity of the ChAdOx1 nCoV-19 vaccine against SARS-CoV-2: a preliminary report of a phase 1/2, single-blind, randomised controlled trial. Lancet, 396, 467–478.

Geurtsvankessel, C. H., Geers, D., Schmitz, K. S., Mykytyn, A. Z., Lamers, M. M., Bogers, S., Scherbeijn, S., Gommers, L., Sablerolles, R. S. G., Nieuwkoop, N. N., Rijsbergen, L. C., Van Dijk, L. L. A., De Wilde, J., Alblas, K., Breugem, T. I., Rijnders, B. J. A., De Jager, H., Weiskopf, D., Van Der Kuy, P. H. M., Sette, A., Koopmans, M. P. G., Grifoni, A., Haagmans, B. L. & De Vries, R. D. 2022. Divergent SARS-CoV-2 Omicron-reactive T and B cell responses in COVID-19 vaccine recipients. Sci Immunol, 7, eabo2202.

Greinacher, A., Thiele, T., Warkentin, T. E., Weisser, K., Kyrle, P. A. & Eichinger, S. 2021. Thrombotic Thrombocytopenia after ChAdOx1 nCov-19 Vaccination. N Engl J Med, 384, 2092–2101.

Hänzelmann, S., Castelo, R. & Guinney, J. 2013. GSVA: gene set variation analysis for microarray and RNA-Seq data. BMC Bioinformatics, 14, 7.

Harrell Jr, F.E. & Harrell Jr, M. F. E. 2019. Package ‘hmisc’. CRAN2018, 2019, 235–236.

Herati, R. S., Muselman, A., Vella, L., Bengsch, B., Parkhouse, K., Del Alcazar, D., Kotzin, J., Doyle, S. A., Tebas, P., Hensley, S. E., Su, L. F., Schmader, K. E. & Wherry, E. J. 2017. Successive annual influenza vaccination induces a recurrent oligoclonotypic memory response in circulating T follicular helper cells. Sci Immunol, 2, eaag2152.

Hoffmann, M., Kleine-Weber, H. & Pohlmann, S. 2020. A Multibasic Cleavage Site in the Spike Protein of SARS-CoV-2 Is Essential for Infection of Human Lung Cells. Mol Cell, 78, 779–784 e5.

Howe, K. L., Achuthan, P., Allen, J., Allen, J., Alvarez-Jarreta, J., Amode, M. R., Armean, I. M., Azov, A. G., Bennett, R., Bhai, J., Billis, K., Boddu, S., Charkhchi, M., Cummins, C., Da Rin Fioretto, L., Davidson, C., Dodiya, K., El Houdaigui, B., Fatima, R., Gall, A., Garcia Giron, C., Grego, T., Guijarro-Clarke, C., Haggerty, L., Hemrom, A., Hourlier, T., Izuogu, O. G., Juettemann, T., Kaikala, V., Kay, M., Lavidas, I., Le, T., Lemos, D., Gonzalez Martinez, J., Marugan, J. C., Maurel, T., Mcmahon, A. C., Mohanan, S., Moore, B., Muffato, M., Oheh, D. N., Paraschas, D., Parker, A., Parton, A., Prosovetskaia, I., Sakthivel, M. P., Salam, A. I. A., Schmitt, B. M., Schuilenburg, H., Sheppard, D., Steed, E., Szpak, M., Szuba, M., Taylor, K., Thormann, A., Threadgold, G., Walts, B., Winterbottom, A., Chakiachvili, M., Chaubal, A., De Silva, N., Flint, B., Frankish, A., Hunt, S. E., Gr, I. I., Langridge, N., Loveland, J. E., Martin, F. J., Mudge, J. M., Morales, J., Perry, E., Ruffier, M., Tate, J., Thybert, D., Trevanion, S. J., Cunningham, F., Yates, A. D., Zerbino, D. R. & Flicek, P. 2021. Ensembl 2021. Nucleic Acids Res, 49, D884–D891.

Hsieh, C.-L., Goldsmith, J. A., Schaub, J. M., Divenere, A. M., Kuo, H.-C., Javanmardi, K., Le, K. C., Wrapp, D., Lee, A. G. & Liu, Y. 2020. Structure-based design of prefusion-stabilized SARS-CoV-2 spikes. Science, 369, 1501–1505.

Huynh, K., Barlow, C. K., Jayawardana, K. S., Weir, J. M., Mellett, N. A., Cinel, M., Magliano, D. J., Shaw, J. E., Drew, B. G. & Meikle, P. J. 2019. High-Throughput Plasma Lipidomics: Detailed Mapping of the Associations with Cardiometabolic Risk Factors. Cell Chem Biol, 26, 71–84.e4.

Kalemera, M. D., Cappella-Pujol, J., Chumbe, A., Underwood, A., Bull, R. A., Schinkel, J., Sliepen, K. & Grove, J. 2020. Optimised cell systems for the investigation of hepatitis C virus E1E2 glycoproteins. bioRxiv, 2020.06.18.159442.

Keck, Z. Y., Li, S. H., Xia, J., Von Hahn, T., Balfe, P., Mckeating, J. A., Witteveldt, J., Patel, A. H., Alter, H., Rice, C. M. & Foung, S. K. 2009. Mutations in hepatitis C virus E2 located outside the CD81 binding sites lead to escape from broadly neutralizing antibodies but compromise virus infectivity. J Virol, 83, 6149–60.

Khoo, N. K. H., Lim, J. M. E., Gill, U. S., De Alwis, R., Tan, N., Toh, J. Z. N., Abbott, J. E., Usai, C., Ooi, E. E., Low, J. G. H., Le Bert, N., Kennedy, P. T. F. & Bertoletti, A. 2022. Differential immunogenicity of homologous versus heterologous boost in Ad26.COV2.S vaccine recipients. Med (N Y), 3, 104–118.e4.

Kim, D., Langmead, B. & Salzberg, S. L. 2015. HISAT: a fast spliced aligner with low memory requirements. Nat Methods, 12, 357–60.

Koutsakos, M., Wheatley, A. K., Loh, L., Clemens, E. B., Sant, S., Nussing, S., Fox, A., Chung, A. W., Laurie, K. L., Hurt, A. C., Rockman, S., Lappas, M., Loudovaris, T., Mannering, S. I., Westall, G. P., Elliot, M., Tangye, S. G., Wakim, L. M., Kent, S. J., Nguyen, T. H. O. & Kedzierska, K. 2018. Circulating TFH cells, serological memory, and tissue compartmentalization shape human influenza-specific B cell immunity. Sci Transl Med, 10, eaan8405.

Lee, S. W., Moon, J. Y., Lee, S. K., Lee, H., Moon, S., Chung, S. J., Yeo, Y., Park, T. S., Park, D. W., Kim, T. H., Sohn, J. W., Yoon, H. J. & Kim, S. H. 2021. Anti-SARS-CoV-2 Spike Protein RBD Antibody Levels After Receiving a Second Dose of ChAdOx1 nCov-19 (AZD1222) Vaccine in Healthcare Workers: Lack of Association With Age, Sex, Obesity, and Adverse Reactions. Front Immunol, 12, 779212.

Leek, J. T. 2014. svaseq: removing batch effects and other unwanted noise from sequencing data. Nucleic Acids Res, 42, e161.

Levin, E. G., Lustig, Y., Cohen, C., Fluss, R., Indenbaum, V., Amit, S., Doolman, R., Asraf, K., Mendelson, E., Ziv, A., Rubin, C., Freedman, L., Kreiss, Y. & Regev-Yochay, G. 2021. Waning Immune Humoral Response to BNT162b2 Covid-19 Vaccine over 6 Months. N Engl J Med, 385, e84.

Li, S., Rouphael, N., Duraisingham, S., Romero-Steiner, S., Presnell, S., Davis, C., Schmidt, D. S., Johnson, S. E., Milton, A., Rajam, G., Kasturi, S., Carlone, G. M., Quinn, C., Chaussabel, D., Palucka, A. K., Mulligan, M. J., Ahmed, R., Stephens, D. S., Nakaya, H. I. & Pulendran, B. 2014. Molecular signatures of antibody responses derived from a systems biology study of five human vaccines. Nat Immunol, 15, 195–204.

Liao, Y., Smyth, G. K. & Shi, W. 2014. featureCounts: an efficient general purpose program for assigning sequence reads to genomic features. Bioinformatics, 30, 923–30.

Liu, J., Yu, J., Mcmahan, K., Jacob-Dolan, C., He, X., Giffin, V., Wu, C., Sciacca, M., Powers, O., Nampanya, F., Miller, J., Lifton, M., Hope, D., Hall, K., Hachmann, N. P., Chung, B., Anioke, T., Li, W., Muench, J., Gamblin, A., Boursiquot, M., Cook, A., Lewis, M. G., Andersen, H. & Barouch, D. H. 2022. CD8 T Cells Contribute to Vaccine Protection Against SARS-CoV-2 in Macaques. *Sci Immunol*, eabq7647.

Lönnberg, T., Svensson, V., James, K. R., Fernandez-Ruiz, D., Sebina, I., Montandon, R., Soon, M. S., Fogg, L. G., Nair, A. S., Liligeto, U., Stubbington, M. J., Ly, L. H., Bagger, F. O., Zwiessele, M., Lawrence, N. D., Souza-Fonseca-Guimaraes, F., Bunn, P. T., Engwerda, C. R., Heath, W. R., Billker, O., Stegle, O., Haque, A. & Teichmann, S. A. 2017. Single-cell RNA-seq and computational analysis using temporal mixture modelling resolves Th1/Tfh fate bifurcation in malaria. Sci Immunol, 2, eaal2192.

Loyal, L., Braun, J., Henze, L., Kruse, B., Dingeldey, M., Reimer, U., Kern, F., Schwarz, T., Mangold, M., Unger, C., Dorfler, F., Kadler, S., Rosowski, J., Gurcan, K., Uyar-Aydin, Z., Frentsch, M., Kurth, F., Schnatbaum, K., Eckey, M., Hippenstiel, S., Hocke, A., Muller, M. A., Sawitzki, B., Miltenyi, S., Paul, F., Mall, M. A., Wenschuh, H., Voigt, S., Drosten, C., Lauster, R., Lachman, N., Sander, L. E., Corman, V. M., Rohmel, J., Meyer-Arndt, L., Thiel, A. & Giesecke-Thiel, C. 2021. Cross-reactive CD4(+) T cells enhance SARS-CoV-2 immune responses upon infection and vaccination. Science, 374, eabh1823.

Lustig, Y., Gonen, T., Meltzer, L., Gilboa, M., Indenbaum, V., Cohen, C., Amit, S., Jaber, H., Doolman, R., Asraf, K., Rubin, C., Fluss, R., Mendelson, E., Freedman, L., Regev-Yochay, G. & Kreiss, Y. 2022. Superior immunogenicity and effectiveness of the third compared to the second BNT162b2 vaccine dose. Nat Immunol, 23, 940–946.

Mateus, J., Dan, J. M., Zhang, Z., Rydyznski Moderbacher, C., Lammers, M., Goodwin, B., Sette, A., Crotty, S. & Weiskopf, D. 2021. Low-dose mRNA-1273 COVID-19 vaccine generates durable memory enhanced by cross-reactive T cells. Science, 374, eabj9853.

Mcmahan, K., Yu, J., Mercado, N. B., Loos, C., Tostanoski, L. H., Chandrashekar, A., Liu, J., Peter, L., Atyeo, C., Zhu, A., Bondzie, E. A., Dagotto, G., Gebre, M. S., Jacob-Dolan, C., Li, Z., Nampanya, F., Patel, S., Pessaint, L., Van Ry, A., Blade, K., Yalley-Ogunro, J., Cabus, M., Brown, R., Cook, A., Teow, E., Andersen, H., Lewis, M. G., Lauffenburger, D. A., Alter, G. & Barouch, D. H. 2021. Correlates of protection against SARS-CoV-2 in rhesus macaques. Nature, 590, 630–634.

Mevorach, D., Anis, E., Cedar, N., Bromberg, M., Haas, E. J., Nadir, E., Olsha-Castell, S., Arad, D., Hasin, T., Levi, N., Asleh, R., Amir, O., Meir, K., Cohen, D., Dichtiar, R., Novick, D., Hershkovitz, Y., Dagan, R., Leitersdorf, I., Ben-Ami, R., Miskin, I., Saliba, W., Muhsen, K., Levi, Y., Green, M. S., Keinan-Boker, L. & Alroy-Preis, S. 2021. Myocarditis after BNT162b2 mRNA Vaccine against Covid-19 in Israel. N Engl J Med, 385, 2140–2149.

Michalik, S., Siegerist, F., Palankar, R., Franzke, K., Schindler, M., Reder, A., Seifert, U., Cammann, C., Wesche, J., Steil, L., Hentschker, C., Gesell-Salazar, M., Reisinger, E., Beer, M., Endlich, N., Greinacher, A. & Volker, U. 2022. Comparative analysis of ChAdOx1 nCoV-19 and Ad26.COV2.S SARS-CoV-2 vector vaccines. Haematologica, 107, 947–957.

Nakayamada, S., Kanno, Y., Takahashi, H., Jankovic, D., Lu, K. T., Johnson, T. A., Sun, H. W., Vahedi, G., Hakim, O., Handon, R., Schwartzberg, P. L., Hager, G. L. & O’shea, J. J. 2011. Early Th1 cell differentiation is marked by a Tfh cell-like transition. Immunity, 35, 919–31.

Patone, M., Mei, X. W., Handunnetthi, L., Dixon, S., Zaccardi, F., Shankar-Hari, M., Watkinson, P., Khunti, K., Harnden, A., Coupland, C. A. C., Channon, K. M., Mills, N. L., Sheikh, A. & Hippisley-Cox, J. 2022. Risks of myocarditis, pericarditis, and cardiac arrhythmias associated with COVID-19 vaccination or SARS-CoV-2 infection. Nat Med, 28, 410–422.

Polack, F. P., Thomas, S. J., Kitchin, N., Absalon, J., Gurtman, A., Lockhart, S., Perez, J. L., Perez Marc, G., Moreira, E. D., Zerbini, C., Bailey, R., Swanson, K. A., Roychoudhury, S., Koury, K., Li, P., Kalina, W. V., Cooper, D., Frenck, R. W., Jr., Hammitt, L. L., Tureci, O., Nell, H., Schaefer, A., Unal, S., Tresnan, D. B., Mather, S., Dormitzer, P. R., Sahin, U., Jansen, K. U., Gruber, W. C. & Group, C. C. T. 2020. Safety and Efficacy of the BNT162b2 mRNA Covid-19 Vaccine. N Engl J Med, 383, 2603–2615.

Provine, N. M., Amini, A., Garner, L. C., Spencer, A. J., Dold, C., Hutchings, C., Silva Reyes, L., Fitzpatrick, M. E. B., Chinnakannan, S., Oguti, B., Raymond, M., Ulaszewska, M., Troise, F., Sharpe, H., Morgan, S. B., Hinks, T. S. C., Lambe, T., Capone, S., Folgori, A., Barnes, E., Rollier, C. S., Pollard, A. J. & Klenerman, P. 2021. MAIT cell activation augments adenovirus vector vaccine immunogenicity. Science, 371, 521–526.

Ramasamy, M. N., Minassian, A. M., Ewer, K. J., Flaxman, A. L., Folegatti, P. M., Owens, D. R., Voysey, M., Aley, P. K., Angus, B., Babbage, G., Belij-Rammerstorfer, S., Berry, L., Bibi, S., Bittaye, M., Cathie, K., Chappell, H., Charlton, S., Cicconi, P., Clutterbuck, E. A., Colin-Jones, R., Dold, C., Emary, K. R. W., Fedosyuk, S., Fuskova, M., Gbesemete, D., Green, C., Hallis, B., Hou, M. M., Jenkin, D., Joe, C. C. D., Kelly, E. J., Kerridge, S., Lawrie, A. M., Lelliott, A., Lwin, M. N., Makinson, R., Marchevsky, N. G., Mujadidi, Y., Munro, A. P. S., Pacurar, M., Plested, E., Rand, J., Rawlinson, T., Rhead, S., Robinson, H., Ritchie, A. J., Ross-Russell, A. L., Saich, S., Singh, N., Smith, C. C., Snape, M. D., Song, R., Tarrant, R., Themistocleous, Y., Thomas, K. M., Villafana, T. L., Warren, S. C., Watson, M. E. E., Douglas, A. D., Hill, A. V. S., Lambe, T., Gilbert, S. C., Faust, S. N., Pollard, A. J. & Oxford, C. V. T. G. 2020. Safety and immunogenicity of ChAdOx1 nCoV-19 vaccine administered in a prime-boost regimen in young and old adults (COV002): a single-blind, randomised, controlled, phase 2/3 trial. Lancet, 396, 1979–1993.

Rappsilber, J., Mann, M. & Ishihama, Y. 2007. Protocol for micro-purification, enrichment, pre-fractionation and storage of peptides for proteomics using StageTips. Nat Protoc, 2, 1896–906.

Ritchie, M. E., Phipson, B., Wu, D., Hu, Y., Law, C. W., Shi, W. & Smyth, G. K. 2015. limma powers differential expression analyses for RNA-sequencing and microarray studies. Nucleic Acids Res, 43, e47.

Robinson, M. D., Mccarthy, D. J. & Smyth, G. K. 2010. edgeR: a Bioconductor package for differential expression analysis of digital gene expression data. Bioinformatics, 26, 139–40.

Röltgen, K., Nielsen, S. C. A., Silva, O., Younes, S. F., Zaslavsky, M., Costales, C., Yang, F., Wirz, O. F., Solis, D., Hoh, R. A., Wang, A., Arunachalam, P. S., Colburg, D., Zhao, S., Haraguchi, E., Lee, A. S., Shah, M. M., Manohar, M., Chang, I., Gao, F., Mallajosyula, V., Li, C., Liu, J., Shoura, M. J., Sindher, S. B., Parsons, E., Dashdorj, N. J., Dashdorj, N. D., Monroe, R., Serrano, G. E., Beach, T. G., Chinthrajah, R. S., Charville, G. W., Wilbur, J. L., Wohlstadter, J. N., Davis, M. M., Pulendran, B., Troxell, M. L., Sigal, G. B., Natkunam, Y., Pinsky, B. A., Nadeau, K. C. & Boyd, S. D. 2022. Immune imprinting, breadth of variant recognition, and germinal center response in human SARS-CoV-2 infection and vaccination. Cell, 185, 1025–1040.e14.

Saggau, C., Martini, G. R., Rosati, E., Meise, S., Messner, B., Kamps, A.-K., Bekel, N., Gigla, J., Rose, R., Voß, M., Geisen, U. M., Reid, H. M., Sümbül, M., Tran, F., Berner, D. K., Khodamoradi, Y., Vehreschild, M. J. G. T., Cornely, O., Koehler, P., Krumbholz, A., Fickenscher, H., Kreuzer, O., Schreiber, C., Franke, A., Schreiber, S., Hoyer, B., Scheffold, A. & Bacher, P. 2022. The pre-exposure SARS-CoV-2-specific T cell repertoire determines the quality of the immune response to vaccination. Immunity.

Sammicheli, S., Kuka, M., Di Lucia, P., De Oya, N. J., De Giovanni, M., Fioravanti, J., Cristofani, C., Maganuco, C. G., Fallet, B., Ganzer, L., Sironi, L., Mainetti, M., Ostuni, R., Larimore, K., Greenberg, P. D., De La Torre, J. C., Guidotti, L. G. & Iannacone, M. 2016. Inflammatory monocytes hinder antiviral B cell responses. Sci Immunol, 1, eaah6789.

Sanz, I., Wei, C., Jenks, S. A., Cashman, K. S., Tipton, C., Woodruff, M. C., Hom, J. & Lee, F. E. 2019. Challenges and Opportunities for Consistent Classification of Human B Cell and Plasma Cell Populations. Front Immunol, 10, 2458.

Schultz, N. H., Sorvoll, I. H., Michelsen, A. E., Munthe, L. A., Lund-Johansen, F., Ahlen, M. T., Wiedmann, M., Aamodt, A. H., Skattor, T. H., Tjonnfjord, G. E. & Holme, P. A. 2021. Thrombosis and Thrombocytopenia after ChAdOx1 nCoV-19 Vaccination. N Engl J Med, 384, 2124–2130.

Scully, M., Singh, D., Lown, R., Poles, A., Solomon, T., Levi, M., Goldblatt, D., Kotoucek, P., Thomas, W. & Lester, W. 2021. Pathologic Antibodies to Platelet Factor 4 after ChAdOx1 nCoV-19 Vaccination. N Engl J Med, 384, 2202–2211.

Simpson, C. R., Shi, T., Vasileiou, E., Katikireddi, S. V., Kerr, S., Moore, E., Mccowan, C., Agrawal, U., Shah, S. A., Ritchie, L. D., Murray, J., Pan, J., Bradley, D. T., Stock, S. J., Wood, R., Chuter, A., Beggs, J., Stagg, H. R., Joy, M., Tsang, R. S. M., De Lusignan, S., Hobbs, R., Lyons, R. A., Torabi, F., Bedston, S., O’leary, M., Akbari, A., Mcmenamin, J., Robertson, C. & Sheikh, A. 2021. First-dose ChAdOx1 and BNT162b2 COVID-19 vaccines and thrombocytopenic, thromboembolic and hemorrhagic events in Scotland. Nat Med, 27, 1290–1297.

Takano, T., Morikawa, M., Adachi, Y., Kabasawa, K., Sax, N., Moriyama, S., Sun, L., Isogawa, M., Nishiyama, A., Onodera, T., Terahara, K., Tonouchi, K., Nishimura, M., Tomii, K., Yamashita, K., Matsumura, T., Shinkai, M. & Takahashi, Y. 2022. Distinct immune cell dynamics correlate with the immunogenicity and reactogenicity of SARS-CoV-2 mRNA vaccine. Cell Rep Med, 3, 100631.

Tarke, A., Coelho, C. H., Zhang, Z., Dan, J. M., Yu, E. D., Methot, N., Bloom, N. I., Goodwin, B., Phillips, E., Mallal, S., Sidney, J., Filaci, G., Weiskopf, D., Da Silva Antunes, R., Crotty, S., Grifoni, A. & Sette, A. 2022. SARS-CoV-2 vaccination induces immunological T cell memory able to cross-recognize variants from Alpha to Omicron. Cell, 185, 847–859.e11.

Therapeutic Goods Administration, A. D. O. H. 2022. COVID-19 vaccine adverse events [Online]. Australian Department of Health. Available: https://www.health.gov.au/initiatives-and-programs/covid-19-vaccines/advice-for-providers/clinical-guidance/adverse-events [Accessed 24/05/2022 2022].

Tsang, J. S., Schwartzberg, P. L., Kotliarov, Y., Biancotto, A., Xie, Z., Germain, R. N., Wang, E., Olnes, M. J., Narayanan, M., Golding, H., Moir, S., Dickler, H. B., Perl, S. & Cheung, F. 2014. Global analyses of human immune variation reveal baseline predictors of postvaccination responses. Cell, 157, 499–513.

Tseng, H. F., Ackerson, B. K., Luo, Y., Sy, L. S., Talarico, C. A., Tian, Y., Bruxvoort, K. J., Tubert, J. E., Florea, A., Ku, J. H., Lee, G. S., Choi, S. K., Takhar, H. S., Aragones, M. & Qian, L. 2022. Effectiveness of mRNA-1273 against SARS-CoV-2 Omicron and Delta variants. Nat Med, 28, 1063–1071.

Vanderlugt, C. L. & Miller, S. D. 2002. Epitope spreading in immune-mediated diseases: implications for immunotherapy. Nature Reviews Immunology, 2, 85–95.

Voysey, M., Clemens, S. A. C., Madhi, S. A., Weckx, L. Y., Folegatti, P. M., Aley, P. K., Angus, B., Baillie, V. L., Barnabas, S. L., Bhorat, Q. E., Bibi, S., Briner, C., Cicconi, P., Collins, A. M., Colin-Jones, R., Cutland, C. L., Darton, T. C., Dheda, K., Duncan, C. J. A., Emary, K. R. W., Ewer, K. J., Fairlie, L., Faust, S. N., Feng, S., Ferreira, D. M., Finn, A., Goodman, A. L., Green, C. M., Green, C. A., Heath, P. T., Hill, C., Hill, H., Hirsch, I., Hodgson, S. H. C., Izu, A., Jackson, S., Jenkin, D., Joe, C. C. D., Kerridge, S., Koen, A., Kwatra, G., Lazarus, R., Lawrie, A. M., Lelliott, A., Libri, V., Lillie, P. J., Mallory, R., Mendes, A. V. A., Milan, E. P., Minassian, A. M., Mcgregor, A., Morrison, H., Mujadidi, Y. F., Nana, A., O’reilly, P. J., Padayachee, S. D., Pittella, A., Plested, E., Pollock, K. M., Ramasamy, M. N., Rhead, S., Schwarzbold, A. V., Singh, N., Smith, A., Song, R., Snape, M. D., Sprinz, E., Sutherland, R. K., Tarrant, R., Thomson, E. C., Torok, M. E., Toshner, M., Turner, D. P. J., Vekemans, J., Villafana, T. L., Watson, M. E. E., Williams, C. J., Douglas, A. D., Hill, A. V. S., Lambe, T., Gilbert, S. C., Pollard, A. J. & Oxford, C. V. T. G. 2021. Safety and efficacy of the ChAdOx1 nCoV-19 vaccine (AZD1222) against SARS-CoV-2: an interim analysis of four randomised controlled trials in Brazil, South Africa, and the UK. Lancet, 397, 99–111.

Watson, O. J., Barnsley, G., Toor, J., Hogan, A. B., Winskill, P. & Ghani, A. C. 2022. Global impact of the first year of COVID-19 vaccination: a mathematical modelling study. Lancet Infect Dis.

Wickham, H. 2016. Package ‘ggplot2’: elegant graphics for data analysis. Springer-Verlag New York. doi, 10, 978-0.

Yoon, S. K., Hegmann, K. T., Thiese, M. S., Burgess, J. L., Ellingson, K., Lutrick, K., Olsho, L. E. W., Edwards, L. J., Sokol, B., Caban-Martinez, A. J., Schaefer-Solle, N., Jones, J. M., Tyner, H., Hunt, A., Respet, K., Gaglani, M., Dunnigan, K., Rose, S., Naleway, A., Groom, H., Kuntz, J., Fowlkes, A. L., Thompson, M. G., Yoo, Y. M., Investigators, H.-R. N. & Investigators, H.-R. N. 2022. Protection with a Third Dose of mRNA Vaccine against SARS-CoV-2 Variants in Frontline Workers. N Engl J Med, 386, 1855–1857.

Zhang, Z., Mateus, J., Coelho, C. H., Dan, J. M., Moderbacher, C. R., Galvez, R. I., Cortes, F. H., Grifoni, A., Tarke, A., Chang, J., Escarrega, E. A., Kim, C., Goodwin, B., Bloom, N. I., Frazier, A., Weiskopf, D., Sette, A. & Crotty, S. 2022. Humoral and cellular immune memory to four COVID-19 vaccines. Cell, 185, 2434–2451 e17.

