## Supplementary file 1 for "A systems immunology study comparing innate and adaptive immune responses in adults to COVID-19 mRNA (BNT162b2/mRNA-1273) and adenovirus vectored vaccines (ChAdOx1-S) after the first, second and third doses"

**REDCap Surveys**

**COVIRS Survey 1 (1 week post 1st vaccination date)**

Thank you for providing a blood sample in the COVIRS study. The main purpose of this survey is to confirm the date you received DOSE 1 of the COVID-19-specific vaccine and to confirm which COVID-19-specific vaccine you had. In addition, we will ask about any reactions you may have had to the vaccine. The second purpose is to collect information about episodes of COVID-19 you may have had since you had your COVID-19-specific vaccine.

Date survey 1 completed: ___________________________________

Details about DOSE 1 of the COVID-19-specific vaccination

You have previously confirmed having DOSE 1 of the following COVID-19 specific vaccination:

Vaccine: [bc_covac_which_01] on [bc_covac_date_01].

Are these details correct?

- Yes
- No, I have not received a COVID-19 specific vaccination as yet
- No, the date or type of vaccination is incorrect

Do you have an expected date for your second COVID-19 specific vaccine?
___________________________________ (Leave blank if you do not know)

Are you booked in to receive a COVID-19 specific vaccination?

- Yes
- No

Please tell us your booking date: ___________________________________

Which COVID-19-specific vaccine are you booked to receive?

- AstraZeneca/Oxford (ChAdOx1, Covishield)
- Pfizer/BioNTech (BNT162b2, Comirnaty)
- Moderna (mRNA-1273)
- Sinovac (CoronaVac)
- Novavax (NVX-CoV2373)
- Johnson and Johnson (Ad26.COV2.S)
- Gam-Covid-Vac (Sputnik V)
- Other

Adverse events after DOSE 1 of the COVID-19-specific vaccine

Did you experience any adverse event up to seven days after receiving DOSE 1 of the COVID-19-specific vaccine?

- None
- Pain at the vaccination site
- Redness at the vaccination site
- Swelling at the vaccination site
- Tenderness at the vaccination site
- Itchiness at the vaccination site
- Lymph node enlargement in region draining the vaccination site
- Fever
- Chills
- Fatigue
- Headache
- Nausea and/or vomiting
- Diarrhoea
- Muscle pain
- Joint pain

On what day after vaccination did the pain start? At day number: ___________________________________ (NB: Day number 1 is the day you received your vaccination)

For how many days did the pain last? ___________________________________ (Days)

On what day after vaccination did the redness start? At day number: ___________________________________ (NB: Day number 1 is the day you received your vaccination)

For how many days did the redness last? ___________________________________ (Days)

What was the largest diameter of the redness, at its worst (in cm)? ___________________________________ (Please provide answer in cm)

On what day after vaccination did the swelling start? At day number: ___________________________________ (NB: Day number 1 is the day you received your vaccination)

For how many days did the swelling last? ___________________________________ (Days)

What was the largest diameter of the swelling at its worst (in cm)?
___________________________________ (Please provide answer in cm)

On what day after vaccination did the tenderness start? At day number:
___________________________________ (Note: Day number 1 is the day you received your vaccination)

For how many days did the tenderness last? ___________________________________ (Days)

Regarding the level of tenderness only: How would you describe the level of discomfort at its worst?

- Mild discomfort to touch
- Discomfort with movement
- Significant discomfort at rest

Did the pain/tenderness and/or swelling at the vaccination site significantly interfere with your daily activities?

- It did not significantly interfere with my daily activities
- It somewhat interfered with my daily activities
- It prevented me from doing my daily activities

Please describe how the pain/tenderness and/or swelling interfered, and for how long:

___________________________________________

On what day after vaccination did the itchiness start? At day number:
___________________________________ (Note: Day number 1 is the day you received your vaccination)

For how many days did the itchiness last? ___________________________________ (Days)

Where was it itchy?

- Only my vaccination site felt itchy
- The itching extended beyond my vaccination site, but not all over my body
- I was itchy all over my body

Did you have to use medication for the itch?

- I did not need to take any medication
- I had to use antihistamine (e.g. Zyrtec, Claratyne, Telfast) for less than 48 hours
- I had to use antihistamine (e.g. Zyrtec, Claratyne, Telfast) for 48 hours or longer

- Other

If other, please tell us where: ___________________________________

On what day after vaccination did the lymph node enlargement start? At day number: ___________________________________ (Note: Day number 1 is the day you received your vaccination)

For how many days did the lymph node enlargement last? ___________________________________ (Days)

Where have you noticed or felt a swollen gland?

- Under the armpit
- In the neck
- Other

If other, please tell us where: ___________________________________

How big was the swollen gland (in cm) under the armpit? ___________________________________ (Please provide answer in cm)

How big was the swollen gland (in cm) in the neck? ___________________________________ (Please provide answer in cm)

How big was the swollen gland (in cm) in another location? ___________________________________ (Please provide answer in cm)

On what day after vaccination did the fever start? At day number: ___________________________________ (Note: Day number 1 is the day you received your vaccination)

For how many days did the fever last? ___________________________________ (Days)

What was your maximum temperature? ___________________________________

On what day after vaccination did the chills start? At day number: ___________________________________ (Note: Day number 1 is the day you received your vaccination)

For how many days did the chills last? ___________________________________ (Days)

On what day after vaccination did the fatigue start? At day number: ___________________________________ (Note: Day number 1 is the day you received your vaccination)

For how many days did the fatigue last? ___________________________________ (Days)

Did the fatigue significantly interfere with your daily activities?

- It did not significantly interfere with my daily activities
- It somewhat interfered with my daily activities
- It prevented me from doing my daily activities

Please describe how the fatigue interfered, and for how long:

___________________________________________

On what day after vaccination did the headache start? At day number: ___________________________________ (Note: Day number 1 is the day you received your vaccination)

For how many days did the headache last? ___________________________________ (Days)

Did the headache significantly interfere with your daily activities?

- It did not significantly interfere with my daily activities
- It somewhat interfered with my daily activities
- It prevented me from doing my daily activities

Please describe how the headache interfered, and for how long:

___________________________________________

On what day after vaccination did the nausea and/or vomiting start? At day number: ___________________________________ (Note: Day number 1 is the day you received your vaccination)

For how many days did the nausea and/or vomiting last? ___________________________________ (Days)

How many episodes per day did you have of vomiting at its worst?

___________________________________

Did the nausea and/or vomiting significantly interfere with your daily activities?

- It did not significantly interfere with my daily activities
- It somewhat interfered with my daily activities

- It prevented me from doing my daily activities

Please describe how the nausea and/or vomiting interfered, and for how long:

___________________________________________

On what day after vaccination did the diarrhoea start? At day number:
___________________________________ (Note: Day number 1 is the day you received your vaccination)

For how many days did the diarrhoea last? ___________________________________ (Days)

How many episodes per day did you have of diarrhoea at its worst?

___________________________________

On what day after vaccination did the muscle pain start? At day number:
___________________________________ (Note: Day number 1 is the day you received your vaccination)

For how many days did the muscle pain last? ___________________________________ (Days)

On what day after vaccination did the joint pain start? At day number:
___________________________________ (Note: Day number 1 is the day you received your vaccination) For how many days did the joint pain last? ___________________________________ (Days)

Did the muscle and/or joint pain significantly interfere with your daily activities?

- It did not significantly interfere with my daily activities
- It somewhat interfered with my daily activities
- It prevented me from doing my daily activities

Please describe how the muscle and/or joint pain interfered, and for how long:

___________________________________________

Did you have to use medication or consult a medical doctor?

I did not need to take any medication, nor see a medical doctor

I had to consult a medical doctor or be hospitalised

I had to use pain medication

Please describe when you saw the doctor, and what was discussed:

___________________________________________

Which medication did you take? ___________________________________

For how many days did you use this medication? ___________________________________ (Days)

Allergic reactions after DOSE 1 of the COVID-19-specific vaccine

Did you have an allergic reaction after DOSE 1 of the vaccination? Please select all that apply:

- None
- Urticaria (hives) or cutaneous rash
- Runny or stuffy nose and sneezing
- Vomiting, diarrhea, or abdominal cramps
- Swollen or itchy lips or tongue
- Swollen or itchy throat, hoarse voice, trouble swallowing, tightness in your throat
- Coughing, wheezing, shortness of breath
- Fainting, dizziness, confusion, or weakness
- Other

Other allergic reaction, please describe: ___________________________________

How long after vaccination (in minutes) did the allergic reaction start?

___________________________________

What treatment did you receive? Please select all that apply:

- No treatment
- Anti-histamine
- Adrenaline Inhaler
- Prednisolone or other steroids
- Transferred to the Emergency department
- Hospitalisation in normal unit (non-ICU)
- Hospitalisation in Intensive Care Unit (ICU)
- Other

If other, please specify: ___________________________________

Please describe what happened: ___________________________________________

Other vaccinations

Did you receive any other vaccines since the start of COVIRS?

- Yes
- No

If yes, which vaccine(s) did you receive?

- Diphtheria-tetanus vaccine (ADT Booster)
- Diphtheria-tetanus-pertussis vaccine (Boostrix, Adacel, Tripacel)
- Diphtheria-tetanus-pertussis-polio vaccine (Boostrix-IPV, Adacel Polio, Quadracel)
- Polio vaccine (IPOL)
- Hepatitis B vaccine (Engerix-B, H-B-Vax II)
- Hepatitis A vaccine (Havrix, Avaxim, Vaqta)
- Hepatitis A-hepatitis B vaccine (Twinrix)
- Hepatitis A-typhoid vaccine (Vivaxim)
- Typhoid injected vaccine (Typhim Vi)
- Typhoid oral vaccine (Vivotif Oral)
- Influenza vaccine (Afluria, Fluad Quad, Fluarix, FluQuadri, Influvac, Vaxigrip, Vaxigroup)
- Papillomavirus vaccine (Cervarix, Gardasil)
- Meningococcal vaccine (Menveo, Menactra, MenQuadfi, NeisVac, Bexsero, Trumenba)
- Pneumococcal vaccine (Prevenar, Synflorix, Pneumosil, Pneumovax)
- Japanese encephalitis vaccine (Imojev, JEspect)
- Rabies vaccine (Rabipur)
- Yellow fever vaccine (Stamaril)
- Measles-mumps-rubella (Priorix, M-M-R II, ProQuad)
- Measles-mumps-rubella-varicella (Priorix-tetra, ProQuad)
- Varicella vaccine (Varilrix, Varivax)
- Zoster live vaccine (Zostavaq)
- Zoster non-live vaccine (Shingrix)
- Tuberculosis vaccine (BCG)
- Other

If other, please specify: ___________________________________

**COVIRS Survey 2 (1 week post 2$^{nd}$ vaccination date)**

Thank you for providing a blood sample in the COVIRS study. The main purpose of this survey is to confirm the date you received DOSE 2 of the COVID-19-specific vaccine and to confirm which COVID-19-specific vaccine you had. In addition, we will ask about any reactions you may have had to the vaccine. The second purpose is to collect information about episodes of COVID-19 you may have had since you had your COVID-19-specific vaccine.

Date survey 2 completed: ___________________________________

Details about DOSE 2 of the COVID-19-specific vaccination

You have previously confirmed having DOSE 2 of the following COVID-19 specific vaccination:

Vaccine: [bc_covac_which_01] on [bc_covac_date_01].

Are these details correct?

- Yes
- No, I have not received a COVID-19 specific vaccination as yet
- No, the date or type of vaccination is incorrect

Adverse events after DOSE 2 of the COVID-19-specific vaccine

Did you experience any adverse event up to seven days after receiving DOSE 1 of the COVID-19-specific vaccine?

- None
- Pain at the vaccination site
- Redness at the vaccination site
- Swelling at the vaccination site
- Tenderness at the vaccination site
- Itchiness at the vaccination site
- Lymph node enlargement in region draining the vaccination site
- Fever
- Chills
- Fatigue
- Headache
- Nausea and/or vomiting
- Diarrhoea
- Muscle pain
- Joint pain

On what day after vaccination did the pain start? At day number: ____________________________________ (NB: Day number 1 is the day you received your vaccination)

For how many days did the pain last? ____________________________________ (Days)

On what day after vaccination did the redness start? At day number:
___________________________________ (NB: Day number 1 is the day you received your vaccination)

For how many days did the redness last? ___________________________________ (Days)

What was the largest diameter of the redness, at its worst (in cm)?
___________________________________ (Please provide answer in cm)

On what day after vaccination did the swelling start? At day number:
___________________________________ (NB: Day number 1 is the day you received your vaccination)

For how many days did the swelling last? ___________________________________ (Days)

What was the largest diameter of the swelling at its worst (in cm)?
___________________________________ (Please provide answer in cm)

On what day after vaccination did the tenderness start? At day number:
___________________________________ (Note: Day number 1 is the day you received your vaccination)

For how many days did the tenderness last? ___________________________________ (Days)

Regarding the level of tenderness only: How would you describe the level of discomfort at its worst?

- Mild discomfort to touch
- Discomfort with movement
- Significant discomfort at rest

Did the pain/tenderness and/or swelling at the vaccination site significantly interfere with your daily activities?

- It did not significantly interfere with my daily activities
- It somewhat interfered with my daily activities
- It prevented me from doing my daily activities

Please describe how the pain/tenderness and/or swelling interfered, and for how long:

___________________________________________

On what day after vaccination did the itchiness start? At day number:
___________________________________ (Note: Day number 1 is the day you received your vaccination)

For how many days did the itchiness last? ___________________________________ (Days)

Where was it itchy?

- Only my vaccination site felt itchy
- The itching extended beyond my vaccination site, but not all over my body
- I was itchy all over my body

Did you have to use medication for the itch?

- I did not need to take any medication
- I had to use antihistamine (e.g. Zyrtec, Claratyne, Telfast) for less than 48 hours
- I had to use antihistamine (e.g. Zyrtec, Claratyne, Telfast) for 48 hours or longer
- Other

If other, please tell us where: ___________________________________

On what day after vaccination did the lymph node enlargement start? At day number: ___________________________________ (Note: Day number 1 is the day you received your vaccination)

For how many days did the lymph node enlargement last? ___________________________________ (Days)

Where have you noticed or felt a swollen gland?

- Under the armpit
- In the neck
- Other

If other, please tell us where: ___________________________________

How big was the swollen gland (in cm) under the armpit? ___________________________________ (Please provide answer in cm)

How big was the swollen gland (in cm) in the neck? ___________________________________ (Please provide answer in cm)

How big was the swollen gland (in cm) in another location? ___________________________________ (Please provide answer in cm)

On what day after vaccination did the fever start? At day number: ___________________________________ (Note: Day number 1 is the day you received your vaccination)

For how many days did the fever last? ___________________________________ (Days)

What was your maximum temperature? ___________________________________

On what day after vaccination did the chills start? At day number: ___________________________________ (Note: Day number 1 is the day you received your vaccination)

For how many days did the chills last? ___________________________________ (Days)

On what day after vaccination did the fatigue start? At day number: ___________________________________ (Note: Day number 1 is the day you received your vaccination)

For how many days did the fatigue last? ___________________________________ (Days)

Did the fatigue significantly interfere with your daily activities?

- It did not significantly interfere with my daily activities
- It somewhat interfered with my daily activities
- It prevented me from doing my daily activities

Please describe how the fatigue interfered, and for how long:

___________________________________________

On what day after vaccination did the headache start? At day number: ___________________________________ (Note: Day number 1 is the day you received your vaccination)

For how many days did the headache last? ___________________________________ (Days)

Did the headache significantly interfere with your daily activities?

- It did not significantly interfere with my daily activities
- It somewhat interfered with my daily activities
- It prevented me from doing my daily activities

Please describe how the headache interfered, and for how long:

___________________________________________

On what day after vaccination did the nausea and/or vomiting start? At day number: ___________________________________ (Note: Day number 1 is the day you received your vaccination)

For how many days did the nausea and/or vomiting last? ___________________________________ (Days)

How many episodes per day did you have of vomiting at its worst?

___________________________________

Did the nausea and/or vomiting significantly interfere with your daily activities?

- It did not significantly interfere with my daily activities
- It somewhat interfered with my daily activities
- It prevented me from doing my daily activities

Please describe how the nausea and/or vomiting interfered, and for how long:

___________________________________________

On what day after vaccination did the diarrhoea start? At day number: ___________________________________ (Note: Day number 1 is the day you received your vaccination)

For how many days did the diarrhoea last? ___________________________________ (Days)

How many episodes per day did you have of diarrhoea at its worst?

___________________________________

On what day after vaccination did the muscle pain start? At day number: ___________________________________ (Note: Day number 1 is the day you received your vaccination)

For how many days did the muscle pain last? ___________________________________ (Days)

On what day after vaccination did the joint pain start? At day number: ___________________________________ (Note: Day number 1 is the day you received your vaccination) For how many days did the joint pain last? ___________________________________ (Days)

Did the muscle and/or joint pain significantly interfere with your daily activities?

- It did not significantly interfere with my daily activities
- It somewhat interfered with my daily activities

- It prevented me from doing my daily activities

Please describe how the muscle and/or joint pain interfered, and for how long:

_____________________________________________

Did you have to use medication or consult a medical doctor?

I did not need to take any medication, nor see a medical doctor

I had to consult a medical doctor or be hospitalised

I had to use pain medication

Please describe when you saw the doctor, and what was discussed:

_____________________________________________

Which medication did you take? ____________________________________

For how many days did you use this medication? ____________________________________ (Days)

Allergic reactions after DOSE 2 of the COVID-19-specific vaccine

Did you have an allergic reaction after DOSE 1 of the vaccination? Please select all that apply:

- None
- Urticaria (hives) or cutaneous rash
- Runny or stuffy nose and sneezing
- Vomiting, diarrhea, or abdominal cramps
- Swollen or itchy lips or tongue
- Swollen or itchy throat, hoarse voice, trouble swallowing, tightness in your throat
- Coughing, wheezing, shortness of breath
- Fainting, dizziness, confusion, or weakness
- Other

Other allergic reaction, please describe: ____________________________________

How long after vaccination (in minutes) did the allergic reaction start?

____________________________________

What treatment did you receive? Please select all that apply:

- No treatment
- Anti-histamine
- Adrenaline Inhaler
- Prednisolone or other steroids

- Transferred to the Emergency department
- Hospitalisation in normal unit (non-ICU)
- Hospitalisation in Intensive Care Unit (ICU)
- Other

If other, please specify: ___________________________________

Please describe what happened: ___________________________________________

<u>Other vaccinations</u>

Did you receive any other vaccines since the start of COVIRS?

- Yes
- No

If yes, which vaccine(s) did you receive?

- Diphtheria-tetanus vaccine (ADT Booster)
- Diphtheria-tetanus-pertussis vaccine (Boostrix, Adacel, Tripacel)
- Diphtheria-tetanus-pertussis-polio vaccine (Boostrix-IPV, Adacel Polio, Quadracel)
- Polio vaccine (IPOL)
- Hepatitis B vaccine (Engerix-B, H-B-Vax II)
- Hepatitis A vaccine (Havrix, Avaxim, Vaqta)
- Hepatitis A-hepatitis B vaccine (Twinrix)
- Hepatitis A-typhoid vaccine (Vivaxim)
- Typhoid injected vaccine (Typhim Vi)
- Typhoid oral vaccine (Vivotif Oral)
- Influenza vaccine (Afluria, Fluad Quad, Fluarix, FluQuadri, Influvac, Vaxigrip, Vaxigroup)
- Papillomavirus vaccine (Cervarix, Gardasil)
- Meningococcal vaccine (Menveo, Menactra, MenQuadfi, NeisVac, Bexsero, Trumenba)
- Pneumococcal vaccine (Prevenar, Synflorix, Pneumosil, Pneumovax)
- Japanese encephalitis vaccine (Imojev, JEspect)
- Rabies vaccine (Rabipur)
- Yellow fever vaccine (Stamaril)
- Measles-mumps-rubella (Priorix, M-M-R II, ProQuad)
- Measles-mumps-rubella-varicella (Priorix-tetra, ProQuad)
- Varicella vaccine (Varilrix, Varivax)
- Zoster live vaccine (Zostavaq)
- Zoster non-live vaccine (Shingrix)
- Tuberculosis vaccine (BCG)
- Other

If other, please specify: ___________________________________

**COVIRS Survey 3 (1 week post 3rd booster vaccination date)**

Thank you for providing a blood sample in the COVIRS study. The main purpose of this survey is to confirm the date you received DOSE 3 of the COVID-19-specific vaccine and to confirm which COVID-19-specific vaccine you had. In addition, we will ask about any reactions you may have had to the vaccine. The second purpose is to collect information about episodes of COVID-19 you may have had since you had your COVID-19-specific vaccine.

Date survey 3 completed: ___________________________________

<u>Details about DOSE 3 of the COVID-19-specific vaccination</u>

You have previously confirmed having DOSE 3 on [bc_covac_date_01].

Are these details correct?

- Yes
- No, the date is incorrect

Which COVID-19-specific vaccine did you receive?

- AstraZeneca/Oxford (ChAdOx1, Covishield)
- Pfizer/BioNTech (BNT162b2, Comirnaty)
- Moderna (mRNA-1273)
- Sinovac (CoronaVac)
- Novavax (NVX-CoV2373)
- Johnson and Johnson (Ad26.COV2.S)
- Gam-Covid-Vac (Sputnik V)
- Other

Have you had confirmed COVID-19 infection?

- Yes
- No

If yes, what date were you diagnosed? _____________________

How many days were you unwell? _____________________

Adverse events after DOSE 3 of the COVID-19-specific vaccine

Did you experience any adverse event up to seven days after receiving DOSE 3 of the COVID-19-specific vaccine?

- None
- Pain at the vaccination site
- Redness at the vaccination site
- Swelling at the vaccination site
- Tenderness at the vaccination site
- Itchiness at the vaccination site
- Lymph node enlargement in region draining the vaccination site
- Fever
- Chills
- Fatigue
- Headache
- Nausea and/or vomiting
- Diarrhoea
- Muscle pain
- Joint pain

On what day after vaccination did the pain start? At day number: ____________________________________ (NB: Day number 1 is the day you received your vaccination)

For how many days did the pain last? ____________________________________ (Days)

On what day after vaccination did the redness start? At day number: ____________________________________ (NB: Day number 1 is the day you received your vaccination)

For how many days did the redness last? ____________________________________ (Days)

What was the largest diameter of the redness, at its worst (in cm)? ____________________________________ (Please provide answer in cm)

On what day after vaccination did the swelling start? At day number: ____________________________________ (NB: Day number 1 is the day you received your vaccination)

For how many days did the swelling last? ____________________________________ (Days)

What was the largest diameter of the swelling at its worst (in cm)?
___________________________________ (Please provide answer in cm)

On what day after vaccination did the tenderness start? At day number:
___________________________________ (Note: Day number 1 is the day you received your vaccination)

For how many days did the tenderness last? ___________________________________ (Days)

Regarding the level of tenderness only: How would you describe the level of discomfort at its worst?

- Mild discomfort to touch
- Discomfort with movement
- Significant discomfort at rest

Did the pain/tenderness and/or swelling at the vaccination site significantly interfere with your daily activities?

- It did not significantly interfere with my daily activities
- It somewhat interfered with my daily activities
- It prevented me from doing my daily activities

Please describe how the pain/tenderness and/or swelling interfered, and for how long:

___________________________________________

On what day after vaccination did the itchiness start? At day number:
___________________________________ (Note: Day number 1 is the day you received your vaccination)

For how many days did the itchiness last? ___________________________________ (Days)

Where was it itchy?

- Only my vaccination site felt itchy
- The itching extended beyond my vaccination site, but not all over my body
- I was itchy all over my body

Did you have to use medication for the itch?

- I did not need to take any medication
- I had to use antihistamine (e.g. Zyrtec, Claratyne, Telfast) for less than 48 hours
- I had to use antihistamine (e.g. Zyrtec, Claratyne, Telfast) for 48 hours or longer
- Other

If other, please tell us where: ___________________________________

On what day after vaccination did the lymph node enlargement start? At day number: ___________________________________ (Note: Day number 1 is the day you received your vaccination)

For how many days did the lymph node enlargement last? ___________________________________ (Days)

Where have you noticed or felt a swollen gland?

- Under the armpit
- In the neck
- Other

If other, please tell us where: ___________________________________

How big was the swollen gland (in cm) under the armpit? ___________________________________ (Please provide answer in cm)

How big was the swollen gland (in cm) in the neck? ___________________________________ (Please provide answer in cm)

How big was the swollen gland (in cm) in another location? ___________________________________ (Please provide answer in cm)

On what day after vaccination did the fever start? At day number: ___________________________________ (Note: Day number 1 is the day you received your vaccination)

For how many days did the fever last? ___________________________________ (Days)

What was your maximum temperature? ___________________________________

On what day after vaccination did the chills start? At day number: ___________________________________ (Note: Day number 1 is the day you received your vaccination)

For how many days did the chills last? ___________________________________ (Days)

On what day after vaccination did the fatigue start? At day number:
___________________________________ (Note: Day number 1 is the day you received your vaccination)

For how many days did the fatigue last? ___________________________________ (Days)

Did the fatigue significantly interfere with your daily activities?

- It did not significantly interfere with my daily activities
- It somewhat interfered with my daily activities
- It prevented me from doing my daily activities

Please describe how the fatigue interfered, and for how long:
___________________________________________

On what day after vaccination did the headache start? At day number:
___________________________________ (Note: Day number 1 is the day you received your vaccination)

For how many days did the headache last? ___________________________________ (Days)

Did the headache significantly interfere with your daily activities?

- It did not significantly interfere with my daily activities
- It somewhat interfered with my daily activities
- It prevented me from doing my daily activities

Please describe how the headache interfered, and for how long:
___________________________________________

On what day after vaccination did the nausea and/or vomiting start? At day number:
___________________________________ (Note: Day number 1 is the day you received your vaccination)

For how many days did the nausea and/or vomiting last? ___________________________________ (Days)

How many episodes per day did you have of vomiting at its worst?
___________________________________

Did the nausea and/or vomiting significantly interfere with your daily activities?

- It did not significantly interfere with my daily activities
- It somewhat interfered with my daily activities

- It prevented me from doing my daily activities

Please describe how the nausea and/or vomiting interfered, and for how long:
____________________________________________

On what day after vaccination did the diarrhoea start? At day number:
____________________________________ (Note: Day number 1 is the day you received your vaccination)

For how many days did the diarrhoea last? ____________________________________ (Days)

How many episodes per day did you have of diarrhoea at its worst?
____________________________________

On what day after vaccination did the muscle pain start? At day number:
____________________________________ (Note: Day number 1 is the day you received your vaccination)

For how many days did the muscle pain last? ____________________________________ (Days)

On what day after vaccination did the joint pain start? At day number:
____________________________________ (Note: Day number 1 is the day you received your vaccination) For how many days did the joint pain last? ____________________________________ (Days)

Did the muscle and/or joint pain significantly interfere with your daily activities?

- It did not significantly interfere with my daily activities
- It somewhat interfered with my daily activities
- It prevented me from doing my daily activities

Please describe how the muscle and/or joint pain interfered, and for how long:
____________________________________________

Did you have to use medication or consult a medical doctor?

I did not need to take any medication, nor see a medical doctor

I had to consult a medical doctor or be hospitalised

I had to use pain medication

Please describe when you saw the doctor, and what was discussed:
____________________________________________

Which medication did you take? ________________________________

For how many days did you use this medication? ________________________________ (Days)

Allergic reactions after DOSE 2 of the COVID-19-specific vaccine

Did you have an allergic reaction after DOSE 1 of the vaccination? Please select all that apply:

- None
- Urticaria (hives) or cutaneous rash
- Runny or stuffy nose and sneezing
- Vomiting, diarrhea, or abdominal cramps
- Swollen or itchy lips or tongue
- Swollen or itchy throat, hoarse voice, trouble swallowing, tightness in your throat
- Coughing, wheezing, shortness of breath
- Fainting, dizziness, confusion, or weakness
- Other

Other allergic reaction, please describe: ________________________________

How long after vaccination (in minutes) did the allergic reaction start?

________________________________

What treatment did you receive? Please select all that apply:

- No treatment
- Anti-histamine
- Adrenaline Inhaler
- Prednisolone or other steroids
- Transferred to the Emergency department
- Hospitalisation in normal unit (non-ICU)
- Hospitalisation in Intensive Care Unit (ICU)
- Other

If other, please specify: ________________________________

Please describe what happened: ________________________________________

Other vaccinations

Did you receive any other vaccines since the start of COVIRS?

- Yes
- No

If yes, which vaccine(s) did you receive?

- Diphtheria-tetanus vaccine (ADT Booster)
- Diphtheria-tetanus-pertussis vaccine (Boostrix, Adacel, Tripacel)
- Diphtheria-tetanus-pertussis-polio vaccine (Boostrix-IPV, Adacel Polio, Quadracel)
- Polio vaccine (IPOL)
- Hepatitis B vaccine (Engerix-B, H-B-Vax II)
- Hepatitis A vaccine (Havrix, Avaxim, Vaqta)
- Hepatitis A-hepatitis B vaccine (Twinrix)
- Hepatitis A-typhoid vaccine (Vivaxim)
- Typhoid injected vaccine (Typhim Vi)
- Typhoid oral vaccine (Vivotif Oral)
- Influenza vaccine (Afluria, Fluad Quad, Fluarix, FluQuadri, Influvac, Vaxigrip, Vaxigroup)
- Papillomavirus vaccine (Cervarix, Gardasil)
- Meningococcal vaccine (Menveo, Menactra, MenQuadfi, NeisVac, Bexsero, Trumenba)
- Pneumococcal vaccine (Prevenar, Synflorix, Pneumosil, Pneumovax)
- Japanese encephalitis vaccine (Imojev, JEspect)
- Rabies vaccine (Rabipur)
- Yellow fever vaccine (Stamaril)
- Measles-mumps-rubella (Priorix, M-M-R II, ProQuad)
- Measles-mumps-rubella-varicella (Priorix-tetra, ProQuad)
- Varicella vaccine (Varilrix, Varivax)
- Zoster live vaccine (Zostavaq)
- Zoster non-live vaccine (Shingrix)
- Tuberculosis vaccine (BCG)
- Other

If other, please specify: ___________________________________
