## Supplementary file 2 for "A systems immunology study comparing innate and adaptive immune responses in adults to COVID-19 mRNA (BNT162b2/mRNA-1273) and adenovirus vectored vaccines (ChAdOx1-S) after the first, second and third doses"

### AIM assay gating:

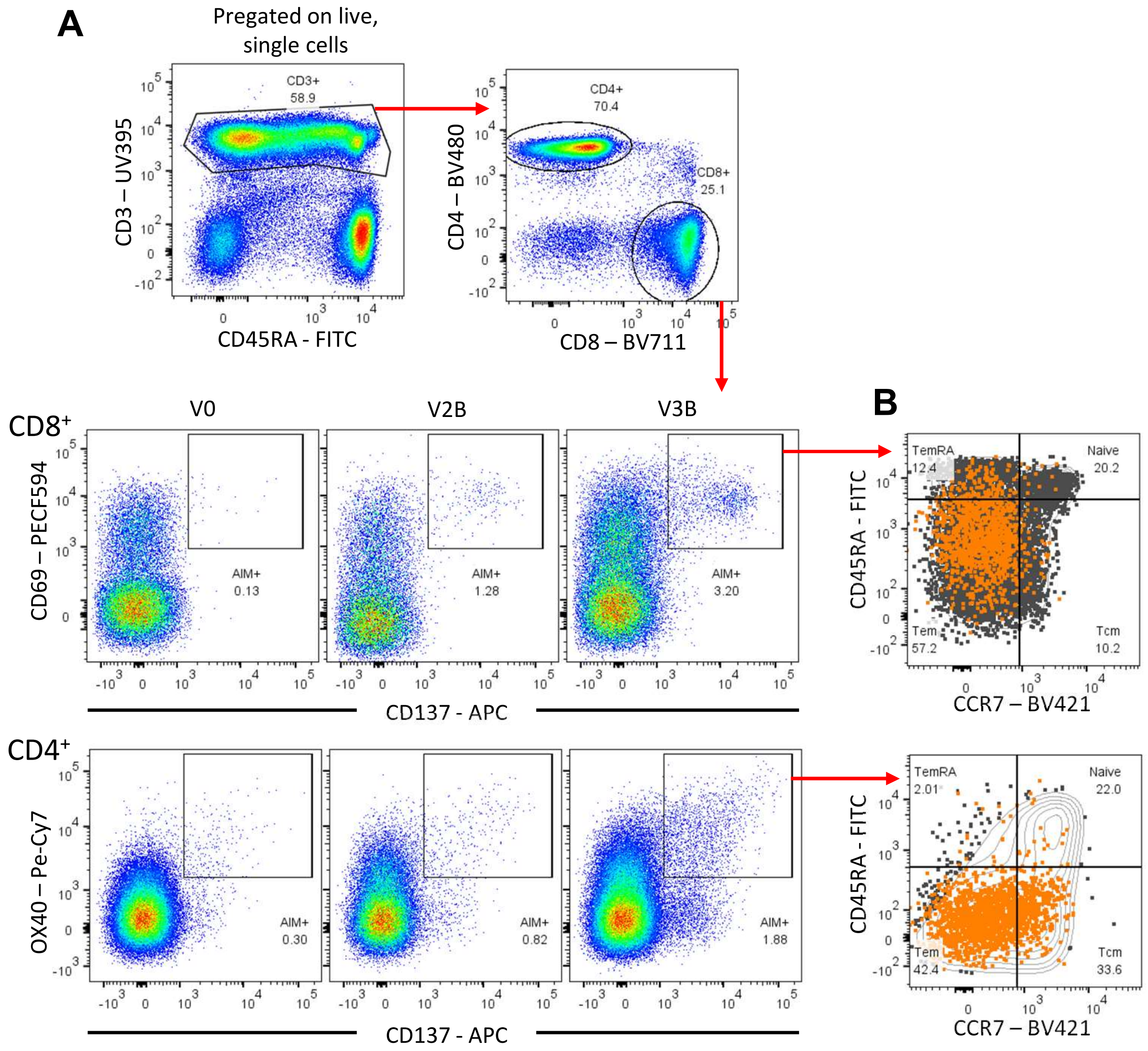


Representative flow cytometry for the identification of activation-induced marker expression on CD4⁺ and CD8⁺ T cells.

**A)** Representative strategie to define CD3+CD4+ and CD3+CD8+ cells and expression of the activation-induced markers CD69 and CD137 on CD8+ T cells and OX40 and CD137 on CD4 T cells at V0, V2B and V3B. **B)** AIM+ memory subsets (orange) were defined based on the expression of CCR7 and CD45RA: central memory (TCM, CCR7+CD45RA−), effector memory (TEM, CCR7-CD45RA−), and terminally differentiated effector cells (TEMRA, CCR7-CD45RA+) overlaid on total CD4 and CD8 T cell subsets.

### Panel 1: Pan Leukocyte gating

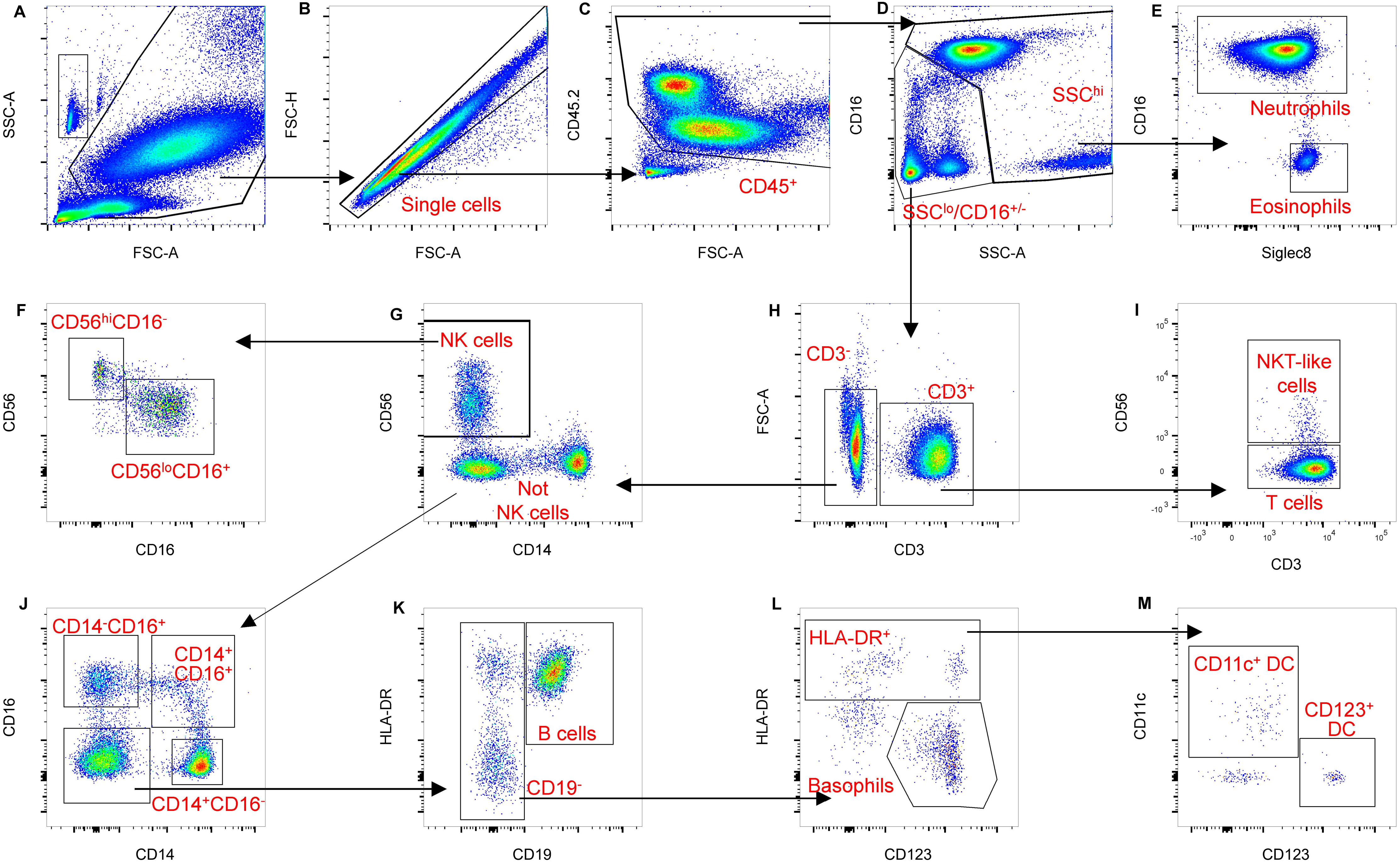

**Panel 1 gating strategy.**

**A)** Leukocytes identified based on FSC and SSC. Count beads also identified by high SSC and low SSC, and counted based on uniform fluorescence into Blue 530/30 and Yellow Green 780/60 channels.
**B)** Doublets eliminated based on FSC and FSH.
**C)** Identification of CD45$^{+}$ leukocytes within single cell population.
**D)** Separation of CD45$^{+}$ cells into CD16$^{+/-}$, high SSC (granulocyte) and CD16$^{+/-}$ low SSC (lymphocyte and monocyte) populations.
**E)** Identification of neutrophils (CD16$^{+}$) and eosinophils (Siglec8$^{+}$CD16$^{-}$) within the CD16$^{+/-}$, high SSC population.
**F)** Identification of CD56$^{high}$CD16$^{-}$ and CD56$^{low}$CD16$^{+}$ natural killer (NK; CD3$^{-}$CD56$^{+}$) cell populations.
**G)** Identification of NK cells (CD56$^{+}$) within the CD3$^{-}$, CD16$^{+/-}$ low SSC population.
**H)** Identification of T-cells (CD3$^{+}$) within CD16$^{+/-}$ low SSC population.
**I)** Identification of natural killer T (NKT) like cells (CD3$^{+}$CD56$^{+}$) and conventional T cells (CD3$^{+}$CD56$^{-}$) within the CD3$^{+}$, CD16$^{+/-}$ low SSC population.
**J)** Identification of classical monocytes (CD14$^{+}$CD16$^{-}$), intermediate monocytes (CD14$^{+}$CD16$^{+}$), and non-classical monocytes (CD14$^{-}$CD16$^{+}$) cells within the CD3$^{-}$, CD56$^{-}$, low SSC population.
**K)** Identification of B-cells (CD19$^{+}$HLA-DR$^{+}$) cells within CD14$^{-}$,CD16$^{-}$, CD3$^{-}$, CD56$^{-}$, low SSC population.
**L)** Identification of dendritic cells (DC; HLA-DR$^{+}$, CD123$^{+/-)}$ and basophils (CD123$^{+}$HLA-DR$^{-}$) within the CD19$^{-}$, CD14$^{-}$,CD16$^{-}$, CD3$^{-}$, CD56$^{-}$, low SSC population.
**M)** Identification of conventional DCs (CD11c$^{+}$) and plasmacytoid DCs (CD123$^{+}$) within the dendritic cell population.

**Panel 2: lymphocyte gating**

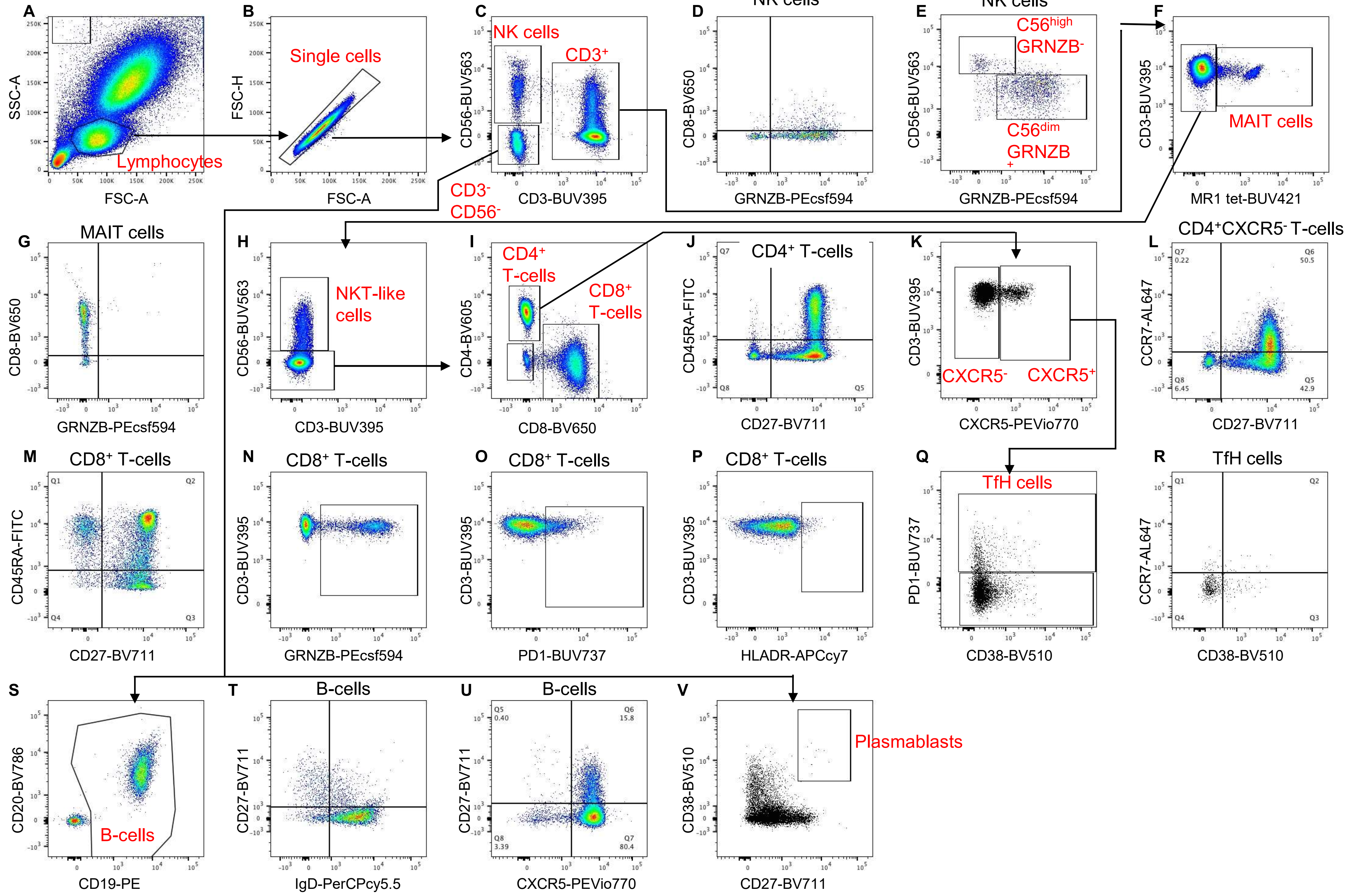

**Panel 2 gating strategy.**

**A)** Lymphocytes identified based on FSC and SSC. Count beads also identified by high SSC and low SSC, and counted based on uniform fluorescence into Blue 530/30 and Yellow Green 780/60 channels.
**B)** Doublets eliminated based on FSC and FSH.
**C)** Identification of natural killer (NK) cells ($CD56^{+}CD3^{-}$), T cells ($CD3^{+}CD56^{+/-}$) and $CD3^{-}CD56^{-}$ lymphocytes.
**D)** Expression of granzyme B and CD8 on NK cells.
**E)** Identification of $CD56^{high}GRNZB^{-}$ and $CD56^{low}GRNZB^{+}$ NK cell populations.
**F)** Identification of mucosal-associated invariant T-cells (MAIT) cells ($CD3^{+}$MR1-tetramer$^{+}$) from $CD3^{+}CD56^{+/-}$ population.
**G)** Expression of granzyme B and CD8 on MAIT cells.
**H)** Identification of natural killer (NKT)-like cells from $CD3^{+}CD56^{+/-}$MR1-tet$^{-}$ population.
**I)** Identification of conventional $CD4^{+}$ and $CD8^{+}$ T-cells from $CD3^{+}$MR1-tet$^{-}CD56^{-}$ population.
**J)** Identification of naïve ($CD45RA^{+}CD27^{+}$), central memory ($CD45RA^{-}CD27^{+}$), effector memory ($CD45RA^{+}CD27^{-}$) and late differentiated ($CD45RA^{-}CD27^{-}$) cells within the $CD4^{+}$ Conventional T-cell population.
**K)** Identification of $CXCR5^{+}$ and $CXCR5^{-}$ cells within the $CD4^{+}$ Conventional T-cell population.
**L)** Expression of CCR7 and CD27 within the $CXCR5^{-}CD4^{+}$ Conventional T-cell population.
**M)** Identification of naïve ($CD45RA^{+}CD27^{+}$), central memory ($CD45RA^{-}CD27^{+}$), effector memory ($CD45RA^{+}CD27^{-}$) and late differentiated ($CD45RA^{-}CD27^{-}$) cells within the $CD8^{+}$ Conventional T-cell population.
**N)** Expression of granzyme B within the $CD8^{+}$ Conventional T-cell population.
**O)** Expression of PD1 within the $CD8^{+}$ Conventional T-cell population.
**P)** Expression of HLA-DR within the $CD8^{+}$ Conventional T-cell population.
**Q)** Identification of circulating T follicular helper (cTfH) cells by PD1 expression on $CXCR5^{+}CD4^{+}$ Conventional T-cells.
**R)** Identification of $CCR7^{-}CD38^{+}$ and $CCR7^{-}CD38^{-}$ populations within cTfH cells.
**S)** Identification of B-cells ($CD19^{+}CD20^{+/-}$) within $CD3^{-}CD56^{-}$ lymphocyte population.
**T)** Identification of naïve ($CD27^{-}IgD^{+}$), $IgD^{+}$ memory ($CD27^{+}IgD^{+}$) and $IgD^{-}$ memory ($CD27^{+}IgD^{-}$) B-cells.
**U)** Expression of CD27 and CXCR5 within the B-cell gate.
**V)** Identification of plasmablasts ($CD19^{+/-}$, $CD20^{-}$, $CD27^{++}CD38^{++}$) within the $CD3^{-}CD56^{-}$ population.
