## Supplementary file 3 for "A systems immunology study comparing innate and adaptive immune responses in adults to COVID-19 mRNA (BNT162b2/mRNA-1273) and adenovirus vectored vaccines (ChAdOx1-S) after the first, second and third doses"

### Immune cell counts

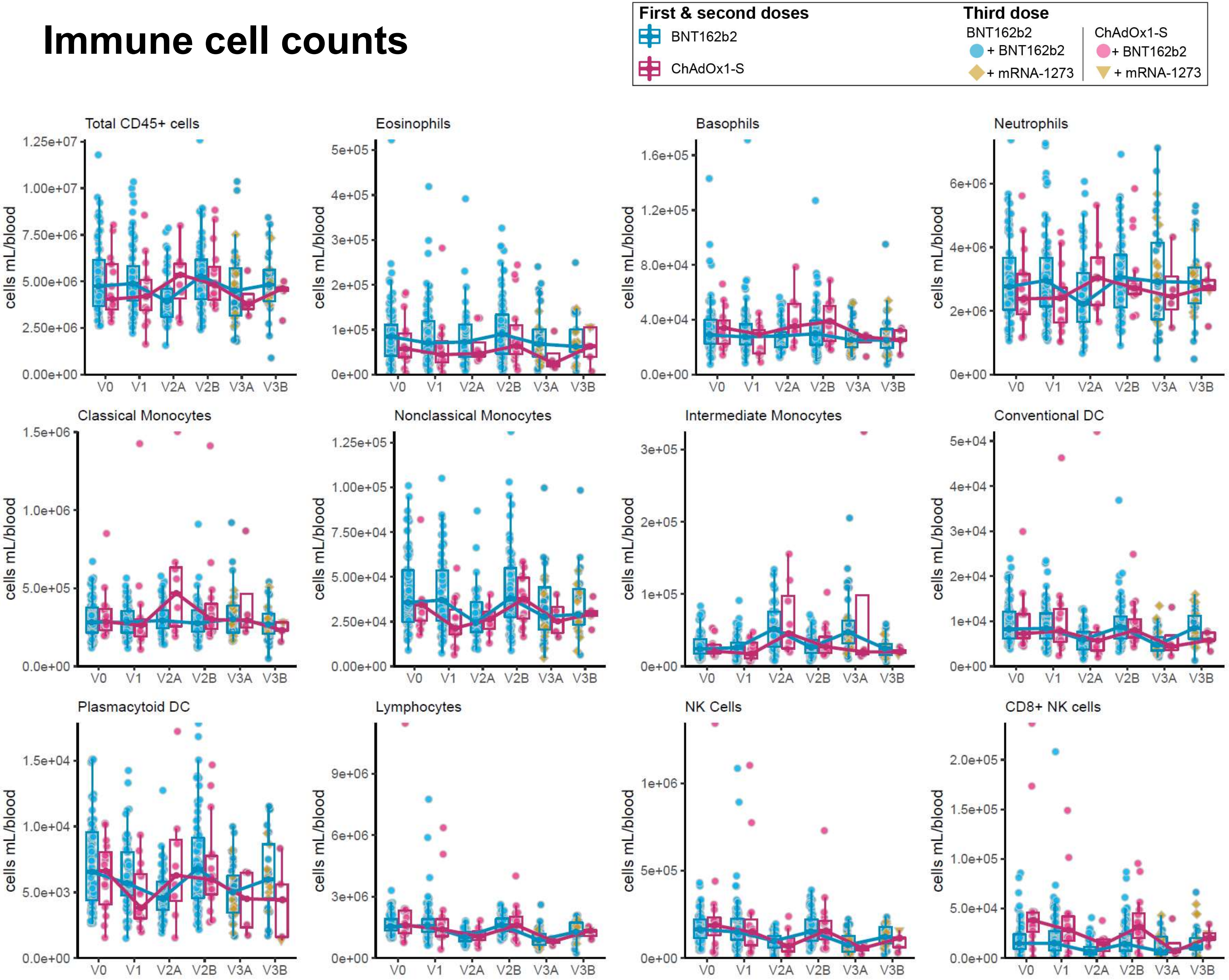

### Immune cell counts

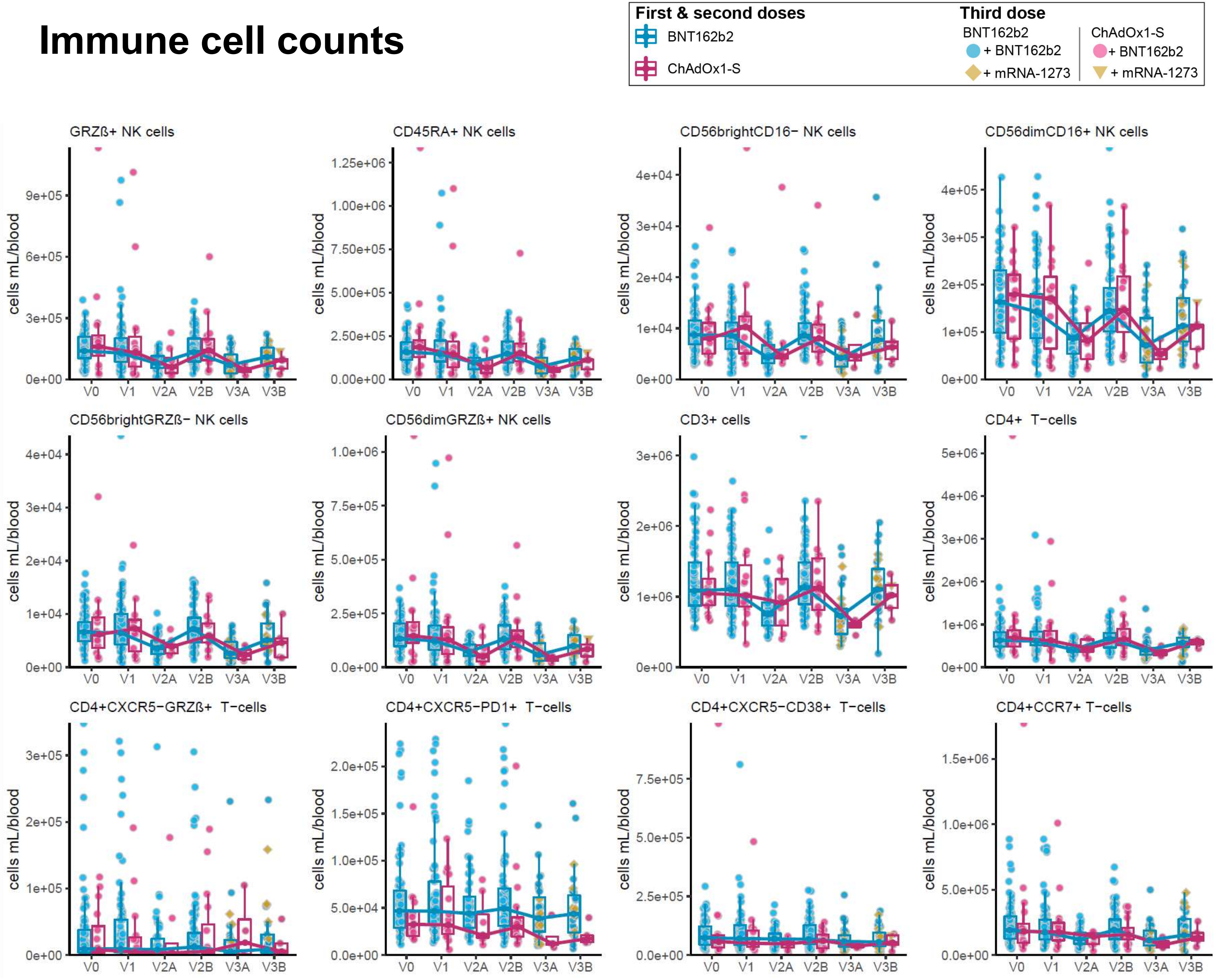

### Immune cell counts

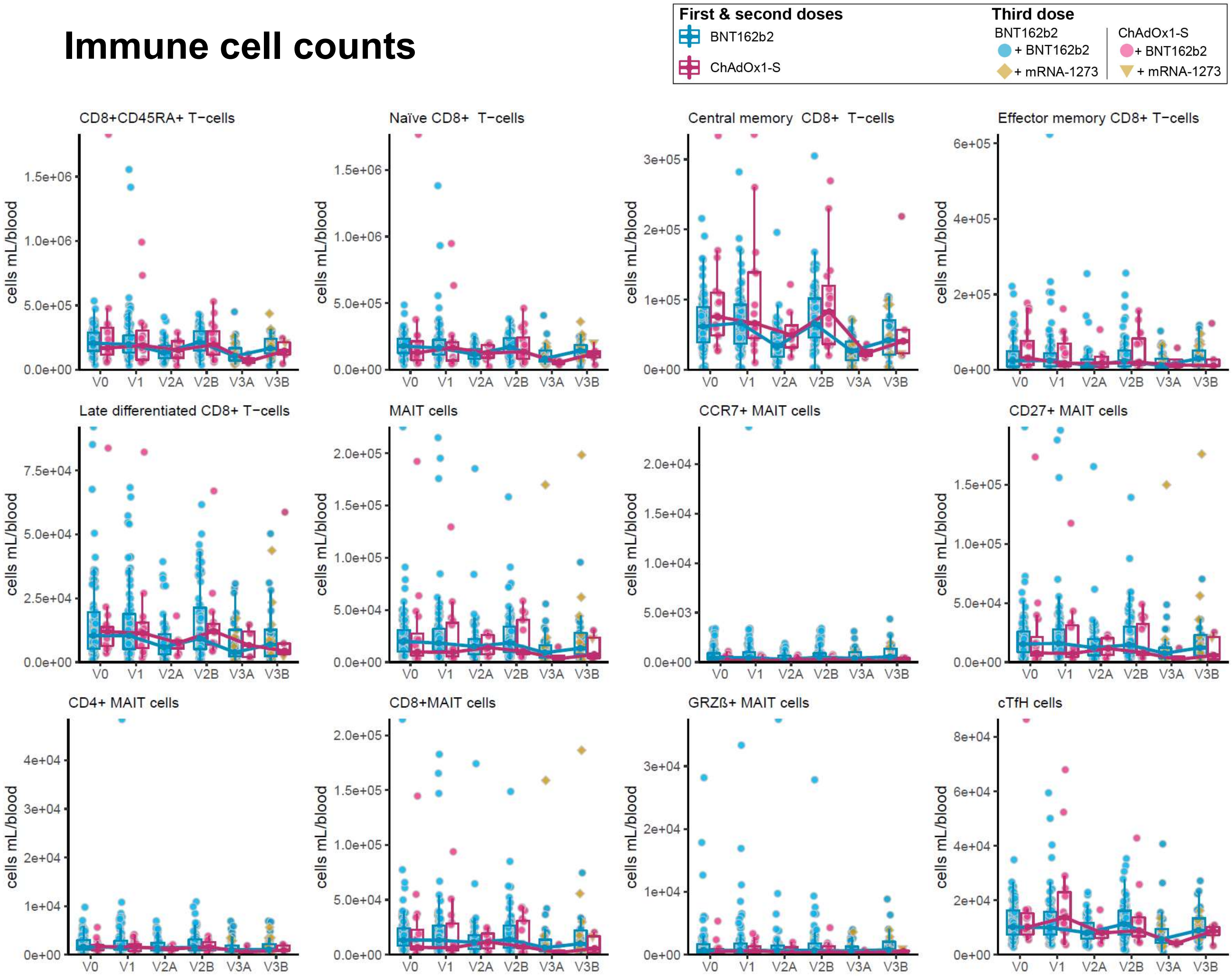

### Immune cell counts

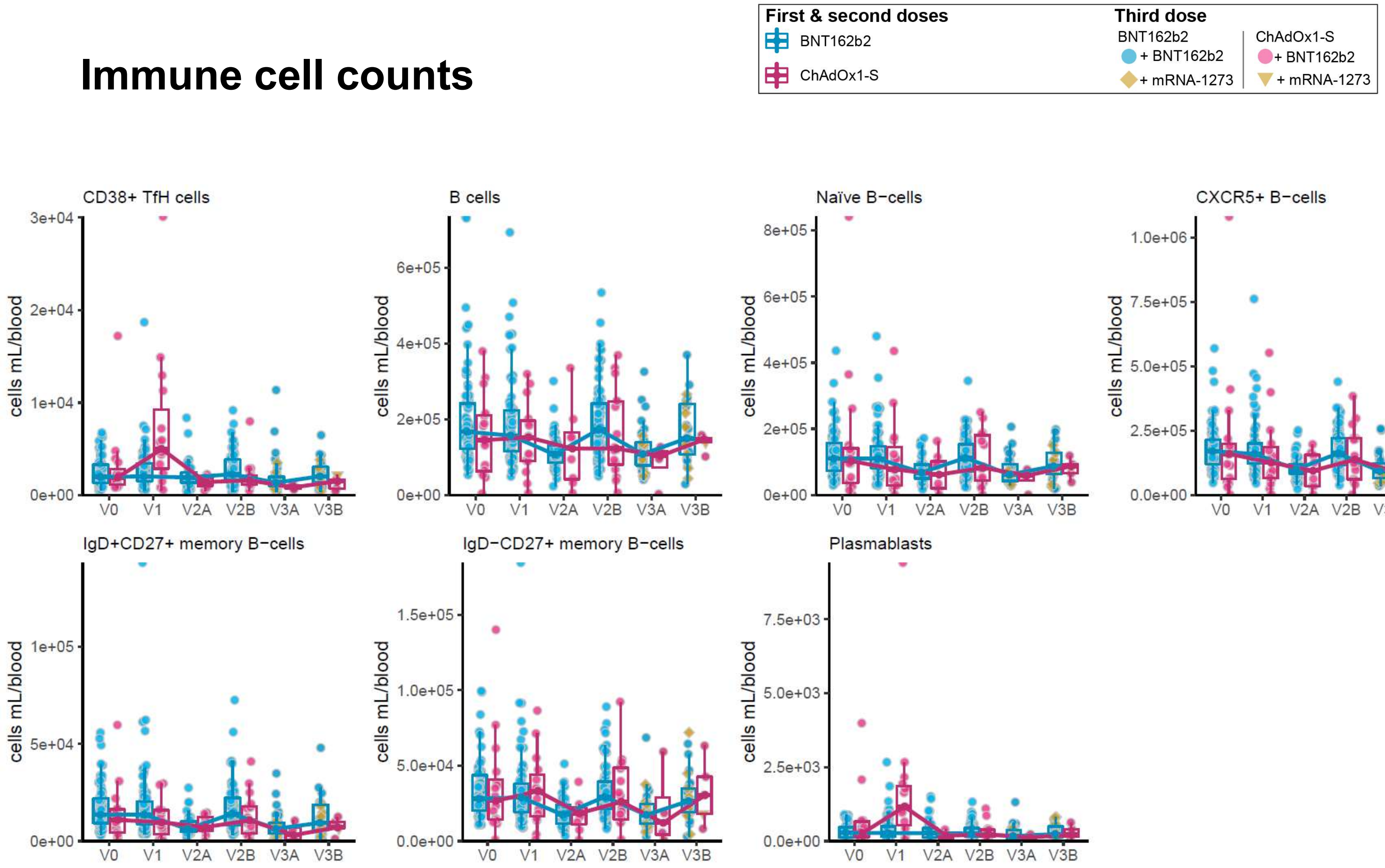

### Immune cell (% of parent)

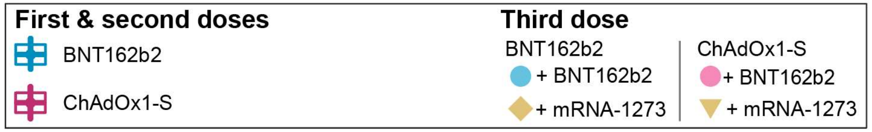


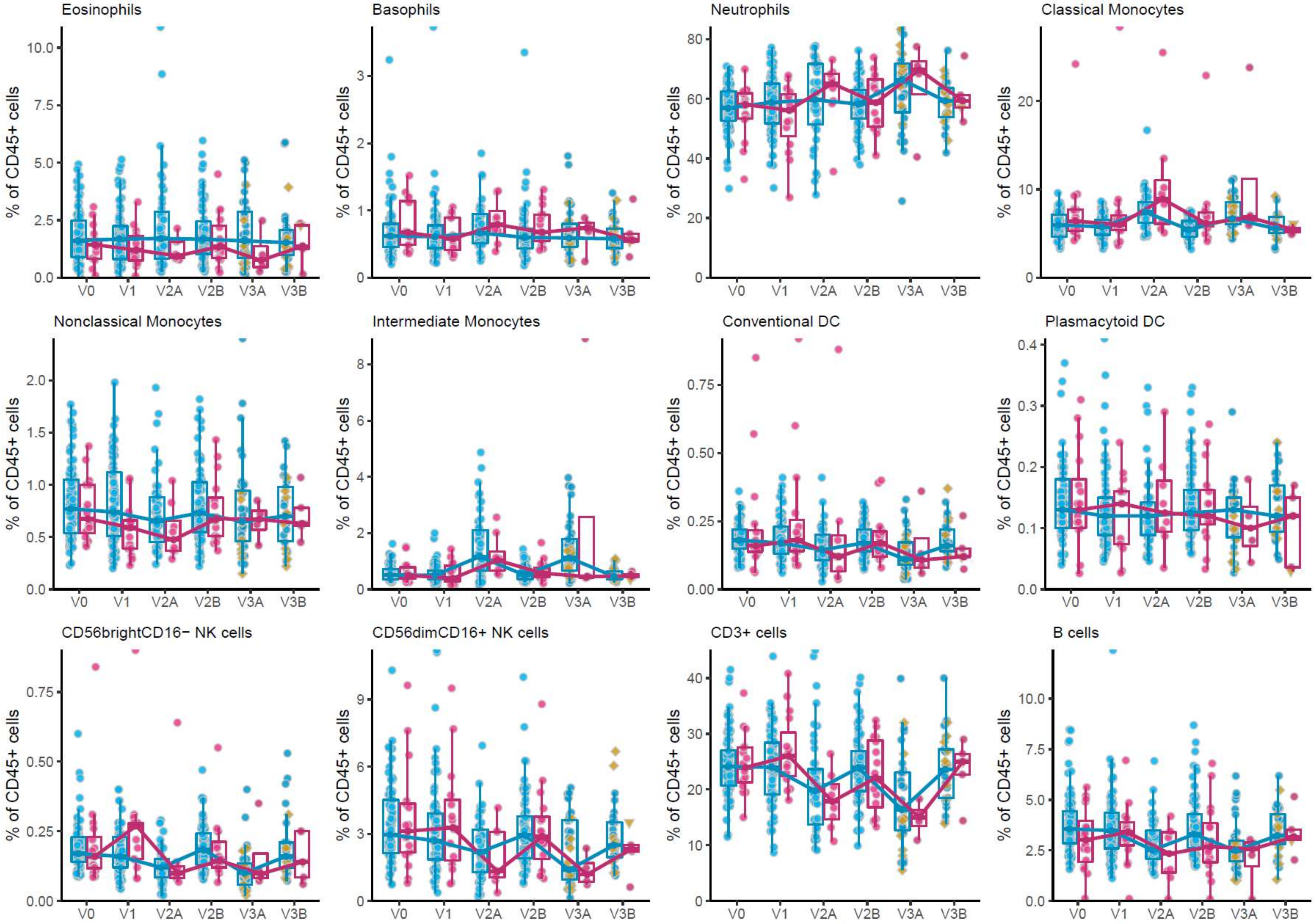

### Immune cell (% of parent)

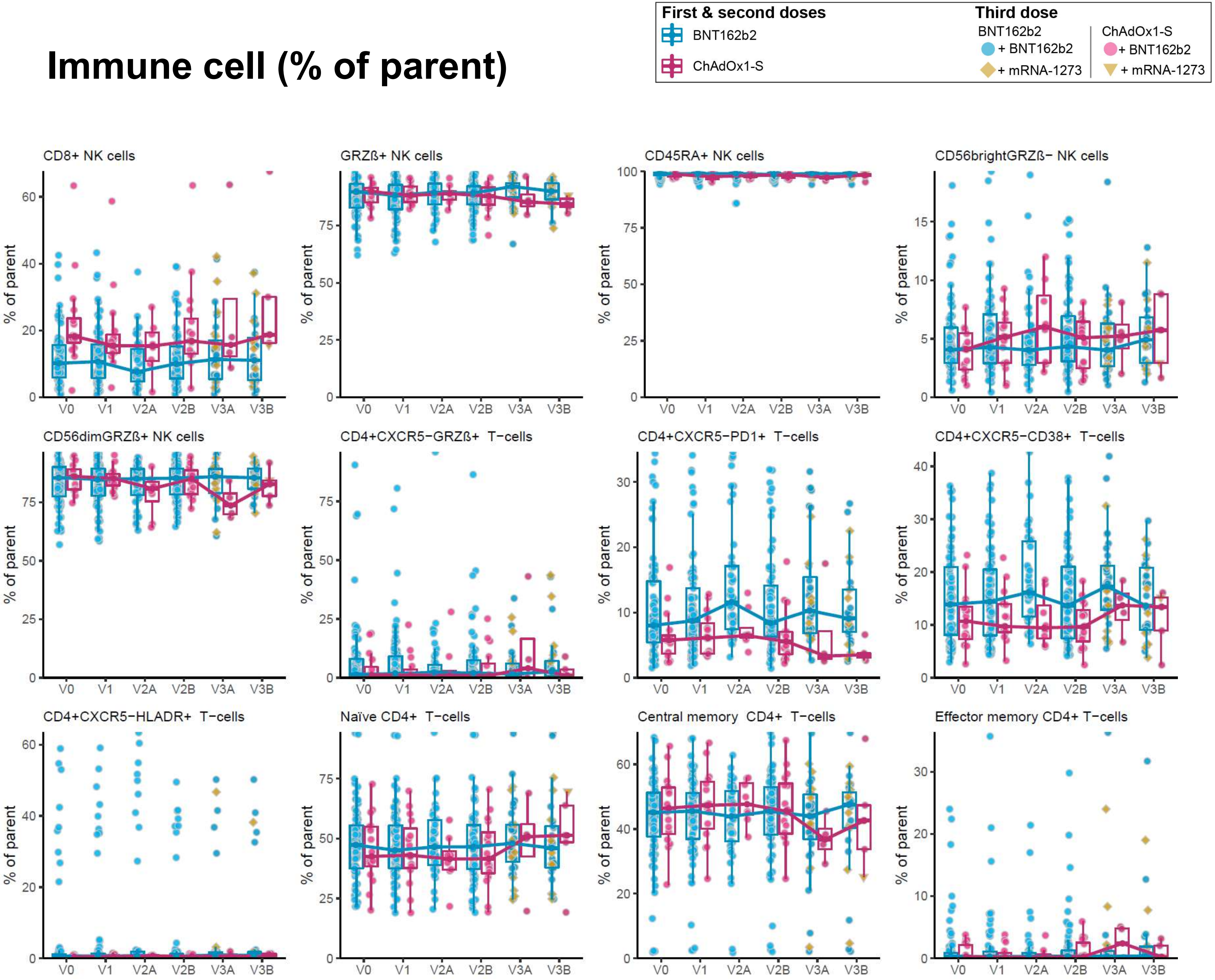

### Myeloid Activation – CD86

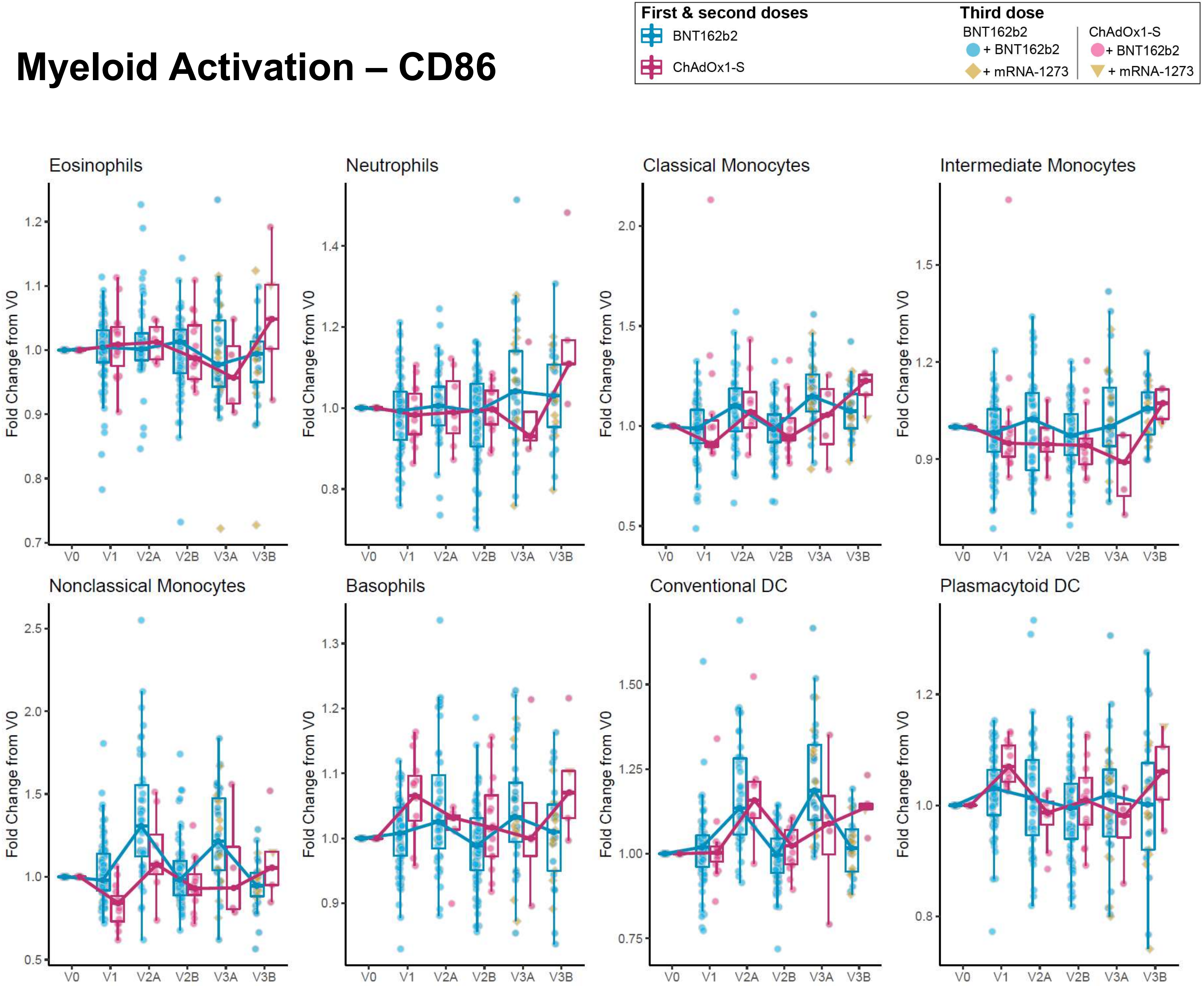

### Myeloid Activation – HLA-DR

First & second doses
BNT162b2
ChAdOx1-S

Third dose
BNT162b2
+ BNT162b2
+ mRNA-1273
ChAdOx1-S
+ BNT162b2
+ mRNA-1273

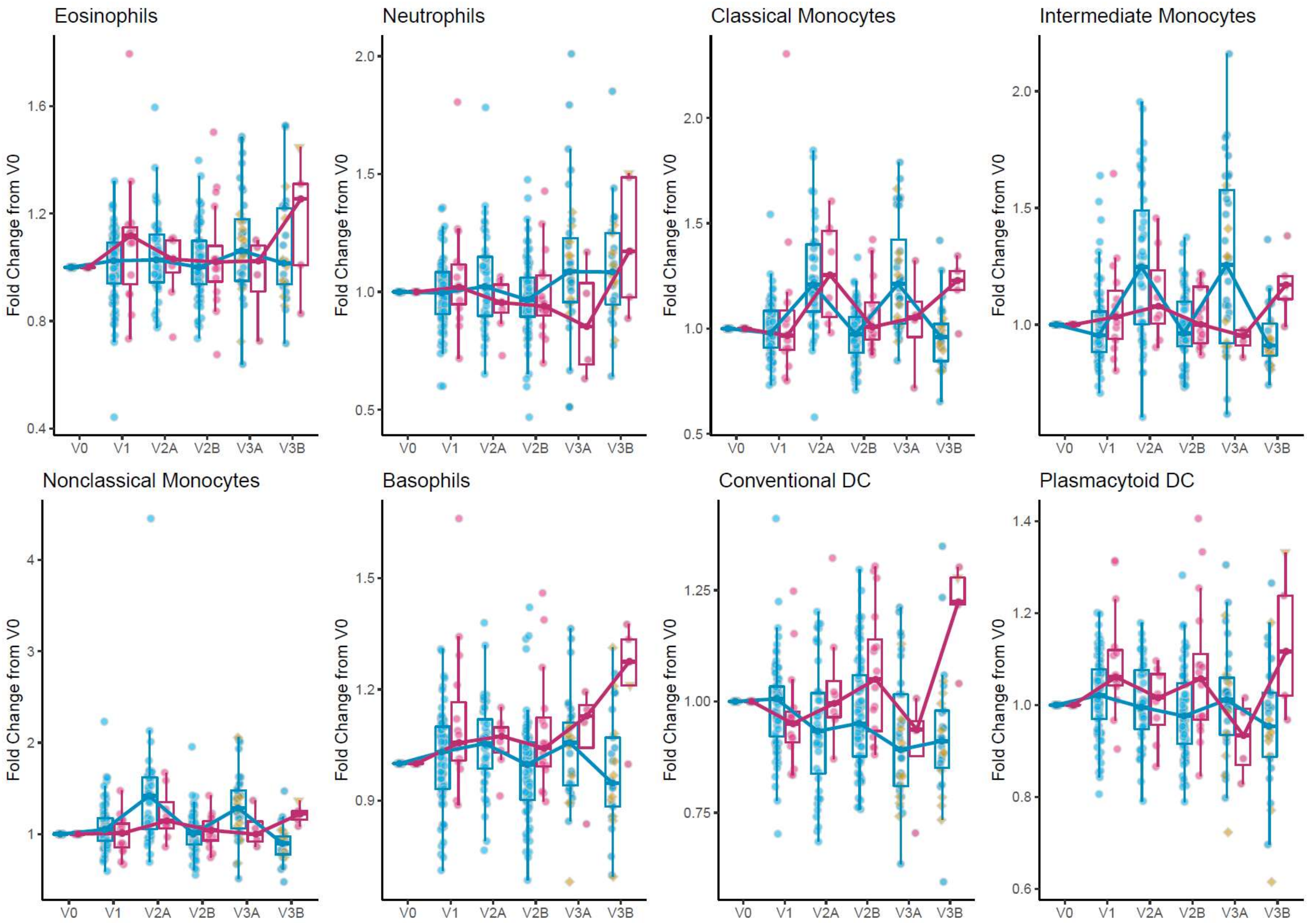

### Immune cell frequency

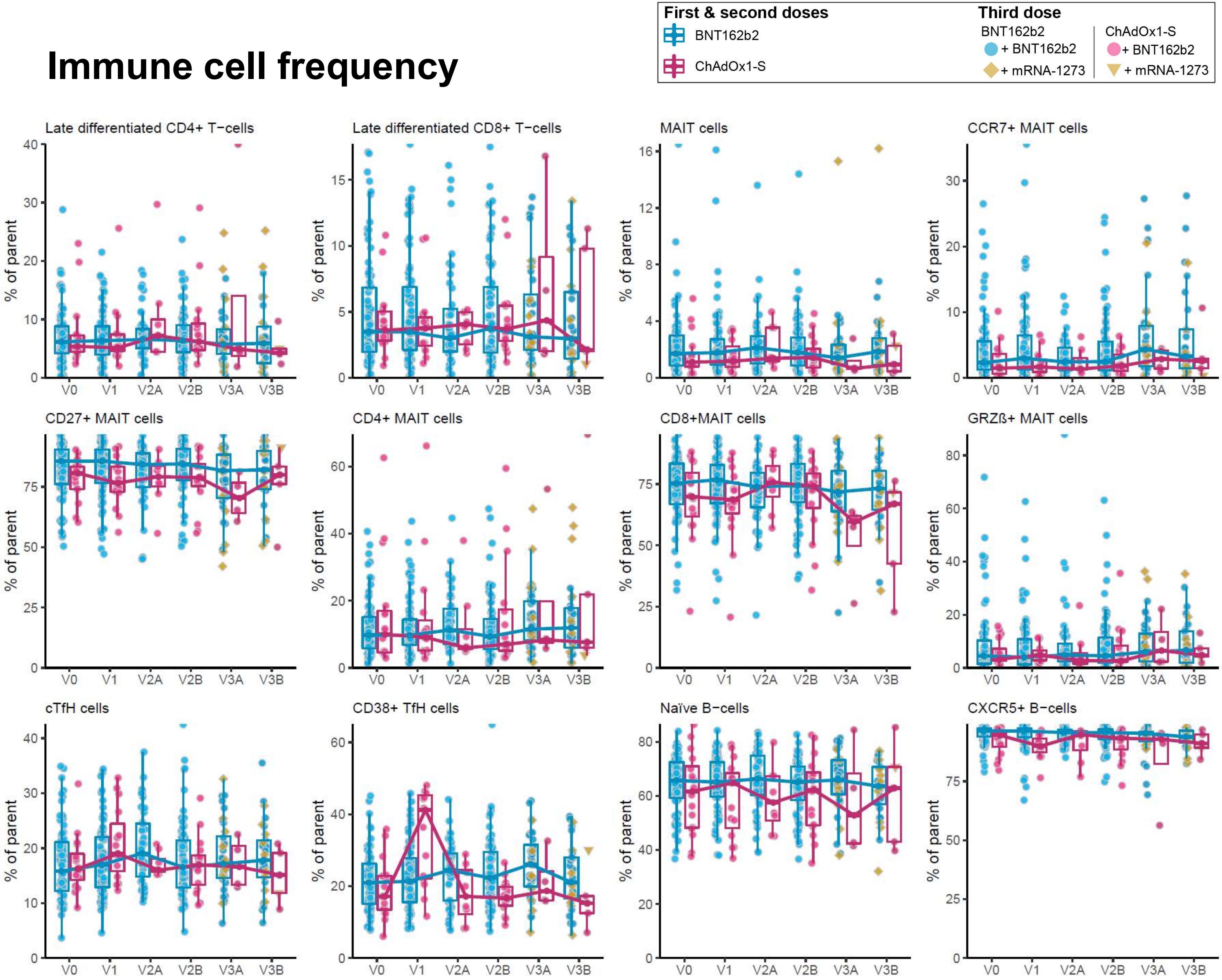
