## Supplementary material for "A systems immunology study comparing innate and adaptive immune responses in adults to COVID-19 mRNA (BNT162b2/mRNA-1273) and adenovirus vectored vaccines (ChAdOx1-S) after the first, second and third doses": STROBE Checklist

STROBE Statement—Checklist of items that should be included in reports of cohort studies

| **Item** | | **Criteria met** |
| --- | --- | --- |
| 1 | Title and abstract | **X** |
| 2 | Introduction - Background/rationale | **X** |
| 3 | Intoruction -Objectives | **X** |
| 4 | Methods - Study Design | **X** |
| 5 | Methods - Setting | **X** |
| 6 | Methods - Participants | **X** |
| 7 | Methods - Variables | **X** |
| 8 | Methods - Data sources/measurement | **X** |
| 9 | Methods - Bias | **X** |
| 10 | Methods - Study size | **X** |
| 11 | Methods - Quantiative variables | **X** |
| 12 | Methods - Statistical methods | **X** |
| 13 | Results - Participants | **X** |
| 14 | Results - Descriptive data | **X** |
| 15 | Results - Outcome data | **X** |
| 16 | Results - Main results | **X** |
| 17 | Results - Other analyses | **X** |
| 18 | Discussion - Key results | **X** |
| 19 | Discussion - Limitations | **X** |
| 20 | Discussion - Interpretation | **X** |
| 21 | Discussion - Generalisability | **X** |
| 22 | Other information - Funding | **X** |
